# Pneumococcal Serotype Prevalence in Southeast Asia: A Systematic Review and Meta-Analysis

**DOI:** 10.1101/2023.07.20.23292974

**Authors:** Alex J. J. Lister, Evelin Dombay, David W. Cleary, Stuart C. Clarke

## Abstract

**Background:** The prevalence of *Streptococcus pneumoniae* serotypes in the ASEAN region is not well studied despite severe pneumococcal infections being a major cause of death among children in the region. This information is crucial for understanding the epidemiology of the disease and guiding vaccine policies. Our study aimed to provide a thorough analysis of the pneumococcal serotypes in ASEAN prior to vaccine introduction to assist countries in formulating evidence-based vaccine policies.

**Methods:** We conducted a systematic review and meta-analysis of studies reporting *S. pneumoniae* serotypes from carriage, invasive disease (IPD) and non-invasive disease (non-IPD) published up until 31^st^ December 2022 using PubMed, PubMed Central, Ovid MEDLINE and Scopus databases, reference lists and search engines. Data prior to the national introduction of conjugate vaccines in the ASEAN region were included. Non-English, animal, case studies, reviews, and studies on antibiotic resistance only were excluded. The quality of studies was examined using the CASP and the JBI’s Critical Appraisal Tools. The frequency of serogroups and serotypes was calculated, and vaccine coverage was estimated by the addition of vaccine serotypes as a fraction of the total number of isolates per age group. This study was registered with PROSPERO CRD42022243994.

**Findings:** A total of 940 studies were identified, and 99 and 84 relevant papers were included in the qualitative and quantitative analyses, respectively. A total of 16,396 isolates were identified, spread across all body sites and ages, with vaccine-covered serotypes 19F (n = 2,061, 12.57% [95%CI: 12.07 – 13.09]), 23F (n = 1,508, 9.20% [95%CI: 8.76 – 9.65]) and 6B (n = 1,160, 7.07% [95%CI: 6.69 – 7.48]) occurring most often. Non-vaccine types *e.g*., 6AB (n = 617, 3.76% [95%CI: 3.48 – 4.07]), 15BC (n = 35, 2.57% [95%CI: 2.33 – 2.82]) and 34 (n = 260, 1.59% [95%CI: 1.40 – 1.79]) were also frequently observed.

**Interpretation:** The most common serotypes found in IPD, non-IPD, and carriage in ASEAN are covered by currently available conjugate vaccines. This underscores the importance of vaccination and predicts future success in reducing the burden of pneumococcal disease. The data gathered offers important insights into pneumococcal serotype epidemiology across the different countries belonging to ASEAN.

## Introduction

*Streptococcus pneumoniae* is a bacterial pathogen responsible for potentially life-threatening diseases, such as meningitis, pneumonia, and septicaemia collectively referred to as invasive pneumococcal diseases (IPD). Additionally, infections can result in less severe, non-invasive diseases (non-IPD) including sinusitis and otitis media (OM). The bacteria can be carried asymptomatically in the upper respiratory tract (URT) as part of the normal microflora (1). Asymptomatic carriage can lead to pneumococcal disease if the bacteria cross the mucosal membrane and evade the host immune system (2). While the likelihood of developing pneumococcal disease from carriage is low, it can still contribute to the transmission thereby increasing the overall burden of pneumococcal disease in the population (3). Pneumococcal disease burden and mortality are the highest in low- and middle-income countries (LMICs) due to factors including poverty and malnutrition, limited access to healthcare, high prevalence of antibiotic resistant pneumococci and lower vaccination rates and/or coverage (4, 5). Without improvements in the latter, the burden of pneumococcal disease remains high.

Pneumococcal vaccines have been developed by leveraging the immunogenicity of the pneumococcal capsule, which defines over 100 different serotypes currently recognised (6). The differences in the polysaccharide composition of the capsule allows for serotype-specific protection through vaccination. There are currently two types of pneumococcal vaccines available. The 23-valent pneumococcal polysaccharide vaccine (PPV23) is effective against 23 serotypes and is recommended for all adults aged 65 years and older. Age-specific vaccination with PPV23 is necessary because polysaccharides lack T-cell stimulating activity, which makes it difficult to generate an effective immune response in children and antibody-deficient adults (7). The other formulation, polysaccharide conjugate vaccines (PCVs), such as Synflorix ® (PCV10, GlaxoSmithKline) and Prevnar 13® (PCV13, Pfizer), contain semi-purified capsular polysaccharides from 10 and 13 pneumococcal serotypes, respectively, covalently linked to a protein carrier. These protein carriers elicit a T-cell-dependent immune response, making conjugate vaccines more effective in children (8, 9). The first pneumococcal conjugate vaccine, PCV7 is no longer in use. The serotypes covered by PCV10 are 1, 4, 5, 6B, 7F, 9V, 14, 18C, 19F, and 23F. This vaccine is licensed for use in infants and children from 6 weeks up to 5 years of age (10). A higher valency PCV, the 13-valent vaccine covers the same serotypes as PCV10, with the extension to serotypes 3, 6A and 19A (11). PCV13 and the newer PCV15 are currently recommended by the World Health Organization (WHO) for use in all countries that have not yet introduced pneumococcal conjugate vaccines into their routine immunization programs (12). PCV15 (Vaxneuvance, Merck), has been approved for use as of June 2022 and is being licensed throughout the European Union since 2021. It contains the same serotypes as PCV13 plus serotypes 22F and 33F (13) and it is currently recommended for persons aged <19 years (14, 15). Higher valent vaccines are entering the market, with the twenty-serotype vaccine (PCV20, Apexxnar R, Pfizer) being approved for use in the >18 years of age population by the European Medicines Agency and the Food and Drug Administration (13, 16–19). This vaccine contains the serotypes found in PCV15, with the addition of serotypes 8, 10A, 11A, 12F, and 15B and is recommended to countries that have already implemented PCV to prevent the emergence of non-vaccine types (NVTs).

Between 2010 and 2019, it is estimated that PCV13 vaccination prevented 175.2 million cases of pneumococcal diseases and 624,904 deaths worldwide (20). According to the World Health Organization (WHO), as of 2021, more than 150 countries have introduced pneumococcal conjugate vaccines into their routine immunization programs. However, the extent of implementation varies. Some countries have PCVs as part of their routine immunization schedule, while others use it on an *ad hoc* basis where individuals are given a choice to purchase the vaccine. Unfortunately, certain low- and middle-income countries (LMICs) are unable to adopt vaccination due to financial constraints and vaccine policies needing review. The paucity of country-specific data on disease burden and the cost of pneumococcal conjugate vaccines (PCVs) are among the primary reasons for the slow adoption of PCVs in LMICs (21).

Ten nations in the South-East Asian region form the Association of Southeast Asian Nations (ASEAN) and are amongst those with the highest burden of childhood pneumonia globally. The incidence of pneumonia in children in Southeast Asia is 2,500 cases per 100,000 children which is higher than the global average of 1,400 cases per 100,000 children according to data by the UNICEF (22). The disparity in vaccine availability discussed above is seen in Southeast Asian countries. In Thailand, PCV13 was first introduced in 2011, not currently as part of the routine immunization programme but it can be purchased at private clinics and hospitals (23). In Malaysia, PCVs were similarly available from around 2005 through private healthcare facilities. However, in 2020 Malaysia took the next step and included PCV10 in their NIPs. Other Southeast Asian countries that have introduced PCV in their NIPs are Cambodia, the Philippines, Myanmar and Laos, and there has been partial introduction in Indonesia. Vaccines are given to certain risk groups in Brunei, and Vietnam is yet to introduce a PCV.

The primary objective of this study was to quantify the importance of PCV vaccine implementation and/or amendment in the remainder of ASEAN countries by providing a comprehensive review of the prevalence of pneumococcal serotypes across the region and an update on the 2012 review in the region (24). Using a systematic approach, this study aims to estimate the vaccine coverage in countries where PCV has been implemented and in those where the vaccine is yet to be introduced. With the evidence gathered in this study, we aim to inform policy makers about the importance of pneumococcal vaccination in reducing the burden and mortality of pneumococcal diseases so that further considerations can be made on an improved vaccine policy.

## Methods

Online databases PubMed, PubMed Central, Ovid MEDLINE and Scopus were systematically searched for literature regarding the serotype prevalence of *S. pneumoniae* across the Southeast Asian countries belonging to ASEAN. No lower date limit was set and included papers published up until the 31^st^ December 2022. Search terms included: ‘Brunei’, ‘Cambodia’, ‘Indonesia’, ‘Malaysia’, ‘Myanmar’, ‘Laos’, ‘Philippines’, ‘Singapore’, ‘Thailand’, ‘Timor’, ‘Vietnam’, ‘SE Asia*’, ‘South East Asia*’, Southeast Asia*’, ‘Southeastern Asia*’, ‘Streptococcus pneumoniae’, ‘S pneumoniae’, ‘pneumococc*’, ‘pneumonia’, ‘serotype*’, ‘serogroup*’, ‘seroprevalence’, separated by the binary operators ‘OR’ and ‘AND’. Asterisks represent a truncation of the term.

Inclusion criteria included full texts that reported *S. pneumoniae* serotypes in countries that belonged to ASEAN, those which reported both invasive, non-invasive disease data with the inclusion of carriage studies regardless of age or gender reported. Both serogroup and serotype data were included. Exclusion criteria of studies included those that were not written in English, animal studies, case studies, reviews, studies that only reported strains with antibiotic resistance, studies of single serotypes, articles on biochemical techniques or genetics for serotyping, non-ASEAN studies, and studies which only reported infection numbers or antibody levels without reporting serotypes. From the initial search, returned papers were scanned for duplicates, which were then removed. Titles and abstracts were analysed, and any papers deemed not relevant were removed. Remaining papers’ full texts were then reviewed by two researchers, using the criteria for inclusion/exclusion in the qualitative analysis. Studies that then met inclusion criteria for quantitative analysis were then reviewed and serotype data was extracted. Exclusion criteria for quantitative analysis included studies that were conducted after the respective countries’ date of PCV implementation and those studies that overlapped with others already included.

The assessment of articles from a quality perspective was undertaken using a by using a consolidation of two checklist-style quality assessment frameworks: The Critical Appraisal Skills Programme (CASP) checklists and the Joanna Briggs Institute Critical Appraisal Checklist for Prevalence Studies (25, 26).

Country data was grouped into source type categories (IPD, non-IPD and carriage). IPD isolates were defined as samples taken from a normally sterile site such as blood or cerebrospinal fluid (CSF), non-IPD isolates were defined as those taken from a non-sterile site and experienced non-invasive disease such as bronchitis, otitis media and other mild pneumococcal infection. Carriage isolates were defined as those taken from patients experiencing neither IPD nor non-IPD, including generally healthy populations. Serotypes and groups were classed as vaccine type (VT) serotypes, which were those serotypes found in the PCVs. Non-vaccine types (NVT), unencapsulated non-typeable (NT) serotypes and unknown (N/A) serotypes formed a separate category. Unknown serotypes were classed as those that could not be determined through methodologies or from studies which do not explicitly state the specific serotype. Some studies reported serotypes that were either VTs or NVTs but remained unidentified, so were grouped into additional ‘PCV related unknown’ and ‘non-PCV related unknown’ serotypes. Serotype prevalence was stratified by age categories including ‘under-five years’, ‘over five years’ and ‘unreported’, the latter used for studies that did not specify the age of the participant or did not fit the over and under five classifications, such as data that spanned the age ranges without further details.

GraphPad Prism 9 (GraphPad Software, Inc.) was used for data management and graph construction. Numbers of isolates were grouped into their respective serotypes under each country, as well as by age category. Percentage coverage of the vaccines was calculated by the addition of vaccine serotypes as a fraction of the total number of isolates per age group. Percentages were expressed with 95% confidence intervals (95%CI) in tabular and boxplot format. GraphPad Prism 9 was used for graph construction and to perform the statistical analysis.

The funder of the study had no role in the study design, data collection, data analysis, data interpretation or writing of the report.

## Results

In total, 940 articles were identified. The initial database search resulted in n = 934 articles (PubMed = 396, Ovid Medline = 153, Scopus = 294, PMC = 91) with the remaining six identified from search engine keywords. From this collection, n = 452 studies were duplicates. Screening abstracts and titles resulted in n = 389 records being excluded. From the identified articles, n = 99 met the inclusion criteria for qualitative analysis (Table 1) and of these, n = 84 were included in the quantitative analysis (Figure 1).

**Table 1.** Qualitative analysis of studies investigating pneumococcal serotype prevalence in ASEAN countries *Study omitted to avoid risk of duplication. **Study omitted due to serotype data that could not be determined from the manuscript. ***Study omitted due to serotypes coming from post-PCV in population. ****Study omitted due to no full text

**Figure 1.**
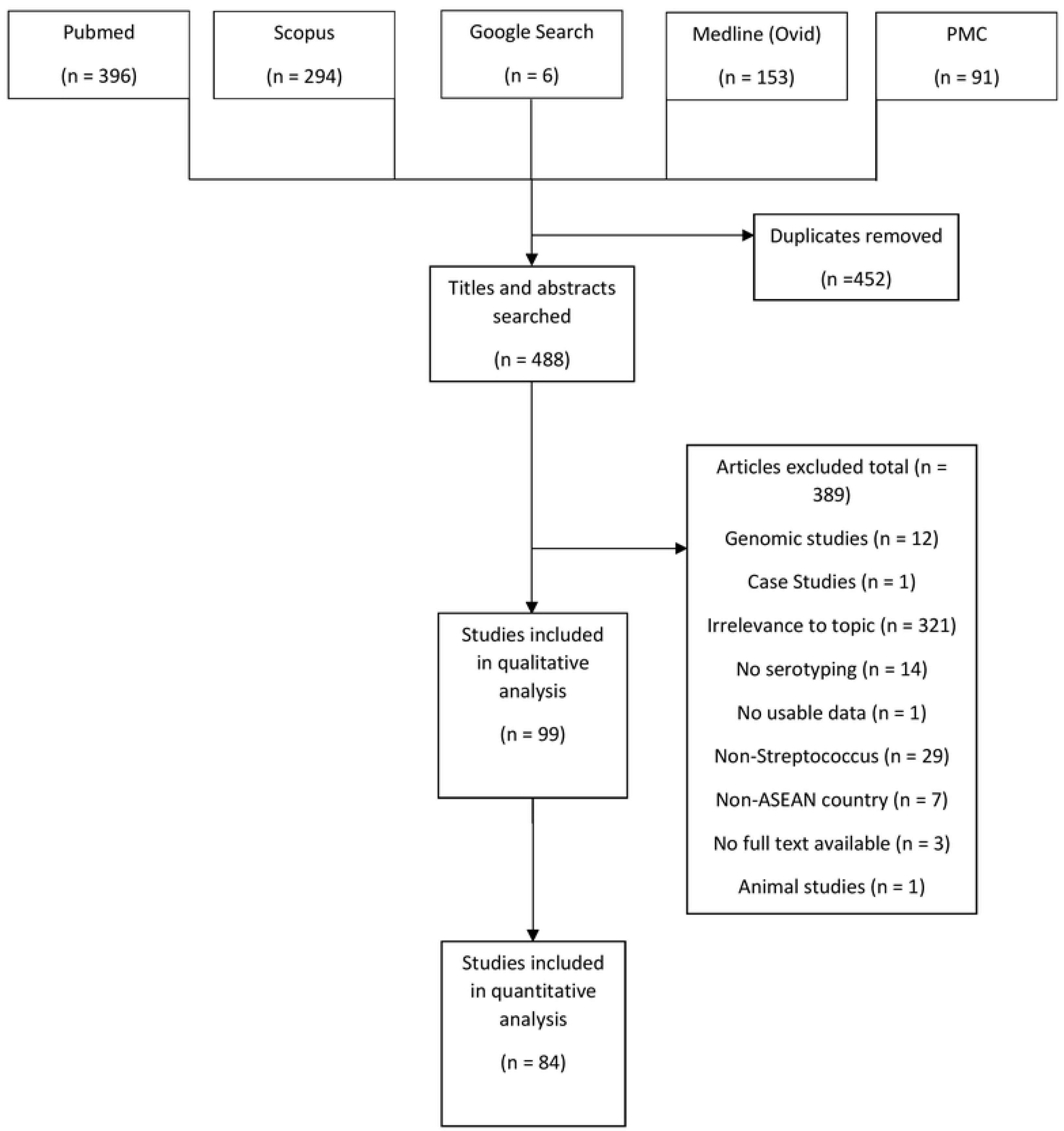
PRISMA Statement showing search and study selection.

It was decided that the study on the Thailand / Myanmar border (27) would be added to the Myanmar data as, despite residing in a refugee camp in Thailand at time of study, the population were originally refugees from Myanmar. The results from all studies were broken down by country.

### All Countries

A total of n = 16,396 isolates were identified with n = 4,129 (25.18% [95%CI: 24.52 – 25.85]) from IPD, n = 1,752 isolates (10.69%, [95%CI:10.22 – 11.17]) from non-IPD, and n = 10,053 isolates from carriage (61.31%, [95%CI: 60.56 – 62.06]) (Table 2). The remaining n = 462 (2.82%, [95%CI: 2.57 – 3.08]) were from unknown sources. The most common vaccine types across all sources were 19F (n = 2,061, 12.57% [95%CI: 12.07 – 13.09]), followed by 23F (n = 1,508, 9.20% [95%CI: 8.76 – 9.65]) and 6B (n = 1,160, 7.07% [95%CI: 6.69 –7.48]) (Table 2). The three most common NVTs were 6AB (n = 617, 3.76% [95%CI: 3.48 – 4.07]), 15BC (n = 35, 2.57% [95%CI: 2.33 – 2.82]) and 34 (n = 260, 1.59% [95%CI: 1.40 – 1.79]). PCV10 and PCV13, PCV15 and PCV20 serotypes accounted for 40.06% [95%CI: 39.31 – 40.81], 49.93% [95%CI: 49.16 – 50.70], 50.22% [95%CI: 49.45 – 50.99] and 51.95% [95%CI: 51.18 – 52.71] respectively. Non-typeable isolates totalled n = 1,708 (10.42%, [95%CI: 9.95 – 10.89]) and unknown serotypes, including those that were reported as autoagglutinated, totalled n = 2,350 (14.33% [95%CI: 13.80 – 14.88]) of total isolates. Fig2, Fig 3, Fig4 and Fig5 show the split of serotype counts from across IPD, non-IPD, carriage and unknown sources.

**Table 2.** Serotype counts and percentages for all countries in ASEAN, split by age category and invasive, non-invasive, carriage sources and from sources which could not be determined.

**Fig2. Serotypes.**
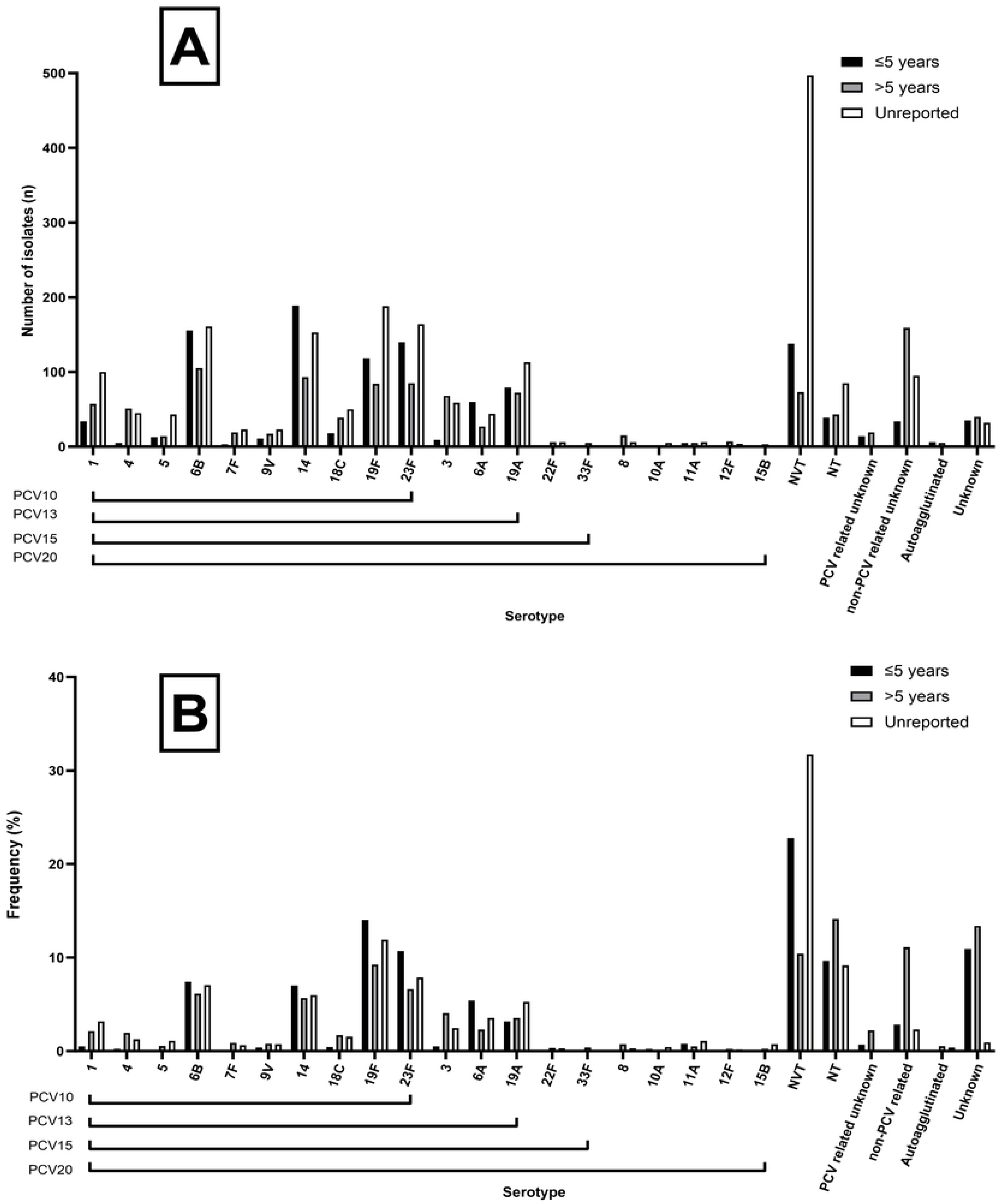
from across ASEAN invasive disease studies. (A) Number of isolates for each respective serotype found in PCV10, 13, 15 and 20, including number of NVTs and unknown serotypes from identified studies on IPD. (B) Frequency of isolates for each respective serotype found in PCV10, 13, 15 and 20, including number of NVTs and unknown serotypes from identified studies.

**Fig3. Serotypes.**
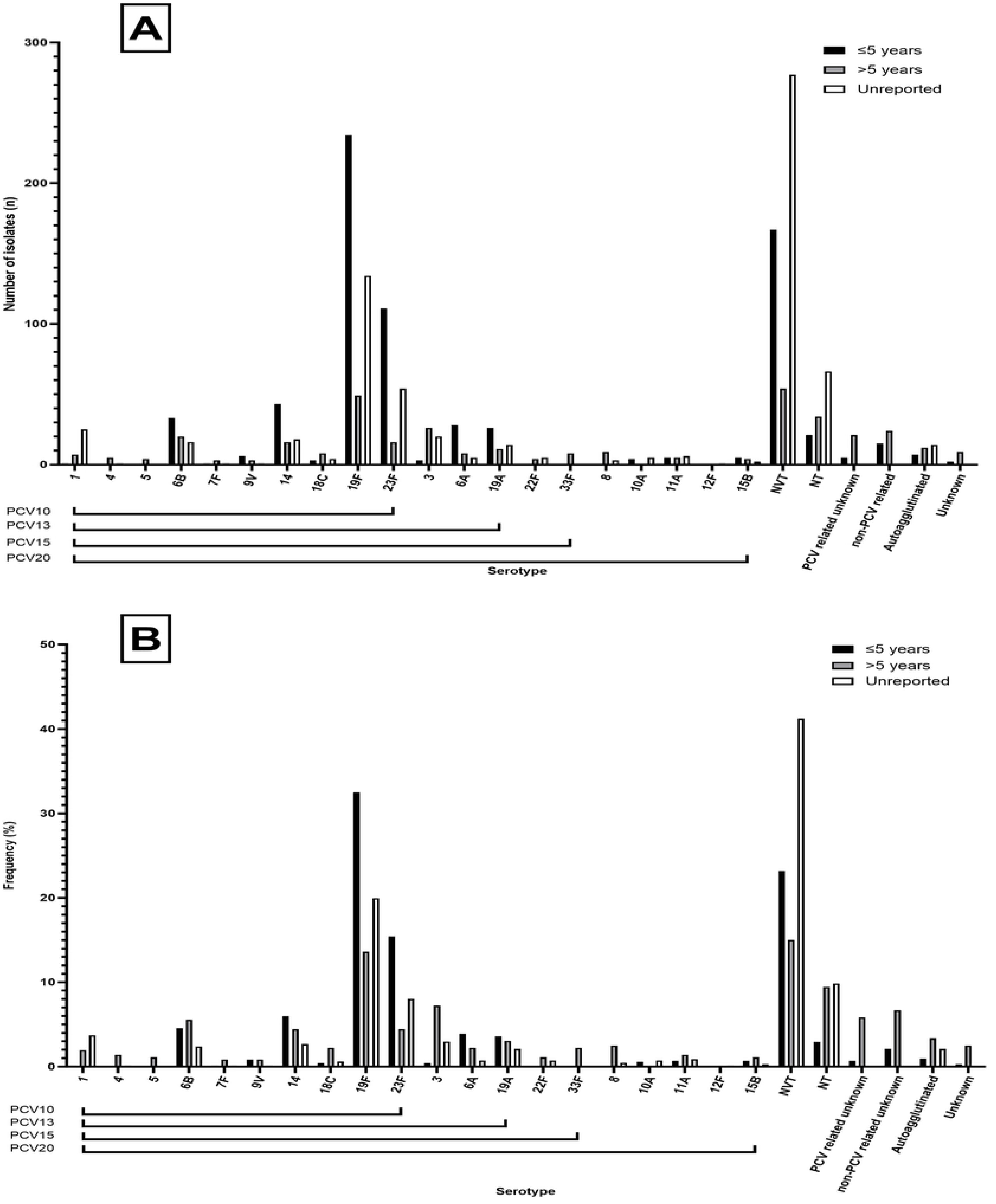
from across ASEAN non-invasive disease studies. (A) Number of isolates for each respective serotype found in PCV10, 13, 15 and 20, including number of NVTs and unknown serotypes from identified studies on non-IPD. (B) Frequency of isolates for each respective serotype found in PCV10, 13, 15 and 20, including number of NVTs and unknown serotypes from identified studies.

**Fig4. Serotypes.**
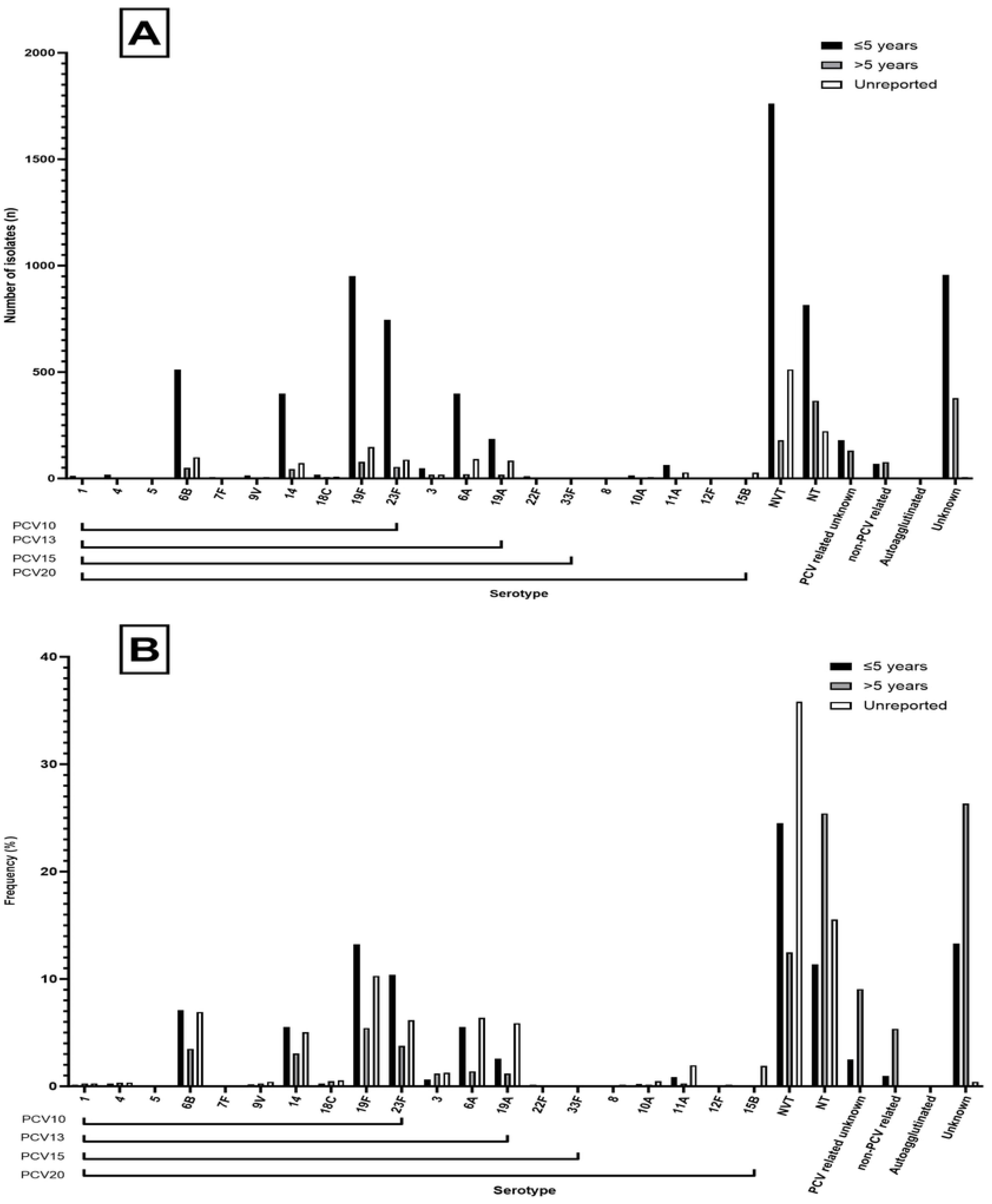
from across ASEAN carriage studies. (A) Number of isolates for each respective serotype found in PCV10, 13, 15 and 20, including number of NVTs and unknown serotypes from identified studies on carriage. (B) Frequency of isolates for each respective serotype found in PCV10, 13, 15 and 20, including number of NVTs and unknown serotypes from identified studies.

**Fig5. Serotypes.**
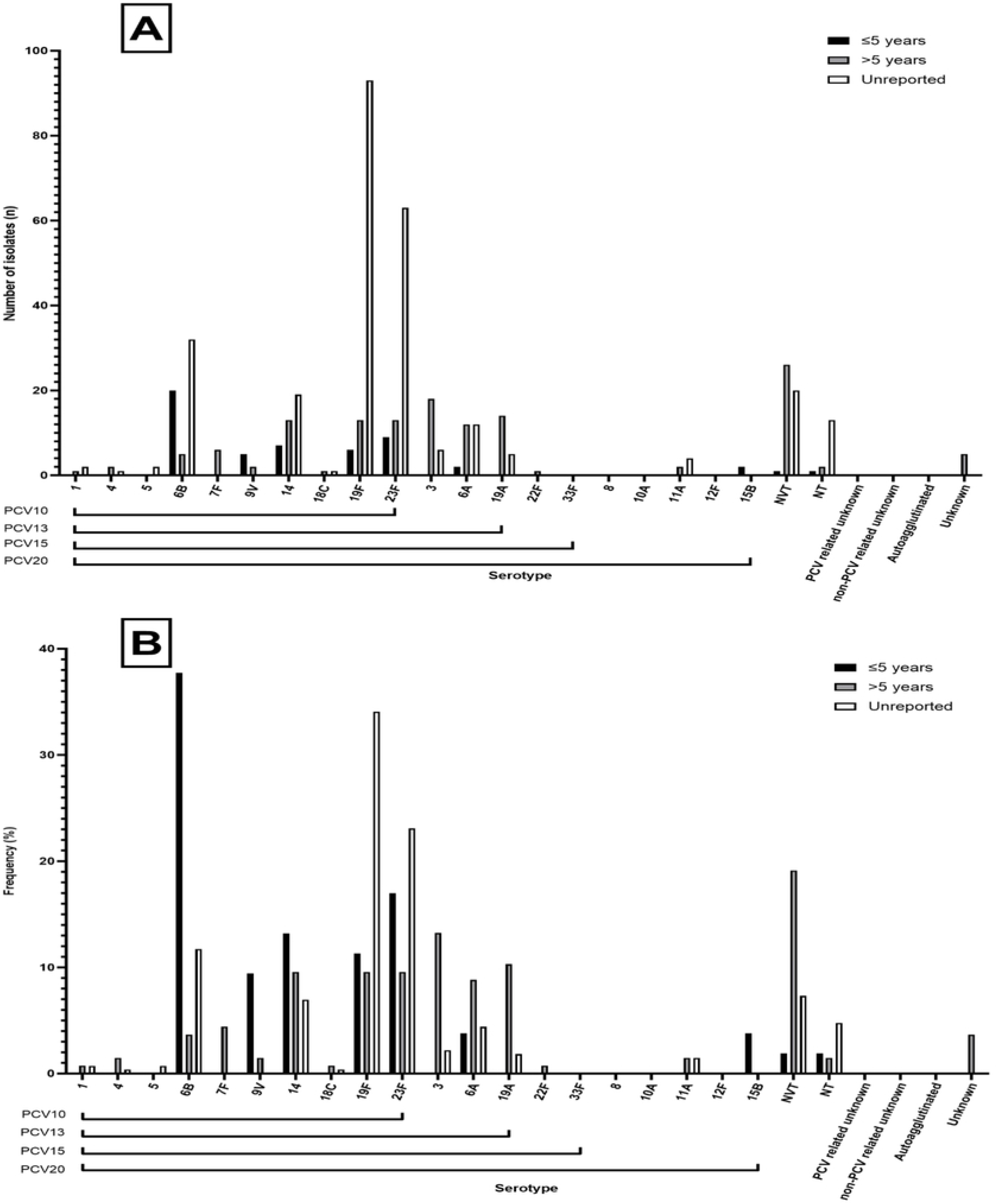
from across ASEAN studies where source could not be determined. (A) Number of isolates for each respective serotype found in PCV10, 13, 15 and 20, including number of NVTs and unknown serotypes from identified studies for which source could not be identified. (B) Frequency of isolates for each respective serotype found in PCV10, 13, 15 and 20, including number of NVTs and unknown serotypes from identified studies.

### Serogroup and serotype prevalence among disease and carriage across ASEAN

The frequency of serogroups and individual serotypes has been calculated for IPD, non-IPD and carriage, separately. The three most common serogroups in all types of conditions were found to be consistently 6, 19 and 23 (Table 3). The rank order of serotypes was found to be not as consistent; serotypes 1 (39.87%), 3 (28.39%), and 4 (21.09%) were identified as the most prevalent in causing IPDs, while serotypes 3 (34.51%), 34 (25.35%), and 1 (23.24%) were commonly observed in non-IPDs. On the other hand, serotypes 34 (53.65%), 3 (21.35%), and 4 (7.29%) were frequently found in carriage (Table 4).

**Table 3.** Rank order of serogroups across IPD, non-IPD and carriage sources in ASEAN.

**Table 4.** Rank order of serotypes across IPD, non-IPD and carriage sources in ASEAN.

### Brunei

No studies were identified for Brunei.

### Cambodia

A total of n = 1,153 isolates were identified with n = 270 (23.42% [95%CI: 21.00 – 25.97]) from IPD, n = 140 isolates (12.14%, [95%CI: 10.31 – 14.17]) from non-IPD and n = 743 isolates from carriage (64.44%, [95%CI: 61.60 – 67.21]). The most common vaccine types across all sources were 19F (n = 143, 12.4% [95%CI: 10.55 – 14.44]), followed by 23F (n = 108, 9.37% [95%CI: 7.75 – 11.20]) and 6B (n = 106, 9.19% [95%CI: 7.59 –11.01]) (Table 5). The three most common NVTs were 6AB (n = 96, 8.33% [95%CI: 6.80 – 10.07]), 34 (n = 35, 3.04% [95%CI: 2.12 – 4.20]) and 35C (n = 19, 1.65% [95%CI: 0.99 – 2.56]). PCV10 and PCV13, PCV15 and PCV20 serotypes accounted for 41.28% [95%CI: 38.42 – 44.19], 58.20% [95%CI: 55.29 – 61.06], 58.37% [95%CI: 55.46 – 61.23] and 64.44% [95%CI: 61.60 – 67.21] of all isolates respectively. Non-typeables accounted for n = 88 (7.63%, [95%CI: 6.17 – 9.32]). No unknown serotypes were reported. Fig6, Fig7, Fig8 show the split of serotype counts from across IPD, non-IPD and carriage sources.

**Table 5.** Serotype counts and percentages for all countries in Cambodia, split by age category and invasive, non-invasive and carriage sources.

**Fig6. Serotypes.**
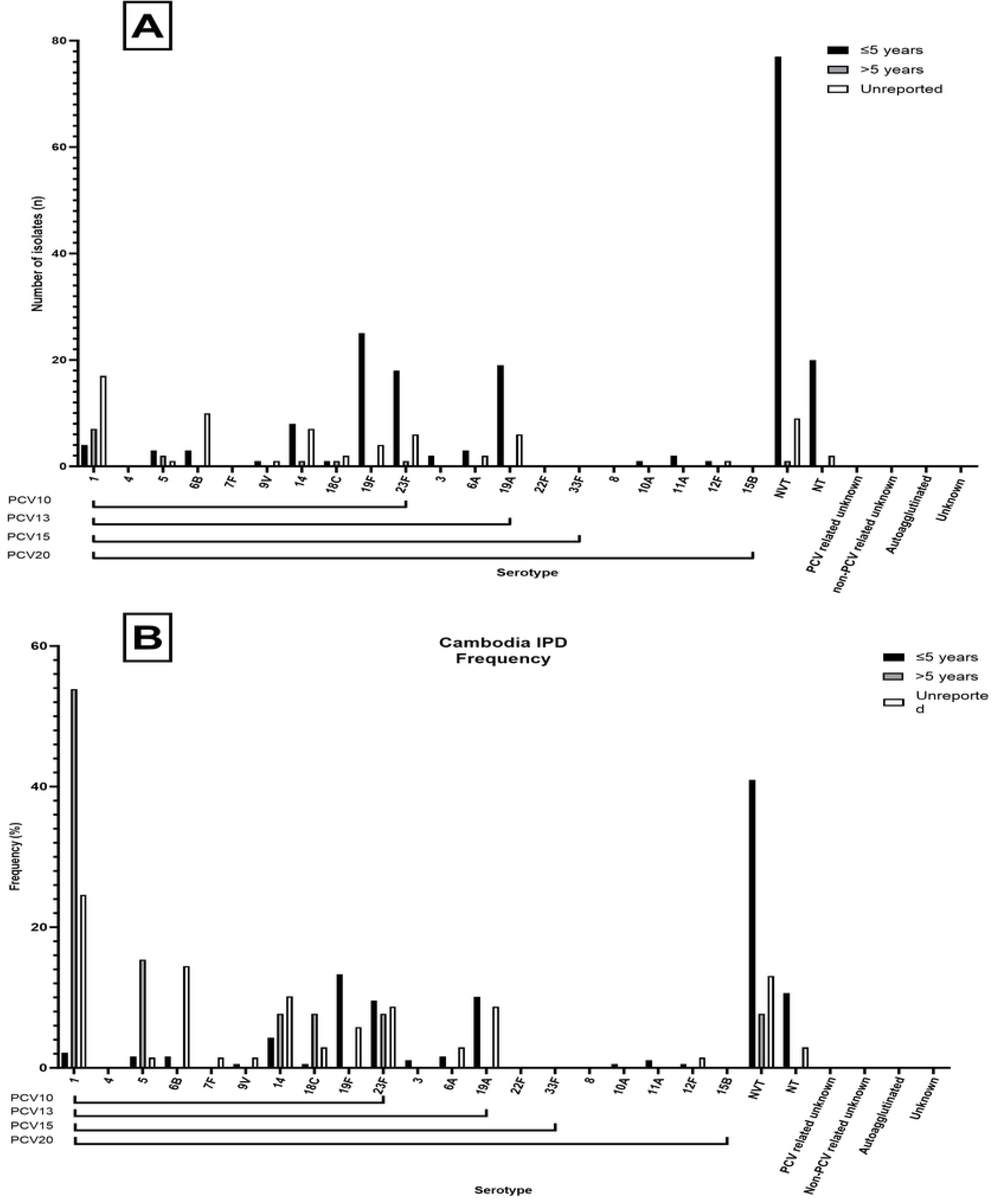
from across Cambodian invasive disease studies. (A) Number of isolates for each respective serotype found in PCV10, 13, 15 and 20, including number of NVTs and unknown serotypes from identified studies for which source could not be identified for Cambodia invasive disease. (B) Frequency of isolates for each respective serotype found in PCV10, 13, 15 and 20, including number of NVTs and unknown serotypes from identified studies.

**Fig7. Serotypes.**
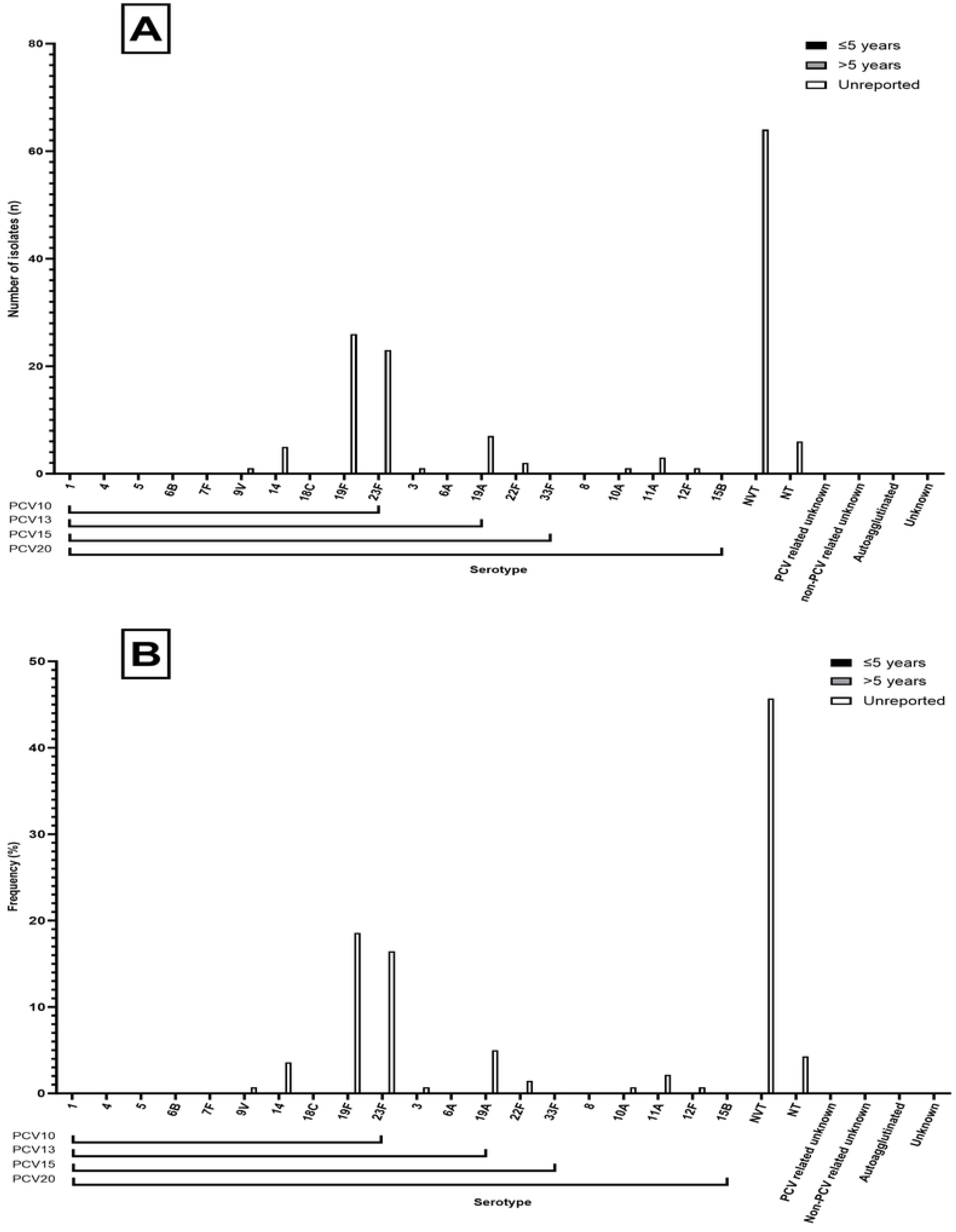
from across Cambodian non-invasive disease studies. (A) Number of isolates for each respective serotype found in PCV10, 13, 15 and 20, including number of NVTs and unknown serotypes from identified studies for which source could not be identified for Cambodia non-invasive disease. (B) Frequency of isolates for each respective serotype found in PCV10, 13, 15 and 20, including number of NVTs and unknown serotypes from identified studies.

**Fig8. Serotypes.**
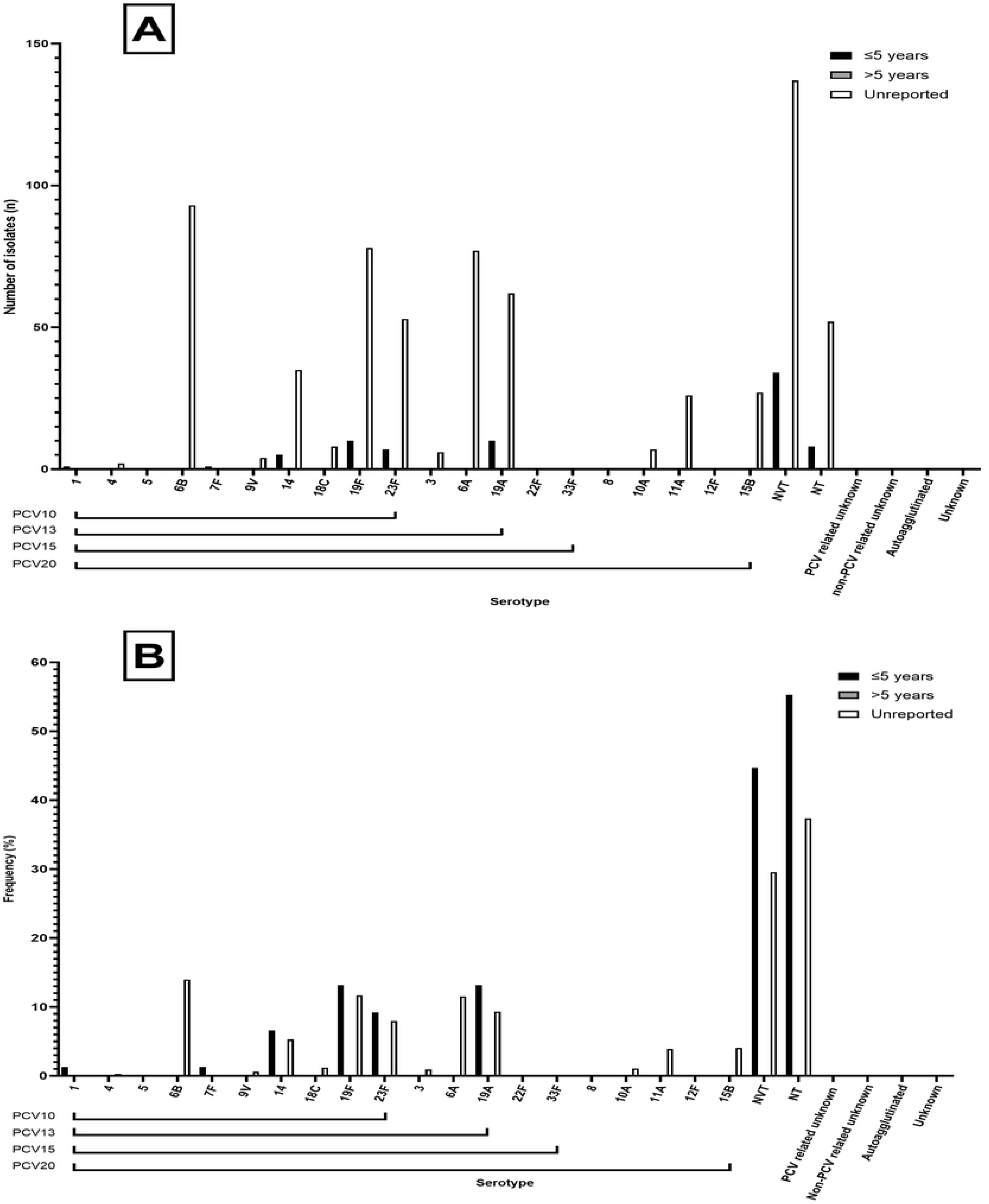
from across Cambodian carriage studies. (A) Number of isolates for each respective serotype found in PCV10, 13, 15 and 20, including number of NVTs and unknown serotypes from identified studies for which source could not be identified for Cambodia carriage. (B) Frequency of isolates for each respective serotype found in PCV10, 13, 15 and 20, including number of NVTs and unknown serotypes from identified studies.

### Indonesia

A total of n = 2,012 isolates were identified. N = 4 (0.20% [95%CI: 0.05 – 0.51]) were from IPD, n = 65 isolates (3.23%, [95%CI: 2.50 – 4.10]) from non-IPD and n = 1,943 isolates from carriage (96.57%, [95%CI: 95.68 – 97.32]). The most common vaccine types across all sources were 19F (n = 175, 8.70% [95%CI: 7.50 – 10.01]), followed by 23F (n = 144, 7.16% [95%CI: 6.07 – 8.37]) and 14 (n = 76, 3.78% [95%CI: 2.99 – 4.71]) (Table 6). The three most common NVTs were 6AB (n = 259, 12.87% [95%CI: 11.44 – 14.42]), 15BC (n = 134, 6.66% [95%CI: 5.61 – 7.84]) and 23A (n = 81, 4.03% [95%CI: 3.21 – 4.98]). PCV10 and PCV13, PCV15 and PCV20 serotypes accounted for 24.45% [95%CI: 22.59 – 26.39], 34.59% [95%CI: 32.51 – 36.72], 35.19% [95%CI: 33.10 – 37.32] and 38.97% [95%CI: 36.83 – 41.14] of all isolates respectively. Non-typeables accounted for n = 312 (15.51%, [95%CI: 13.95 – 17.16]) and unknown serotypes accounted for n = 24 (1.19%, [95%CI: 0.77 – 1.77]) of total isolates. Fig9, Fig10, Fig11 show the split of serotype counts from across IPD, non-IPD and carriage sources.

**Table 6.** Serotype counts and percentages for all countries in Indonesia, split by age category and invasive, non-invasive and carriage sources.

**Fig9. Serotypes.**
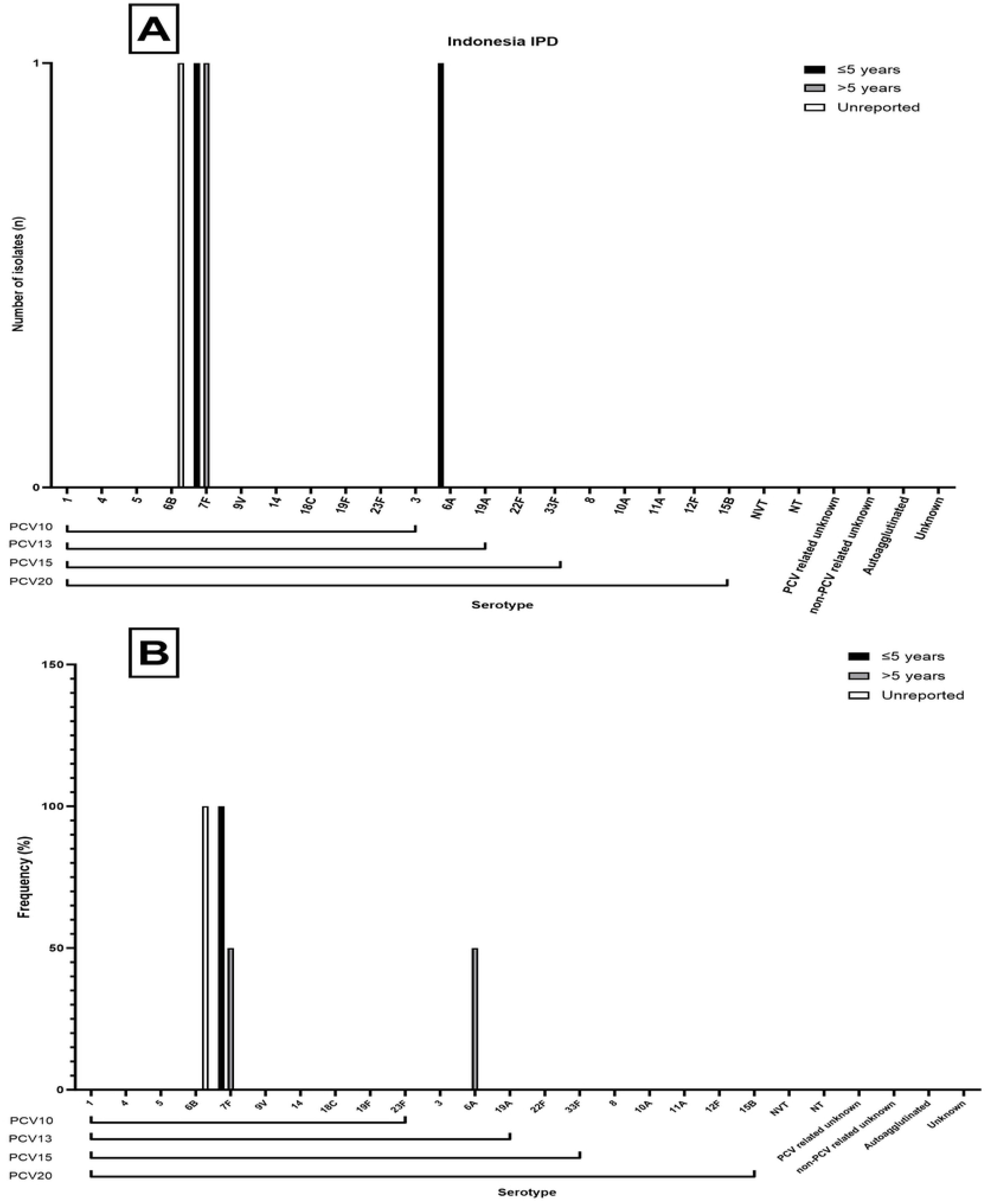
from across Indonesian invasive disease studies. (A) Number of isolates for each respective serotype found in PCV10, 13, 15 and 20, including number of NVTs and unknown serotypes from identified studies for which source could not be identified for Indonesia invasive disease. (B) Frequency of isolates for each respective serotype found in PCV10, 13, 15 and 20, including number of NVTs and unknown serotypes from identified studies.

**Fig10. Serotypes.**
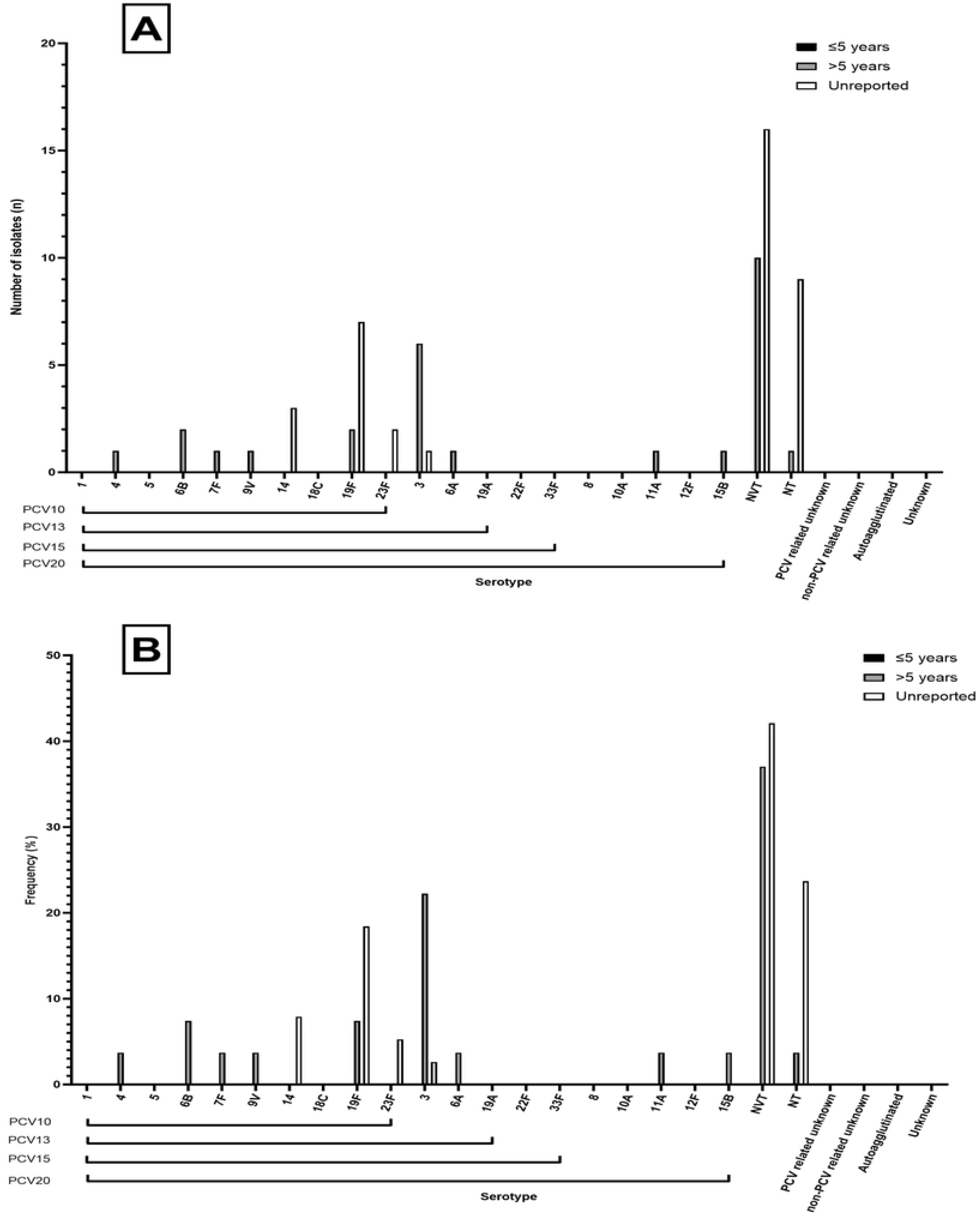
from across Indonesian non-invasive disease studies. (A) Number of isolates for each respective serotype found in PCV10, 13, 15 and 20, including number of NVTs and unknown serotypes from identified studies for which source could not be identified for Indonesia non-invasive disease. (B) Frequency of isolates for each respective serotype found in PCV10, 13, 15 and 20, including number of NVTs and unknown serotypes from identified studies.

**Fig11. Serotypes.**
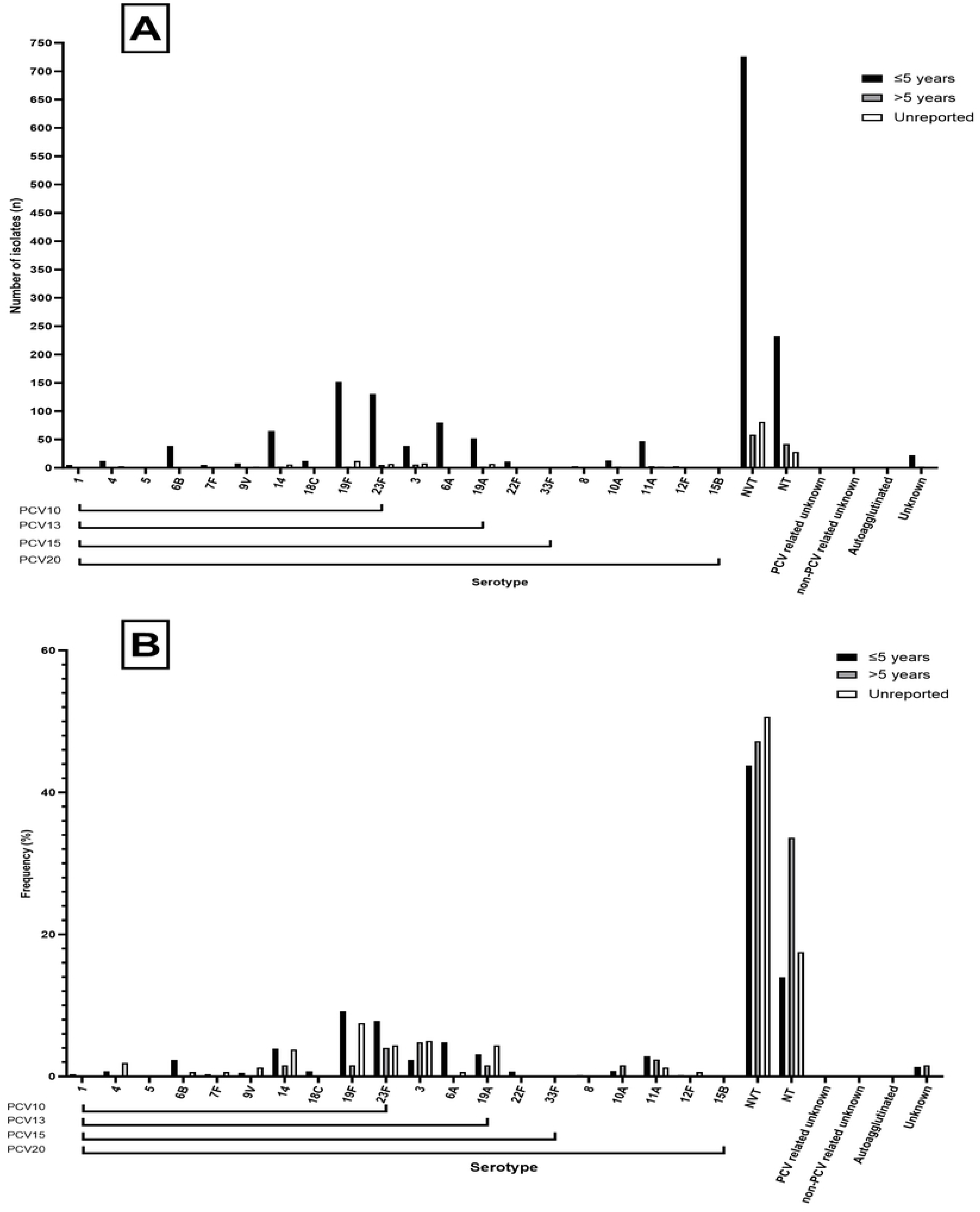
from across Indonesian carriage studies. (A) Number of isolates for each respective serotype found in PCV10, 13, 15 and 20, including number of NVTs and unknown serotypes from identified studies for which source could not be identified for Indonesia carriage. (B) Frequency of isolates for each respective serotype found in PCV10, 13, 15 and 20, including number of NVTs and unknown serotypes from identified studies.

### Laos

A total of n = 217 isolates were identified. N = 33 (15.21%, [95%CI: 10.71 – 20.69]), n = 0 from non-IPD and n = 184 isolates from carriage (84.79%, [95%CI: 79.31 – 89.29]). The most common vaccine types across all sources were 6B (n =49, 22.58% [95%CI: 17/20 – 28.73]), followed by 23F (n = 37, 17.05% [95%CI: 12.30 - 22.73]) and 19F (n = 34, 15.67% [95%CI: 11.10 – 21.20]) (Table 7). The three most common NVTs were 15BC (n = 6, 2.76%, [95%CI: 1.02 – 5.92]), 6ABC (n = 3, 1.38% [95%CI: 0.29 – 3.99]), with the remaining serotypes 15C, 23B and 38 each having n = 1 (0.46%, [95%CI: 0.01 – 2.54]) each. PCV10, PCV13, PCV15 and PCV20 serotypes accounted for 63.59% [95%CI: 56.81 – 70.00], 69.12% [95%CI: 62.52 – 75.20], 69.12% [95%CI: 62.52 – 75.20] and 69.12% [95%CI: 62.52 – 75.20] of all isolates respectively. Non-typeables accounted for n = 55 (25.35%, [95%CI: 19.70 – 31.68]) and unknown serotypes accounted for zero of total isolates. Fig12, Fig13, Fig14 show the split of serotype counts from across IPD, non-IPD and carriage sources.

**Table 7.** Serotype counts and percentages for all countries in Laos, split by age category and invasive, non-invasive and carriage sources.

**Fig12. Serotypes.**
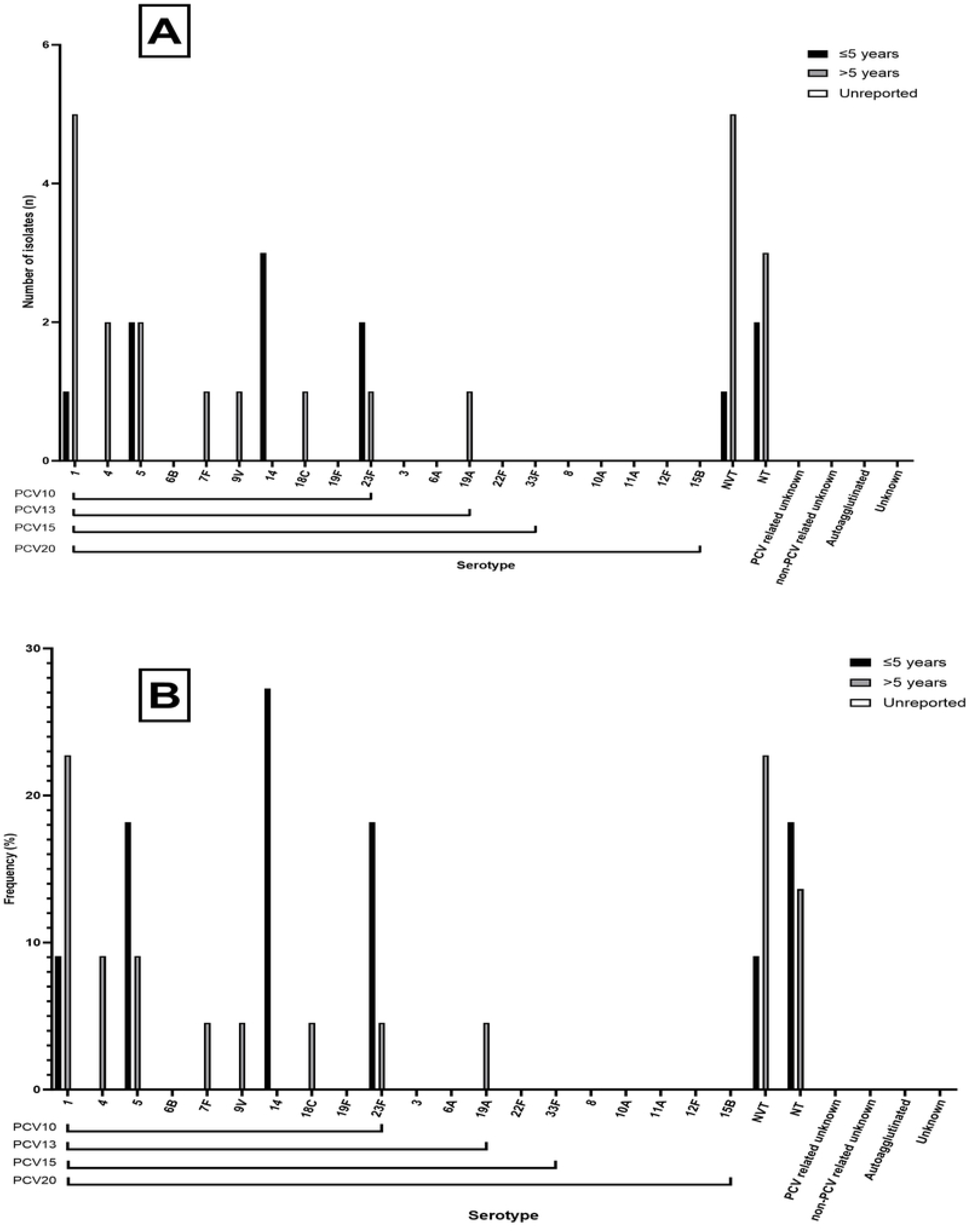
from across Laos invasive disease studies. (A) Number of isolates for each respective serotype found in PCV10, 13, 15 and 20, including number of NVTs and unknown serotypes from identified studies for which source could not be identified for Laos invasive disease. (B) Frequency of isolates for each respective serotype found in PCV10, 13, 15 and 20, including number of NVTs and unknown serotypes from identified studies.

**Fig13. Serotypes.**
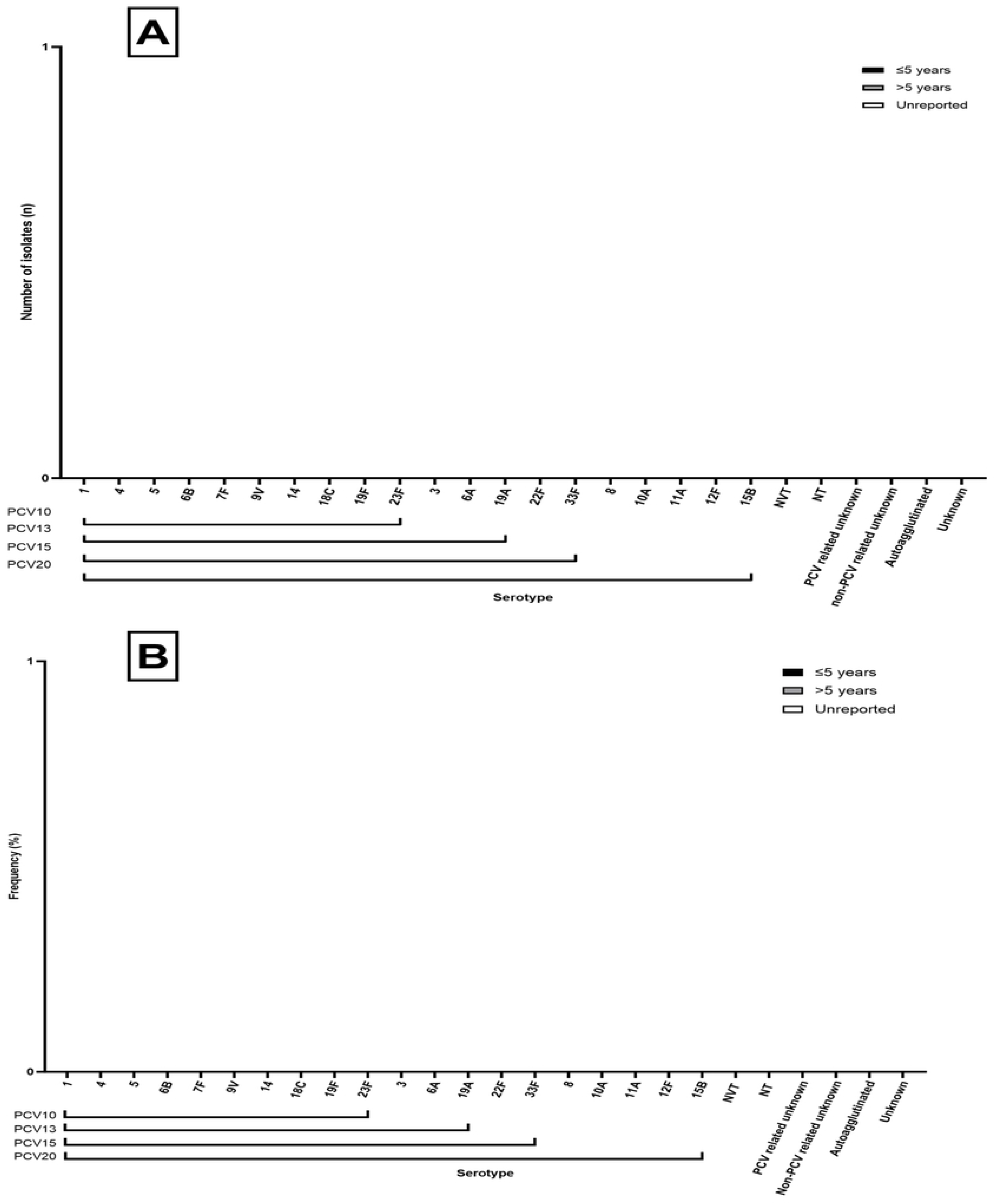
from across Laos non-invasive disease studies. (A) Number of isolates for each respective serotype found in PCV10, 13, 15 and 20, including number of NVTs and unknown serotypes from identified studies for which source could not be identified for Laos non-invasive disease. (B) Frequency of isolates for each respective serotype found in PCV10, 13, 15 and 20, including number of NVTs and unknown serotypes from identified studies. As seen, no serotypes were identified.

**Fig14. Serotypes.**
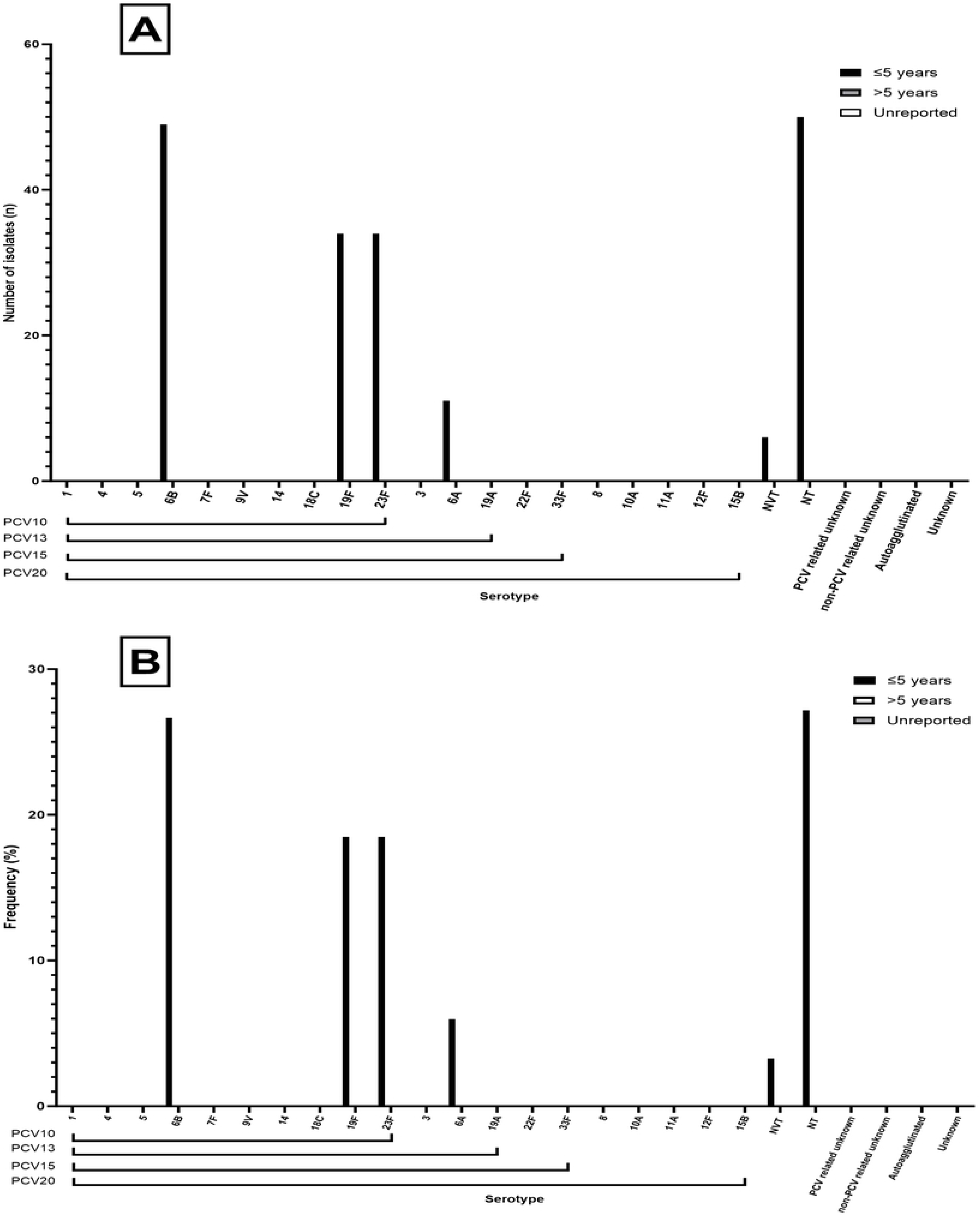
from across Laos carriage studies (A) Number of isolates for each respective serotype found in PCV10, 13, 15 and 20, including number of NVTs and unknown serotypes from identified studies for which source could not be identified for Laos carriage. (B) Frequency of isolates for each respective serotype found in PCV10, 13, 15 and 20, including number of NVTs and unknown serotypes from identified studies.

### Malaysia

A total of n = 2,210 isolates were identified. N = 883 (39.95% [95%CI: 37.90 – 42.03]) were from IPD, n = 778 isolates (35.20%, [95%CI: 33.31 – 37.24]) from non-IPD, n = 426 isolates from carriage (19.28%, [95%CI: 17.65 – 20.98]) and n = 123 (5.57% [95%CI: 4.65 – 6.60]) were from unknown source. The most common vaccine types across all sources were 19F (n = 335, 15.16% [95%CI: 13.69 – 16.72]), followed by 14 (n = 184, 8.33% [95%CI: 7.21 – 9.56]) and 23F (n = 172, 7.78% [95%CI: 6.70 – 8.98]) (Table 8). The three most common NVTs were 6AB (n= 87, 3.94%, [95%CI: 3.16 – 4.83]), 23A (n = 37, 1.67% [95%CI: 1.18 – 2.30]) and 6C (n = 26, 1.18% [95%CI: 0.77 – 1.72]). PCV10 and PCV13, PCV15 and PCV20 serotypes accounted for 47.01% [95%CI: 44.91 – 49.12], 62.40% [95%CI: 60.34 – 64.42], 62.94% [95%CI: 60.89 – 64.96] and 65.16% [95%CI: 63.13 – 67.15] of all isolates respectively. Non-typeables accounted for n = 154 (6.97%, [95%CI: 5.94 – 8.11]) of all isolates and unknown serotypes accounted for n = 34 (1.54% [95%CI: 1.07 – 2.14]) of total isolates. Fig15, Fig16, Fig17, Fig18 show the split of serotype counts from across IPD, non-IPD, carriage and unknown sources.

**Table 8.** Serotype counts and percentages for all countries in Malaysia, split by age category and invasive, non-invasive, carriage sources and from sources which could not be determined.

**Fig15. Serotypes.**
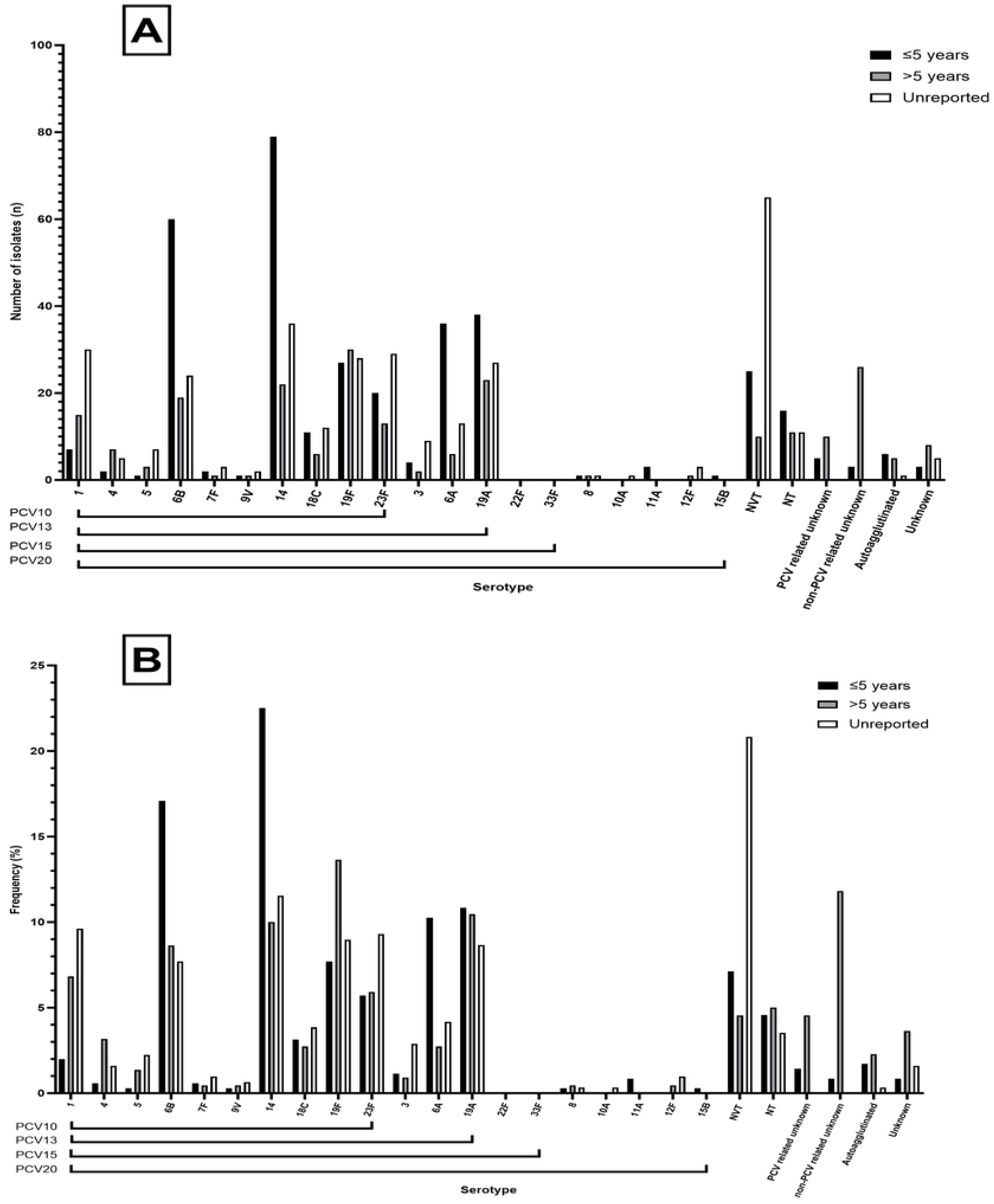
from across Malaysian invasive disease studies. (A) Number of isolates for each respective serotype found in PCV10, 13, 15 and 20, including number of NVTs and unknown serotypes from identified studies for which source could not be identified for Malaysia invasive disease. (B) Frequency of isolates for each respective serotype found in PCV10, 13, 15 and 20, including number of NVTs and unknown serotypes from identified studies.

**Fig16. Serotypes.**
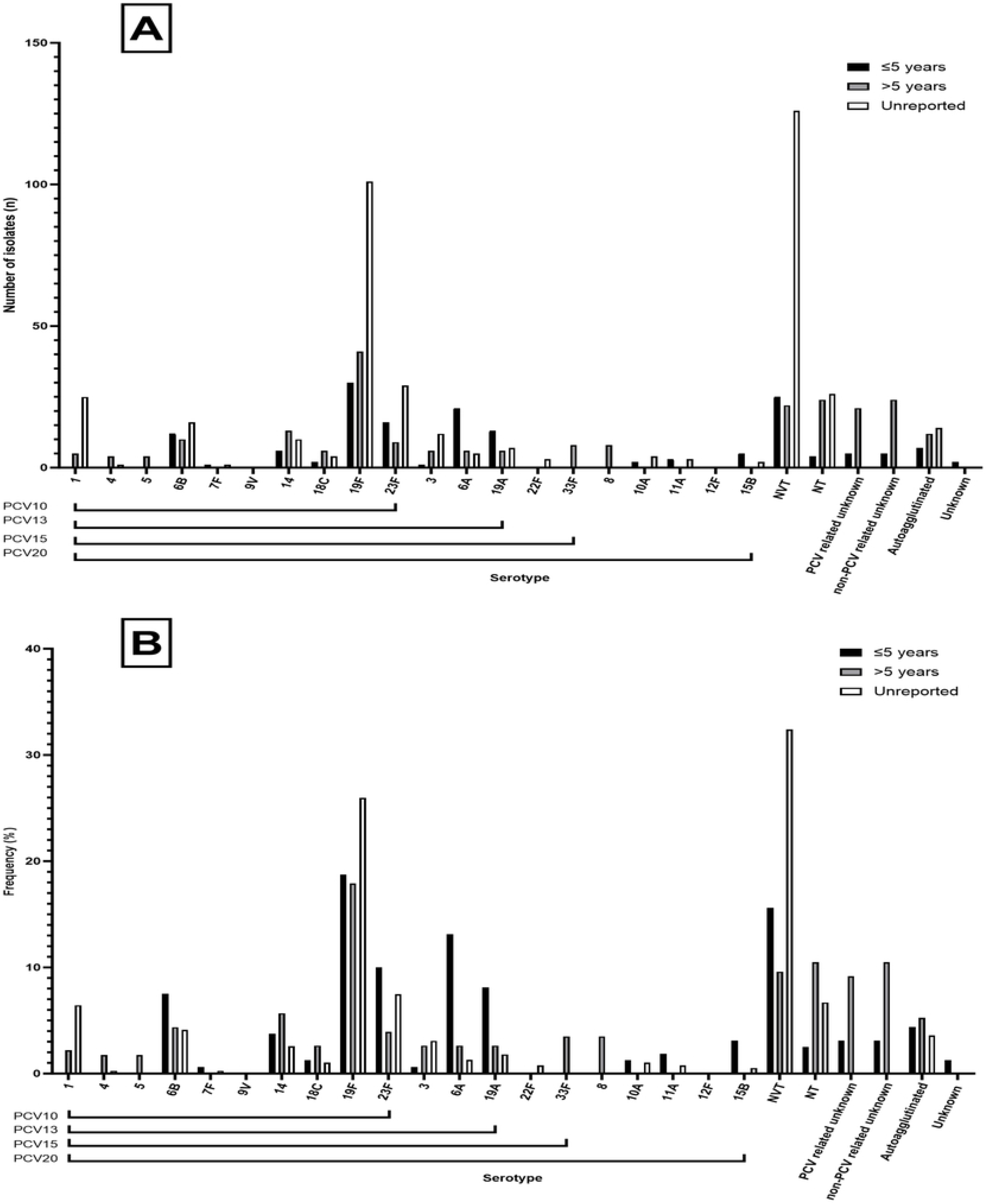
from across Malaysian non-invasive disease studies. (A) Number of isolates for each respective serotype found in PCV10, 13, 15 and 20, including number of NVTs and unknown serotypes from identified studies for which source could not be identified for Malaysia non-invasive disease. (B) Frequency of isolates for each respective serotype found in PCV10, 13, 15 and 20, including number of NVTs and unknown serotypes from identified studies.

**Fig17. Serotypes.**
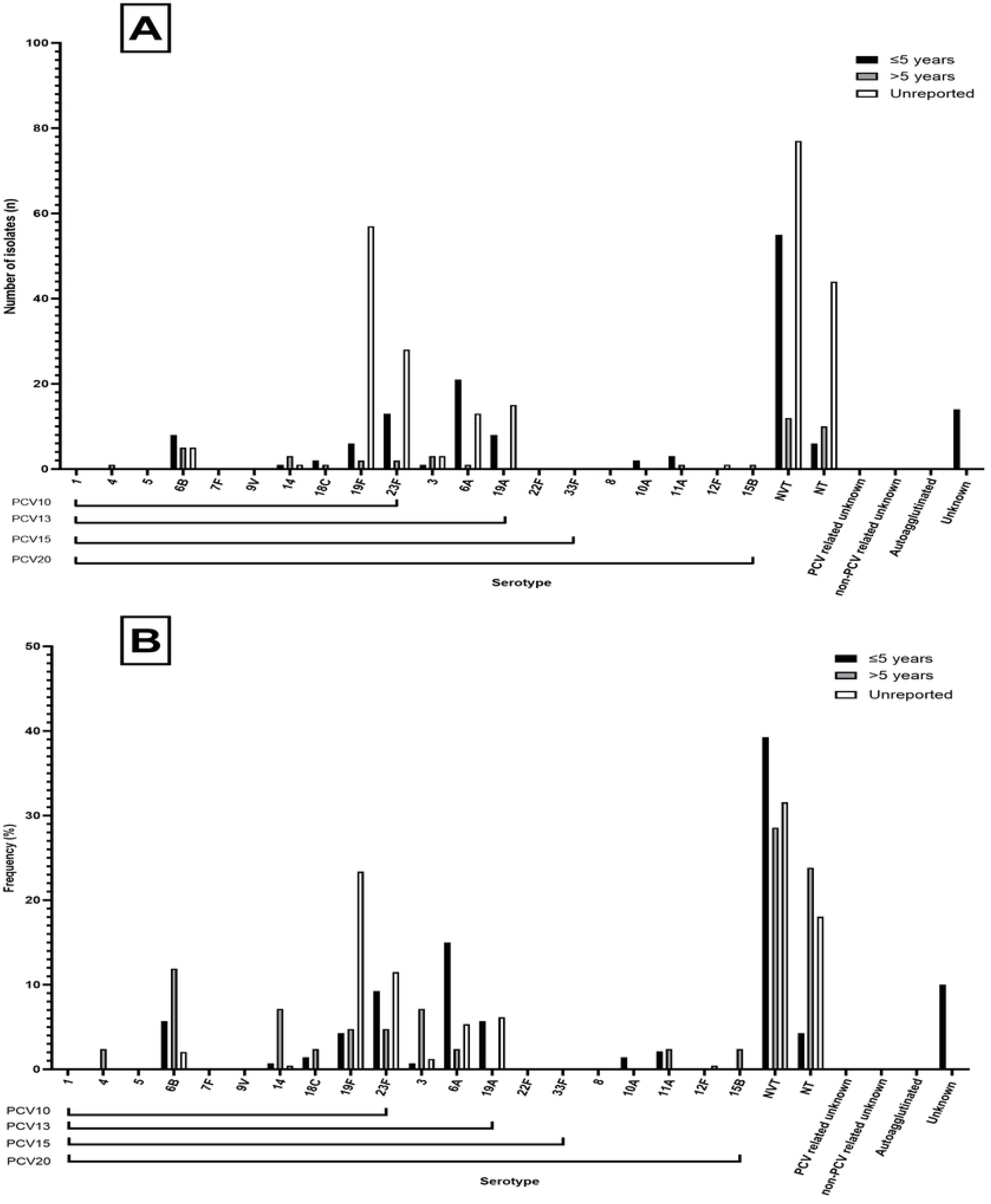
from across Malaysian carriage studies (A) Number of isolates for each respective serotype found in PCV10, 13, 15 and 20, including number of NVTs and unknown serotypes from identified studies for which source could not be identified for Malaysia carriage. (B) Frequency of isolates for each respective serotype found in PCV10, 13, 15 and 20, including number of NVTs and unknown serotypes from identified studies.

**Fig18. Serotypes.**
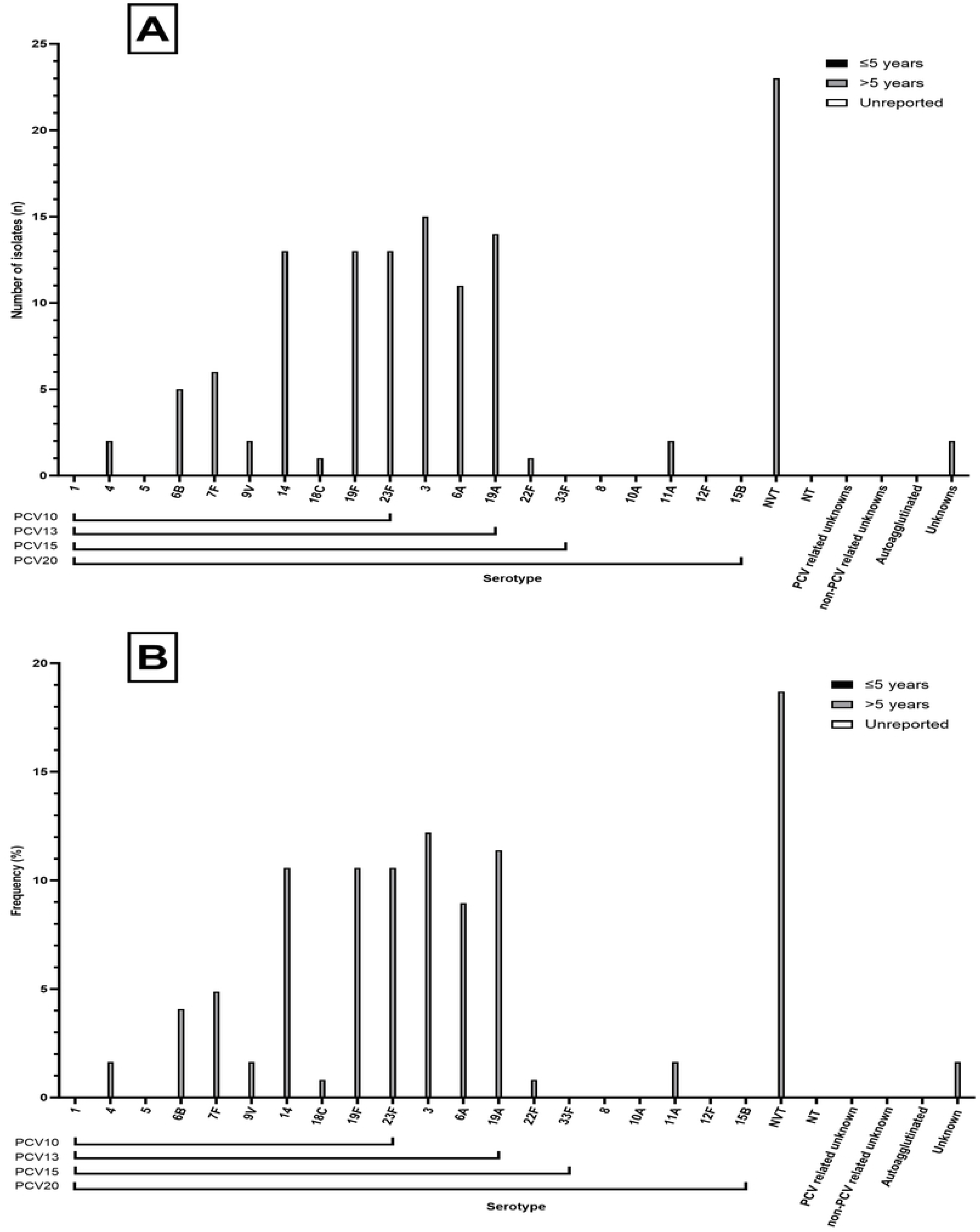
from across Malaysian studies where source could not be determined. (A) Number of isolates for each respective serotype found in PCV10, 13, 15 and 20, including number of NVTs and unknown serotypes from identified studies for which source could not be identified for Malaysia unknown sources. (B) Frequency of isolates for each respective serotype found in PCV10, 13, 15 and 20, including number of NVTs and unknown serotypes from identified studies.

### Myanmar

A total of n = 4,420 isolates were identified. Zero isolates were from IPD, n = 24 isolates (0.54%, [95%CI: 0.35 – 0.81]) from non-IPD and n = 4,396 isolates from carriage (99.46%, [95%CI: 99.19 – 99.65]). The most common vaccine types across all sources were 19F (n = 606, 13.71% [95%CI: 12.71 – 14.76]), followed by 23F (n = 473, 10.70% [95%CI: 9.80 – 11.65]) and 6B (n = 337, 7.62% [95%CI: 6.86 – 8.45]) (Table 9). The three most common NVTs were 15BC (n = 171, 3.87% [95%CI: 3.32 – 4.48]), 34 (n = 139, 3.14% [95%CI: 2.65 – 3.70]), 6C (n = 117, 2.65% [95%CI: 2.19 – 3.16]). PCV10, PCV13, PCV15 and PCV20 serotypes accounted for 36.92% [95%CI: 35.50 – 38.37], 43.67% [95%CI: 42.20 – 45.14], 43.67% [95%CI: 42.20 – 45.14] and 43.67% [95%CI: 42.20 – 45.14] of all isolates respectively. Non-typeables accounted for n = 665 (15.05% [95%CI: 14.00 – 16.13]) of total isolates and unknown serotypes accounted for n = 1148, 25.97% [95%CI: 24.69 – 27.29] of total isolates. Fig19, Fig20, Fig21 show the split of serotype counts from across IPD, non-IPD and carriage sources.

**Table 9.** Serotype counts and percentages for all countries in Myanmar, split by age category and invasive, non-invasive and carriage sources.

**Fig19. Serotypes.**
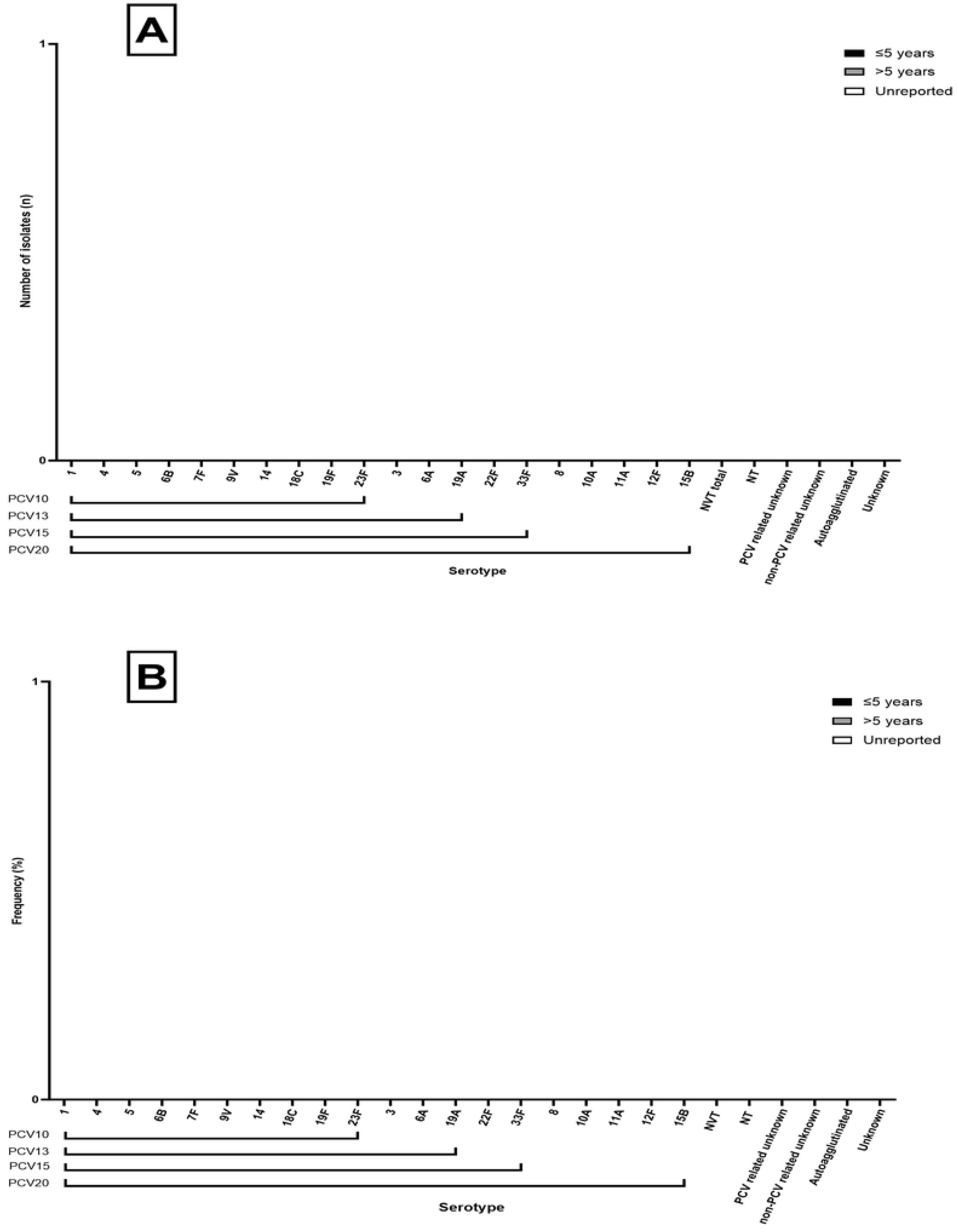
from across Myanmar invasive disease studies. (A) Number of isolates for each respective serotype found in PCV10, 13, 15 and 20, including number of NVTs and unknown serotypes from identified studies for which source could not be identified for Myanmar invasive disease. (B) Frequency of isolates for each respective serotype found in PCV10, 13, 15 and 20, including number of NVTs and unknown serotypes from identified studies.

**Fig20. Serotypes.**
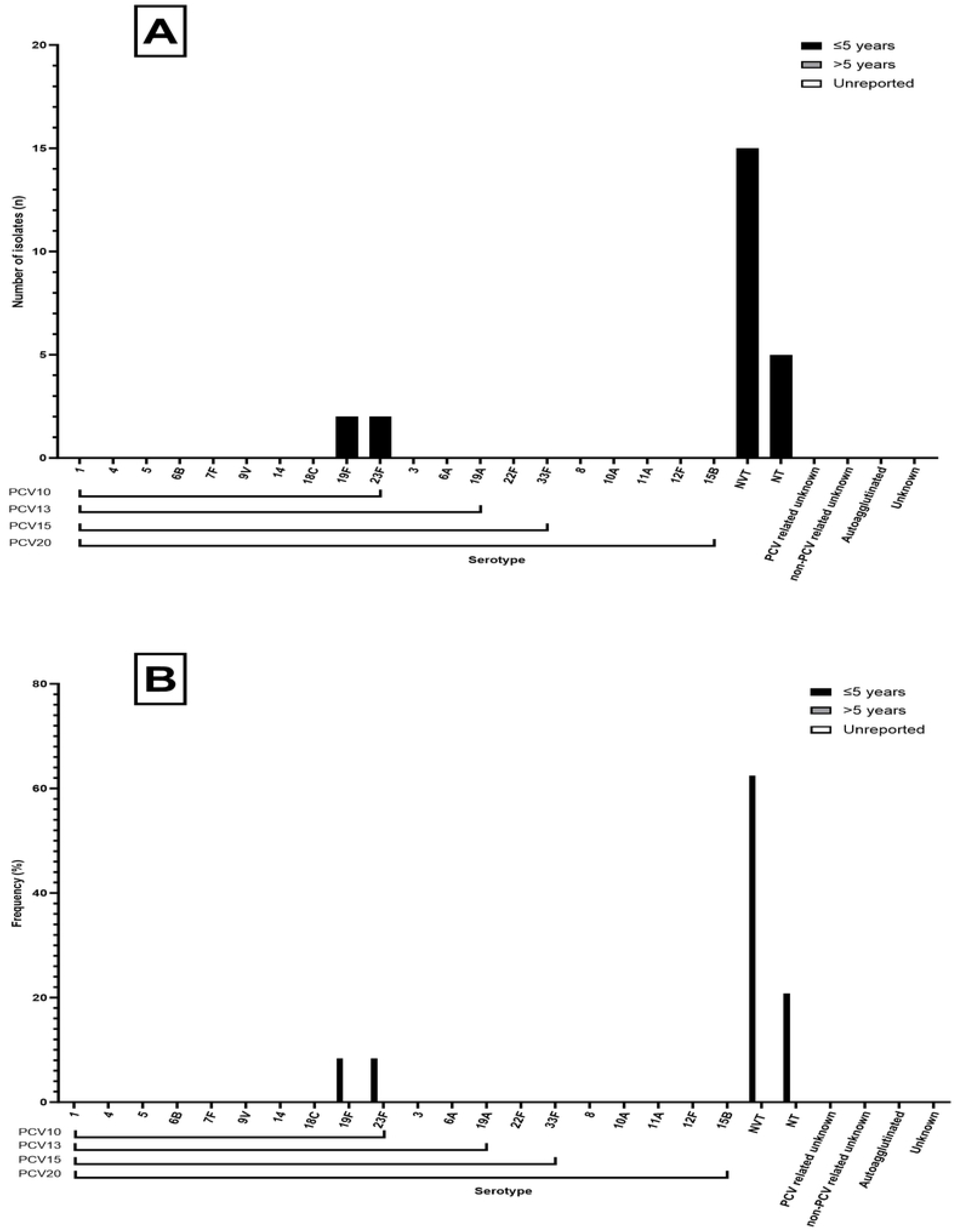
from across Myanmar non-invasive disease studies. (A) Number of isolates for each respective serotype found in PCV10, 13, 15 and 20, including number of NVTs and unknown serotypes from identified studies for which source could not be identified for Myanmar non-invasive disease (B) Frequency of isolates for each respective serotype found in PCV10, 13, 15 and 20, including number of NVTs and unknown serotypes from identified studies.

**Fig21. Serotypes.**
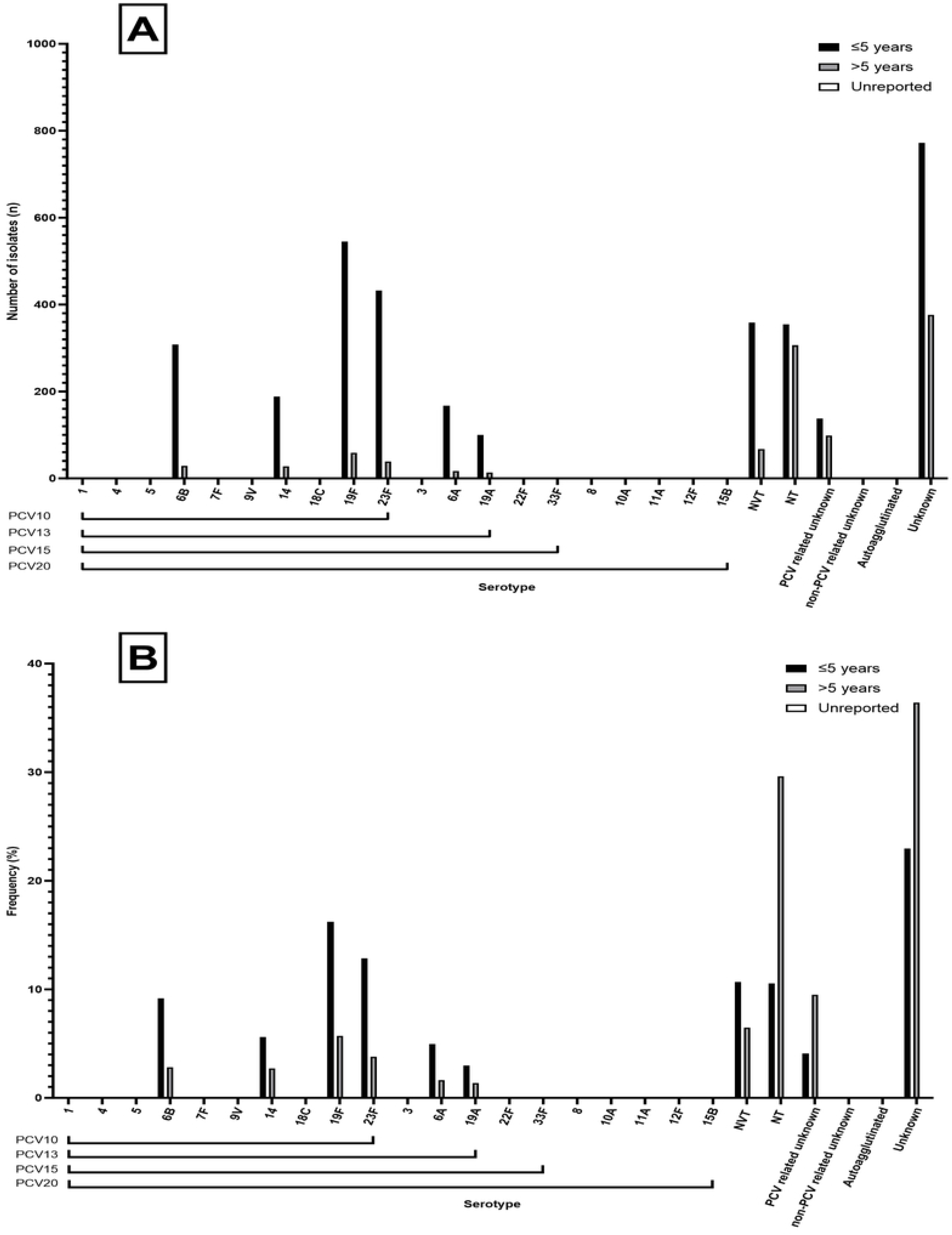
from across Myanmar carriage studies. (A) Number of isolates for each respective serotype found in PCV10, 13, 15 and 20, including number of NVTs and unknown serotypes from identified studies for which source could not be identified for Myanmar carriage (B) Frequency of isolates for each respective serotype found in PCV10, 13, 15 and 20, including number of NVTs and unknown serotypes from identified studies.

### Philippines

A total of n = 587 isolates were identified. N = 365 (62.18% [95%CI: 58.12 – 66.12]) were from IPD, n = 51 isolates (8.69%, [95%CI: 6.54 – 11.27]) from non-IPD, n = 158 isolates from carriage (26.92%, [95%CI: 23.37– 30.70]) and n = 13 (2.10%, [95%CI: 1.18 – 3.76]) were of unknown source. The most common vaccine types across all sources were 1 (n = 57, 9.71% [95%CI: 7.44 – 12.40]), followed by 5 (n = 34, 5.79% [95%CI: 4.04 – 8.00]) and 14 (n = 29, 4.94% [95%CI: 3.33 – 7.02]) (Table 10). The three most common NVTs were 6 (n= 43, 7.33% [95%CI: 5.35 – 9.74]), 23 (n = 23, 3.92% [95%CI: 2.50– 5.82]) and 18 (n = 15, 2.56% [95%CI: 1.44 – 4.18]). PCV10, PCV13, PCV15 and PCV20 serotypes accounted for 32.54% [95%CI: 28.76 – 36.49], 39.52% [95%CI: 35.54 – 43.61], 39.52% [95%CI: 35.54 – 43.61] and 40.55% [95%CI: 36.54 – 44.64] of all isolates respectively. Non-typeables accounted for n = 38 (6.47% [95%CI: 4.62 – 8.78]) and unknown serotypes accounted for n = 84 (14.31% [95%CI: 11.58 – 17.41]) of total isolates. Fig22, Fig23, Fig24, Fig25 show the split of serotype counts from across IPD, non-IPD, carriage and unknown sources.

**Table 10.** Serotype counts and percentages for all countries in Philippines, split by age category and invasive, non-invasive, carriage sources and from sources which could not be determined.

**Fig22.**
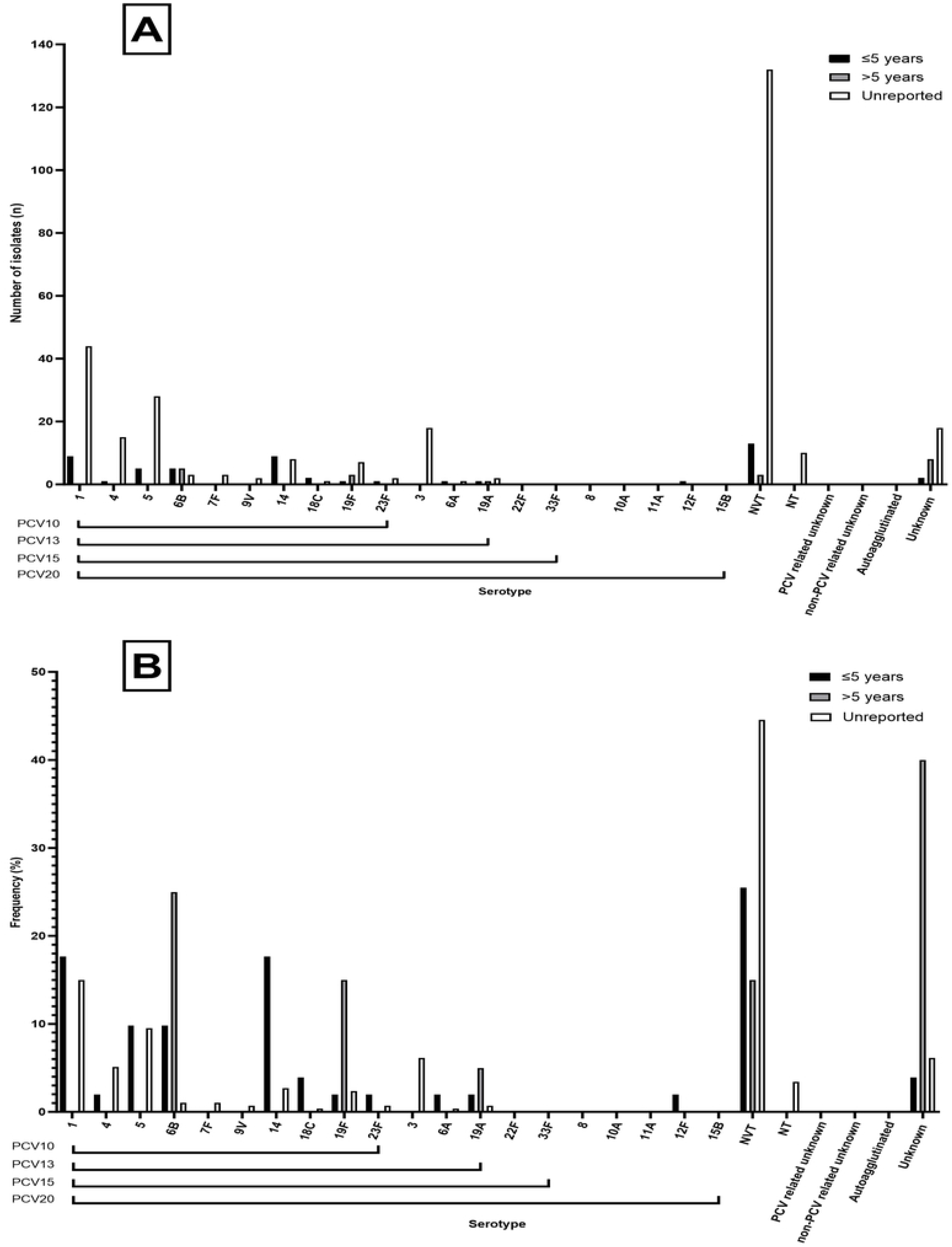
Serotypes from across Philippines invasive disease studies. (A) Number of isolates for each respective serotype found in PCV10, 13, 15 and 20, including number of NVTs and unknown serotypes from identified studies for which source could not be identified for Philippines invasive disease. (B) Frequency of isolates for each respective serotype found in PCV10, 13, 15 and 20, including number of NVTs and unknown serotypes from identified studies.

**Fig23.**
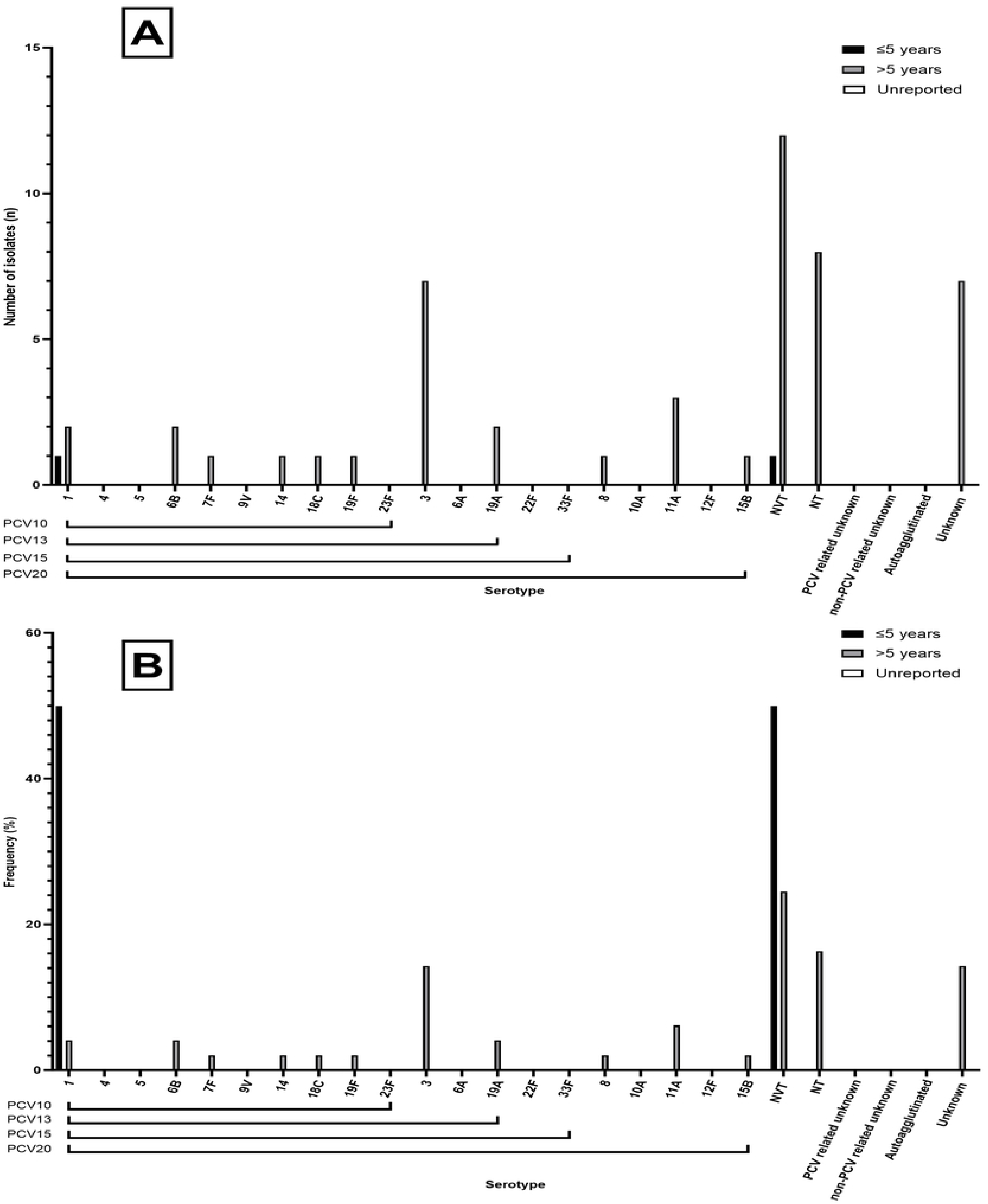
Serotypes from across Philippines non-invasive disease studies. (A) Number of isolates for each respective serotype found in PCV10, 13, 15 and 20, including number of NVTs and unknown serotypes from identified studies for which source could not be identified for Philippines non-invasive disease. (B) Frequency of isolates for each respective serotype found in PCV10, 13, 15 and 20, including number of NVTs and unknown serotypes from identified studies.

**Fig24.**
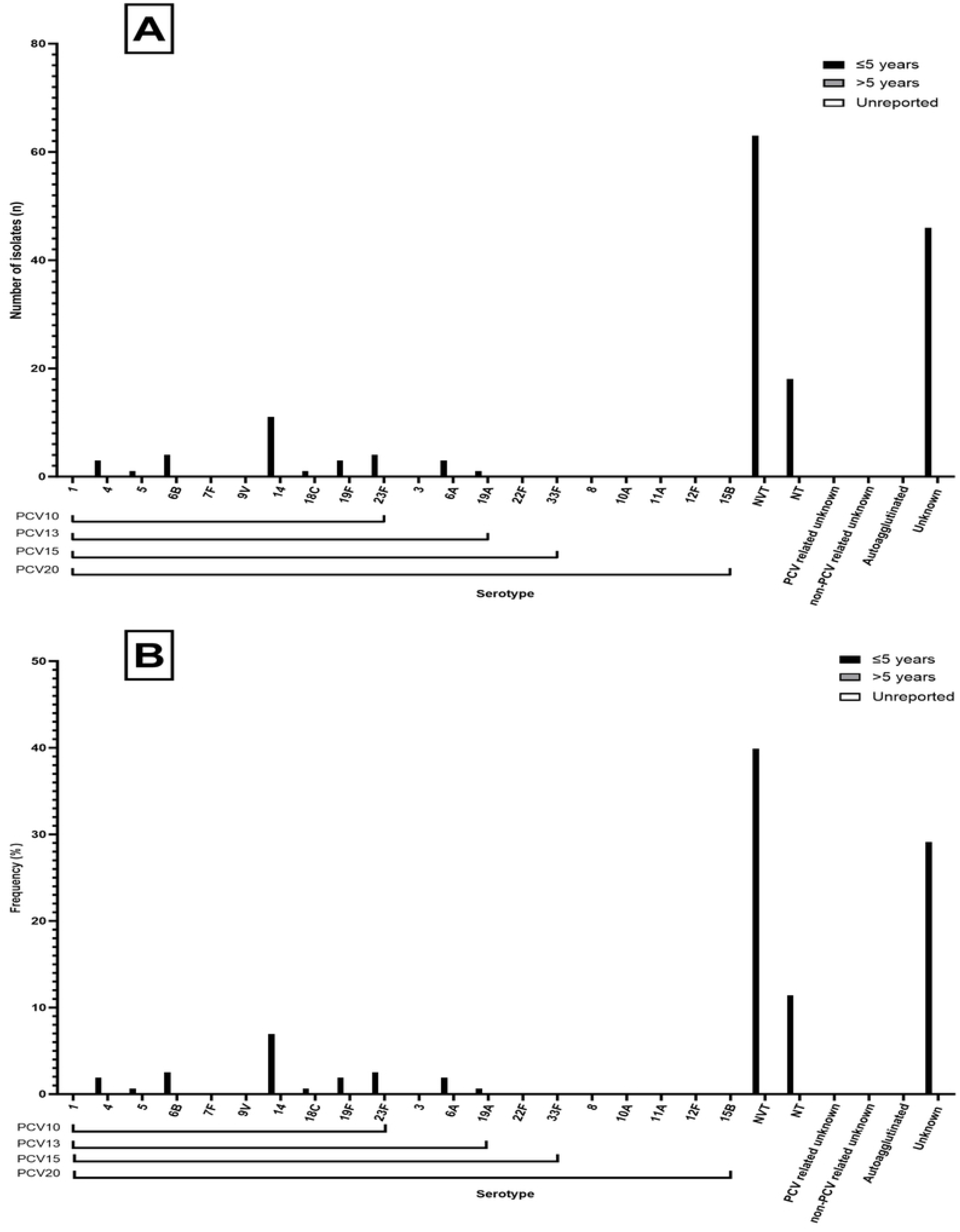
Serotypes from across Philippines carriage disease studies. (A) Number of isolates for each respective serotype found in PCV10, 13, 15 and 20, including number of NVTs and unknown serotypes from identified studies for which source could not be identified for Philippines carriage. (B) Frequency of isolates for each respective serotype found in PCV10, 13, 15 and 20, including number of NVTs and unknown serotypes from identified studies.

**Fig25.**
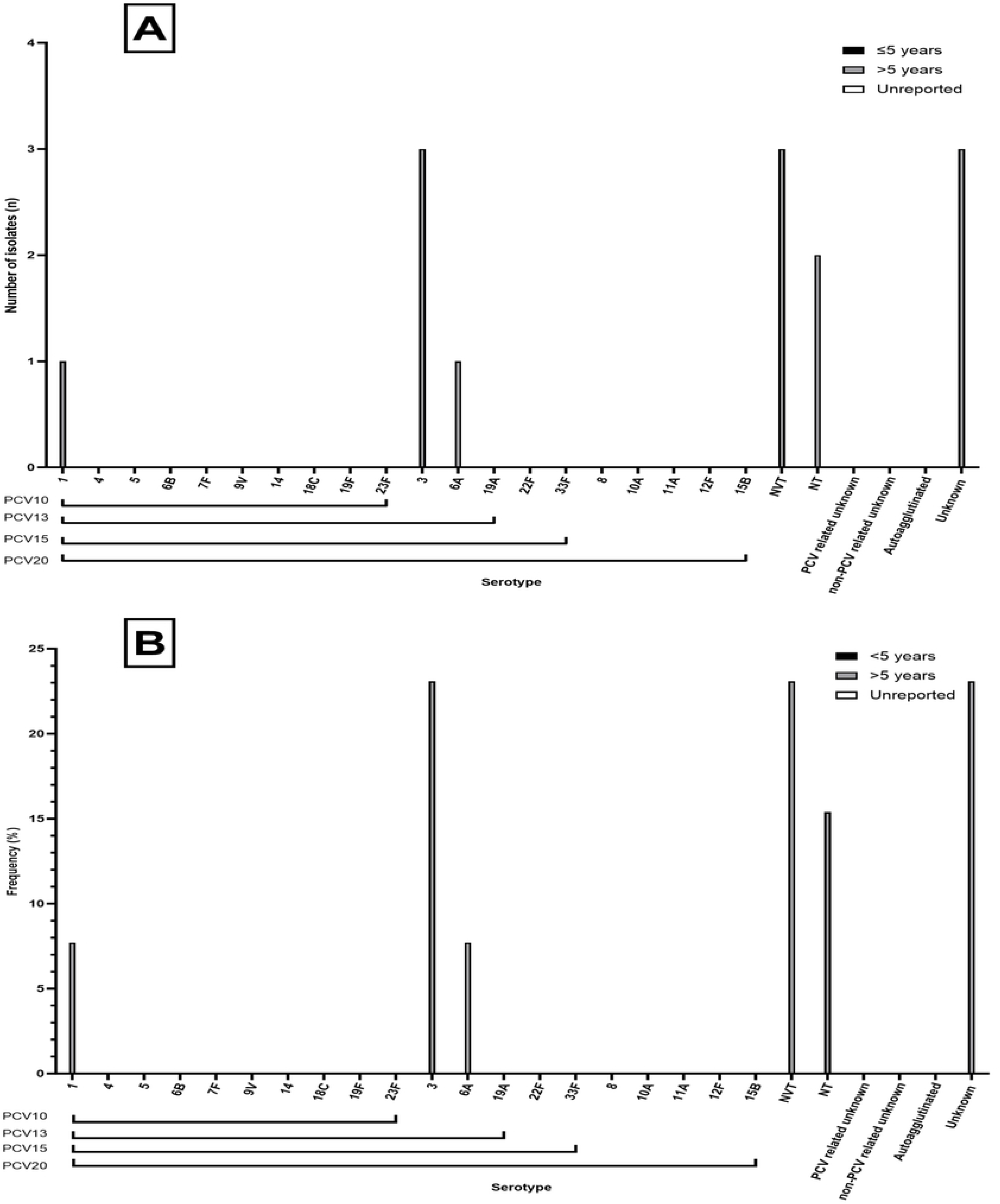
Serotypes from across Philippines studies where source could not be determined. (A) Number of isolates for each respective serotype found in PCV10, 13, 15 and 20, including number of NVTs and unknown serotypes from identified studies for which source could not be identified for Philippines unknown sources. (B) Frequency of isolates for each respective serotype found in PCV10, 13, 15 and 20, including number of NVTs and unknown serotypes from identified studies.

### Singapore

A total of n = 552 isolates were identified. N = 331 (59.96% [95%CI: 55.74 – 64.08]) were from IPD, zero were from non-IPD, n = 41 isolates from carriage (7.43%, [95%CI: 5.38 – 9.94]) and n = 180 (32.61%, [95%CI: 28.71 – 36.70]) were from unknown source. The most common vaccine types across all sources [Figure 9] were 19F (n = 96, 17.39% [95%CI: 14.32 - 20.82]), followed by 14 (n = 91, 16.49% [95%CI: 13.49 – 19.85]) and 23F (n = 74, 13.41% [95%CI: 10.67 – 16.54]) (Table 11). The three most common NVTs were 6 (n = 24, 4.35% [95%CI: 2.81 – 6.40]), 19 (n = 19, 3.44% [95%CI: 2.08 – 5.32]) and 20 (n = 10, 1.81% [95%CI: 0.87 – 3.31]). PCV10 and PCV13, PCV15 and PCV20 serotypes accounted for 60.51% [95%CI: 56.29 – 64.61], 69.20% [95%CI: 65.17 – 73.03], 70.83% [95%CI: 66.85 – 74.60] and 74.64% [95%CI: 70.79 – 78.22] of all isolates respectively. Non-typeables accounted for n = 12 (2.17%, [95%CI: 1.13 – 3.77]) and unknown serotypes accounted for n = 27 (4.89% [95%CI: 3.25 – 7.04]) of total isolates. Fig26, Fig27, Fig28, Fig29 show the split of serotype counts from across IPD, non-IPD, carriage and unknown sources.

**Table 11.** Serotype counts and percentages for all countries in Singapore, split by age category and invasive, non-invasive, carriage sources and from sources which could not be determined.

**Fig26.**
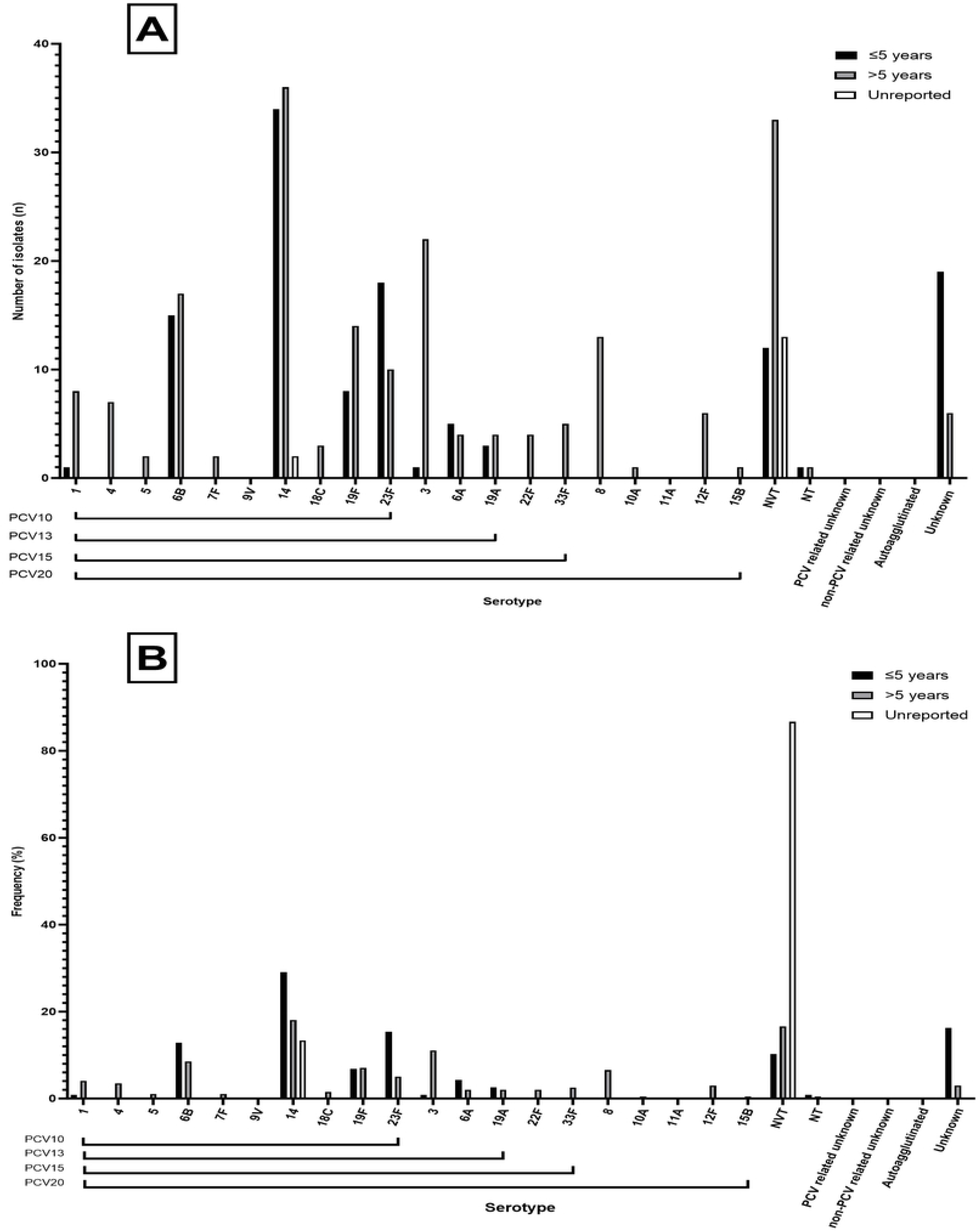
Serotypes from across Singapore invasive disease studies. (A) Number of isolates for each respective serotype found in PCV10, 13, 15 and 20, including number of NVTs and unknown serotypes from identified studies for which source could not be identified for Singapore invasive disease. (B) Frequency of isolates for each respective serotype found in PCV10, 13, 15 and 20, including number of NVTs and unknown serotypes from identified studies.

**Fig27.**
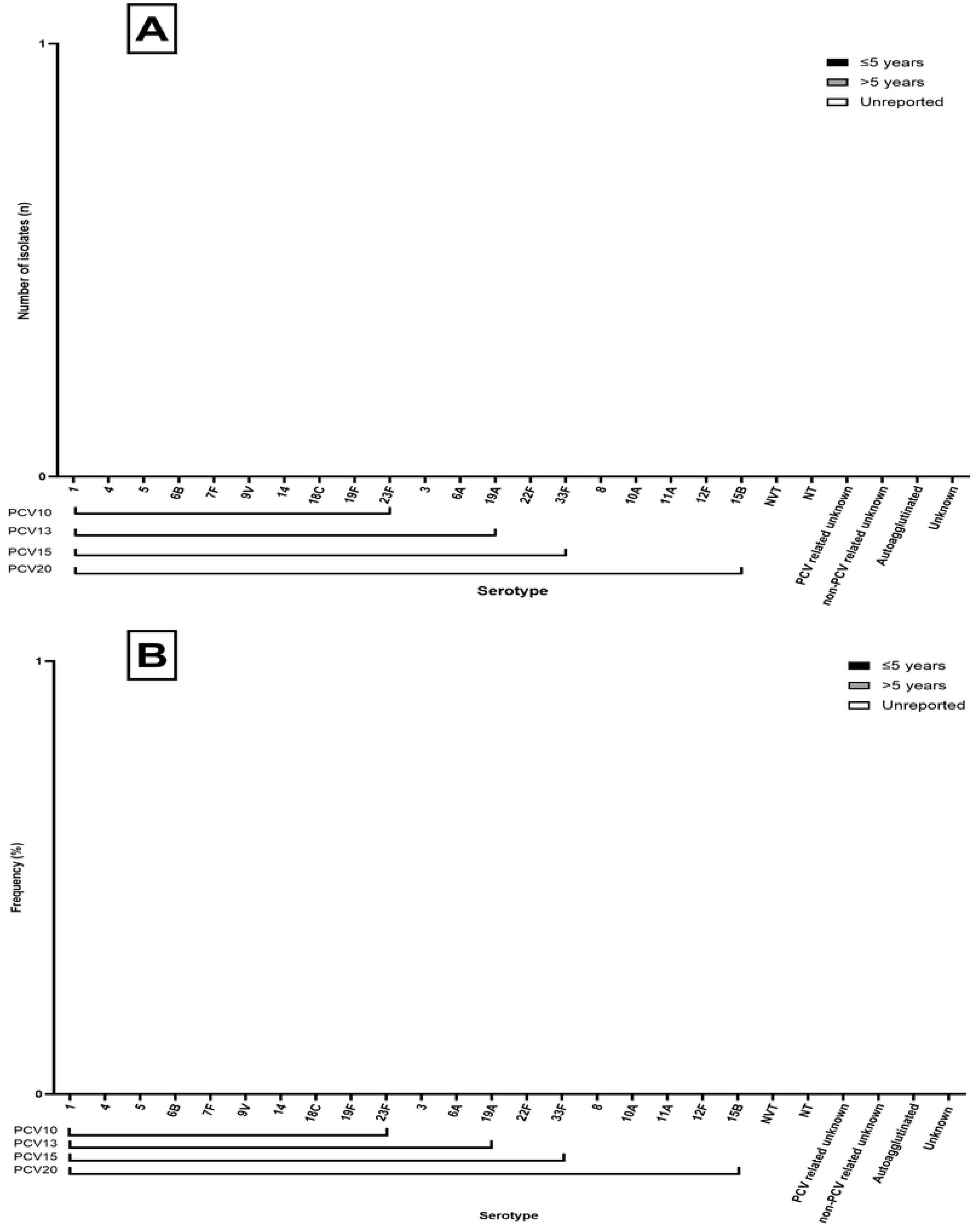
Serotypes from across Singapore non-invasive disease studies. (A) Number of isolates for each respective serotype found in PCV10, 13, 15 and 20, including number of NVTs and unknown serotypes from identified studies for which source could not be identified for Singapore non-invasive disease. (B) Frequency of isolates for each respective serotype found in PCV10, 13, 15 and 20, including number of NVTs and unknown serotypes from identified studies.

**Fig28.**
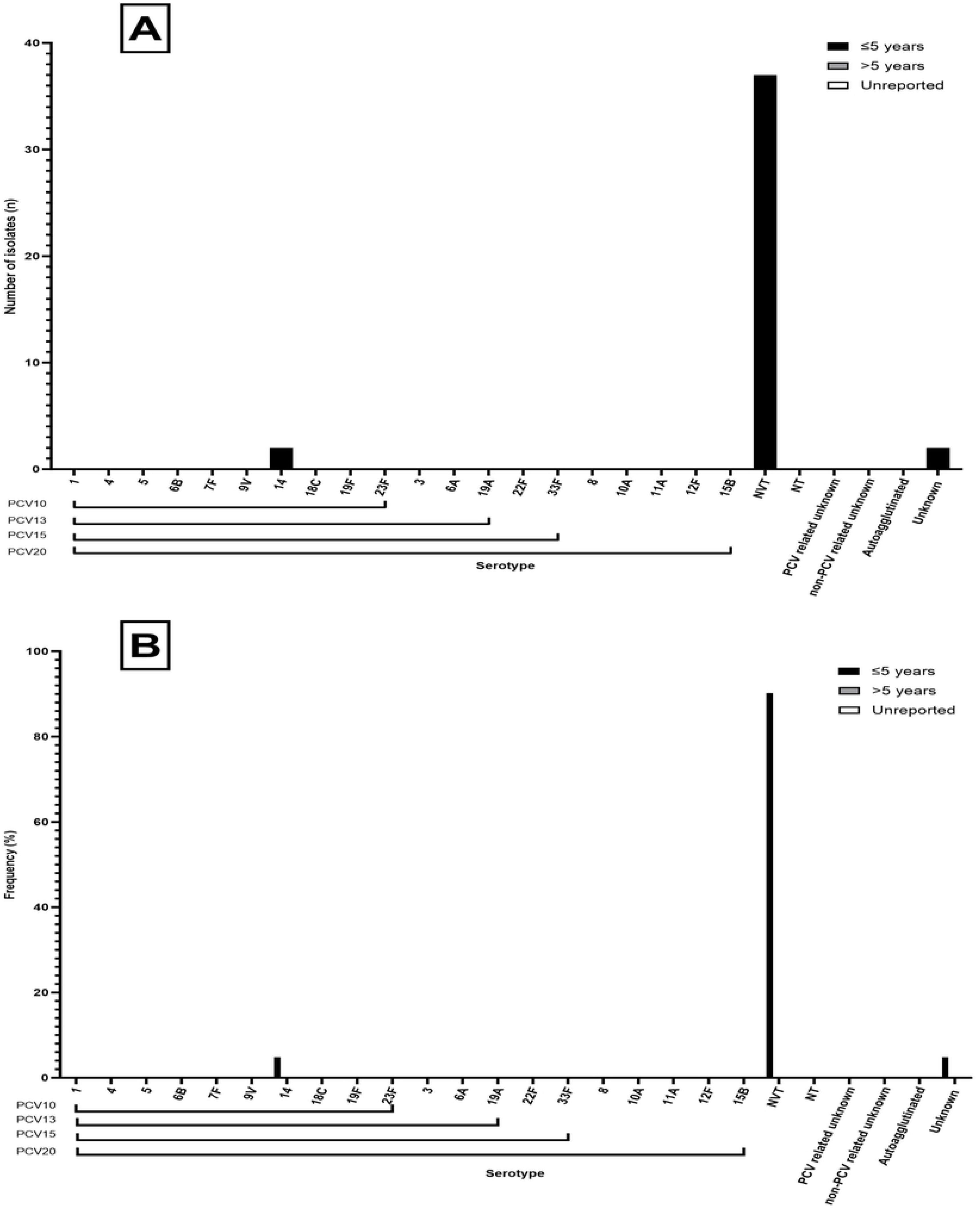
Serotypes from across Singapore carriage studies. (A) Number of isolates for each respective serotype found in PCV10, 13, 15 and 20, including number of NVTs and unknown serotypes from identified studies for which source could not be identified for Singapore carriage. (B) Frequency of isolates for each respective serotype found in PCV10, 13, 15 and 20, including number of NVTs and unknown serotypes from identified studies.

**Fig29.**
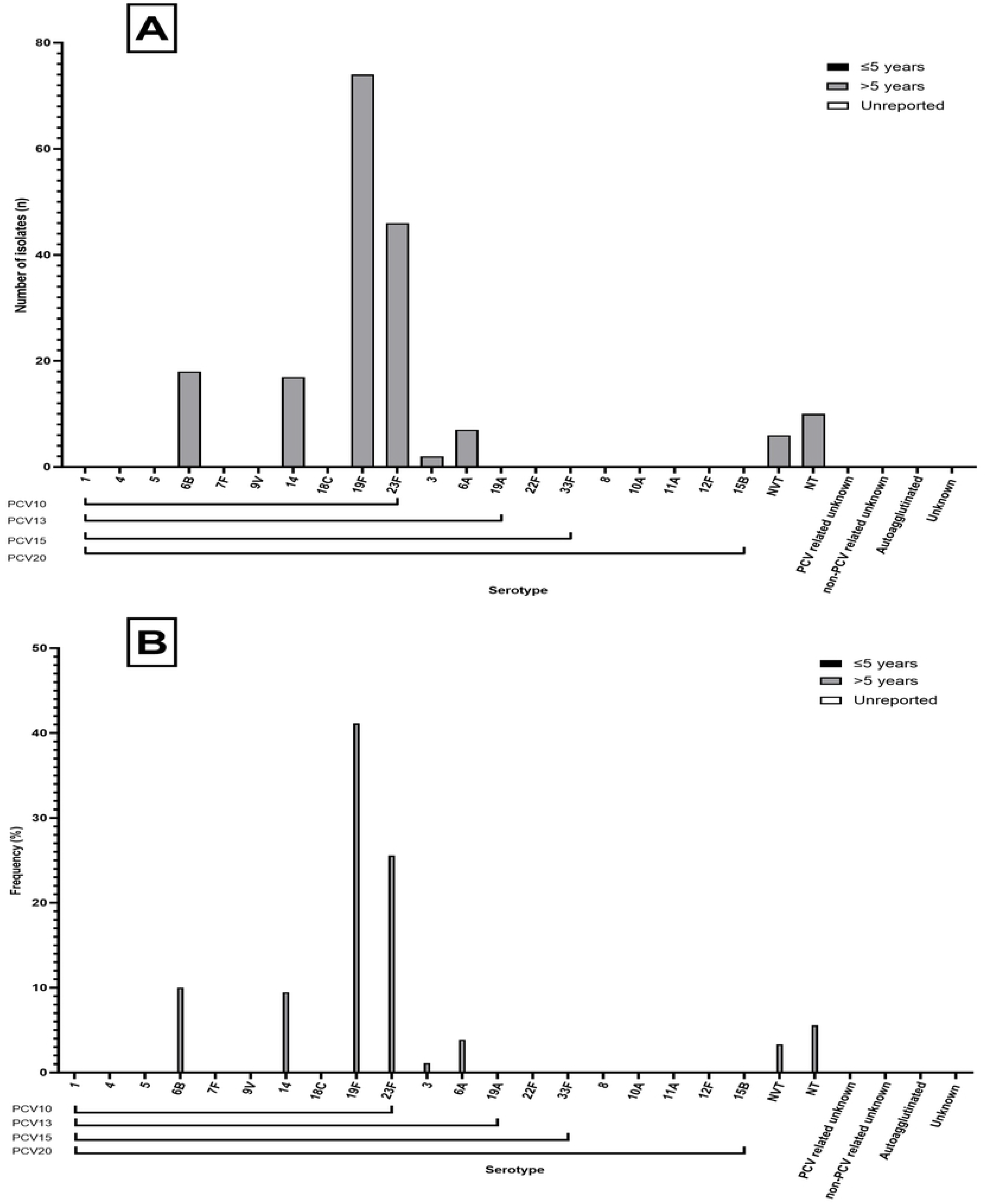
Serotypes from across Singapore studies where source could not be determined. (A) Number of isolates for each respective serotype found in PCV10, 13, 15 and 20, including number of NVTs and unknown serotypes from identified studies for which source could not be identified for Singapore unknown sources. (B) Frequency of isolates for each respective serotype found in PCV10, 13, 15 and 20, including number of NVTs and unknown serotypes from identified studies.

### Timor-Leste

No studies were identified for Timor-Leste.

### Thailand

A total of n = 3,016 isolates were identified. N = 1,887 (62.57% [95%CI: 60.81 – 64.30]) were from IPD, n = 202 isolates (6.70%, [95%CI: 5.83 – 7.65]) from non-IPD, n = 781 isolates from carriage (25.90%, [95%CI: 24.34 – 27.50]) and n = 146 (4.84%, [95%CI: 4.10 – 5.67]) were of unknown source. The most common vaccine types across all sources were 6B (n = 343, 11.37% [95%CI: 10.26 – 12.56]), followed by 23F (n = 263, 8.72% [95%CI: 7.74 – 9.78]) and 19F (n = 214, 7.10% [95%CI: 6.20 – 8.07]) (Table 12). The three most common NVTs were 6 (n = 147, 4.87%, [95%CI: 4.13 – 5.70]), 23 (n = 101, 3.35% [95%CI: 2.74 – 4.05]) and 19 (n = 64, 2.12%, [95%CI: 1.64 – 2.70]). PCV10 and PCV13, PCV15 and PCV20 serotypes accounted for 44.03% [95%CI: 42.25 – 45.82], 54.44% [95%CI: 52.65 – 56.23], 54.81% [95%CI: 53.01 – 56.59] and 55.97% [95%CI: 54.18 – 57.75] of all isolates respectively. Non-typeables accounted for n = 164 (5.44%, [95%CI: 4.66 – 6.31]) and unknown serotypes accounted for n = 574 (19.03% [95%CI: 17.64 – 20.48]) of total isolates. Fig30, Fig31, Fig32, Fig33 show the split of serotype counts from across IPD, non-IPD, carriage and unknown sources.

**Table 12.** Serotype counts and percentages for all countries in Thailand, split by age category and invasive, non-invasive, carriage sources and from sources which could not be determined.

**Fig30.**
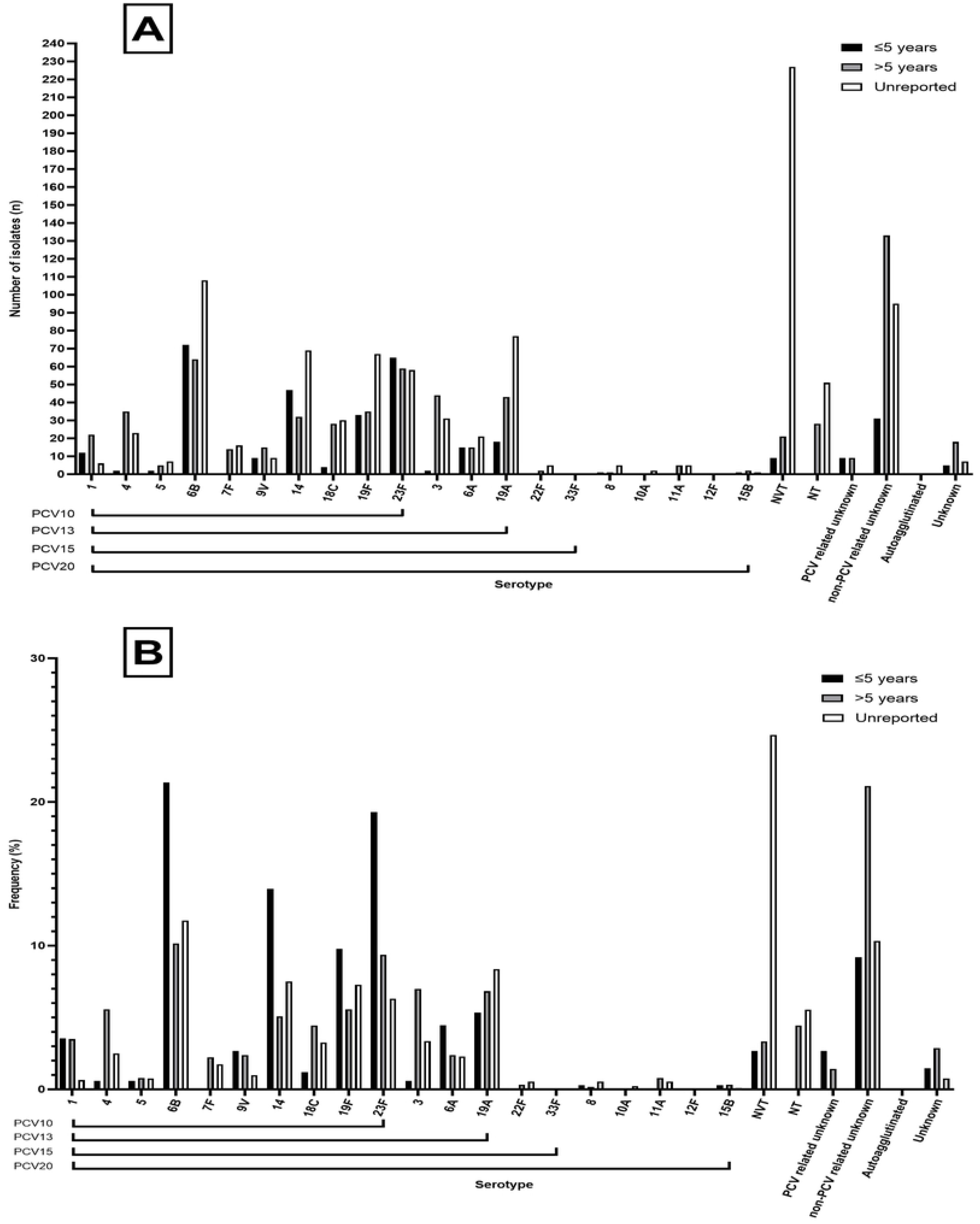
Serotypes from across Thailand invasive disease studies. (A) Number of isolates for each respective serotype found in PCV10, 13, 15 and 20, including number of NVTs and unknown serotypes from identified studies for which source could not be identified for Thailand invasive disease. (B) Frequency of isolates for each respective serotype found in PCV10, 13, 15 and 20, including number of NVTs and unknown serotypes from identified studies.

**Fig31.**
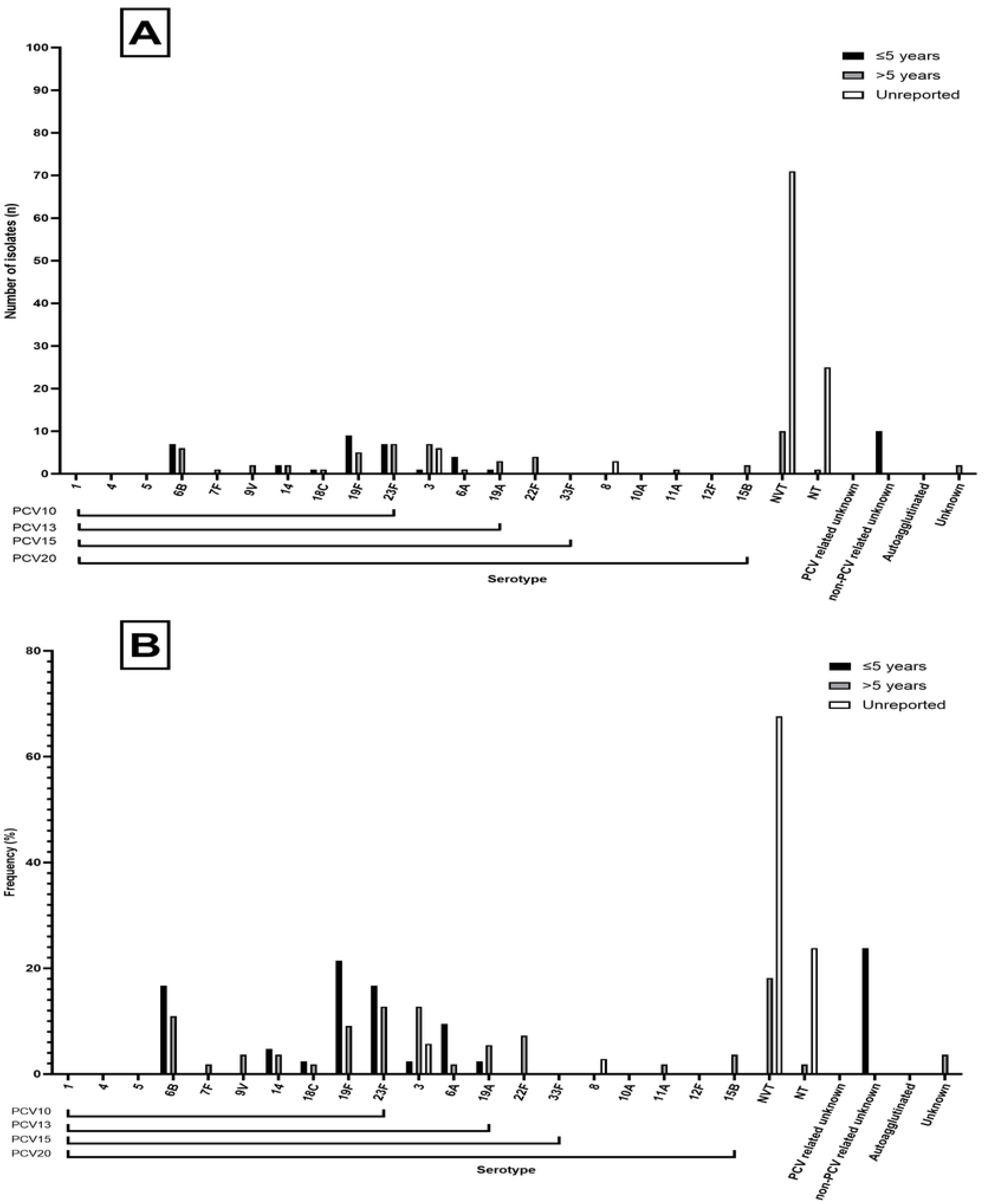
Serotypes from across Thailand non-invasive disease studies. (A) Number of isolates for each respective serotype found in PCV10, 13, 15 and 20, including number of NVTs and unknown serotypes from identified studies for which source could not be identified for Thailand non-invasive disease. (B) Frequency of isolates for each respective serotype found in PCV10, 13, 15 and 20, including number of NVTs and unknown serotypes from identified studies.

**Fig32.**
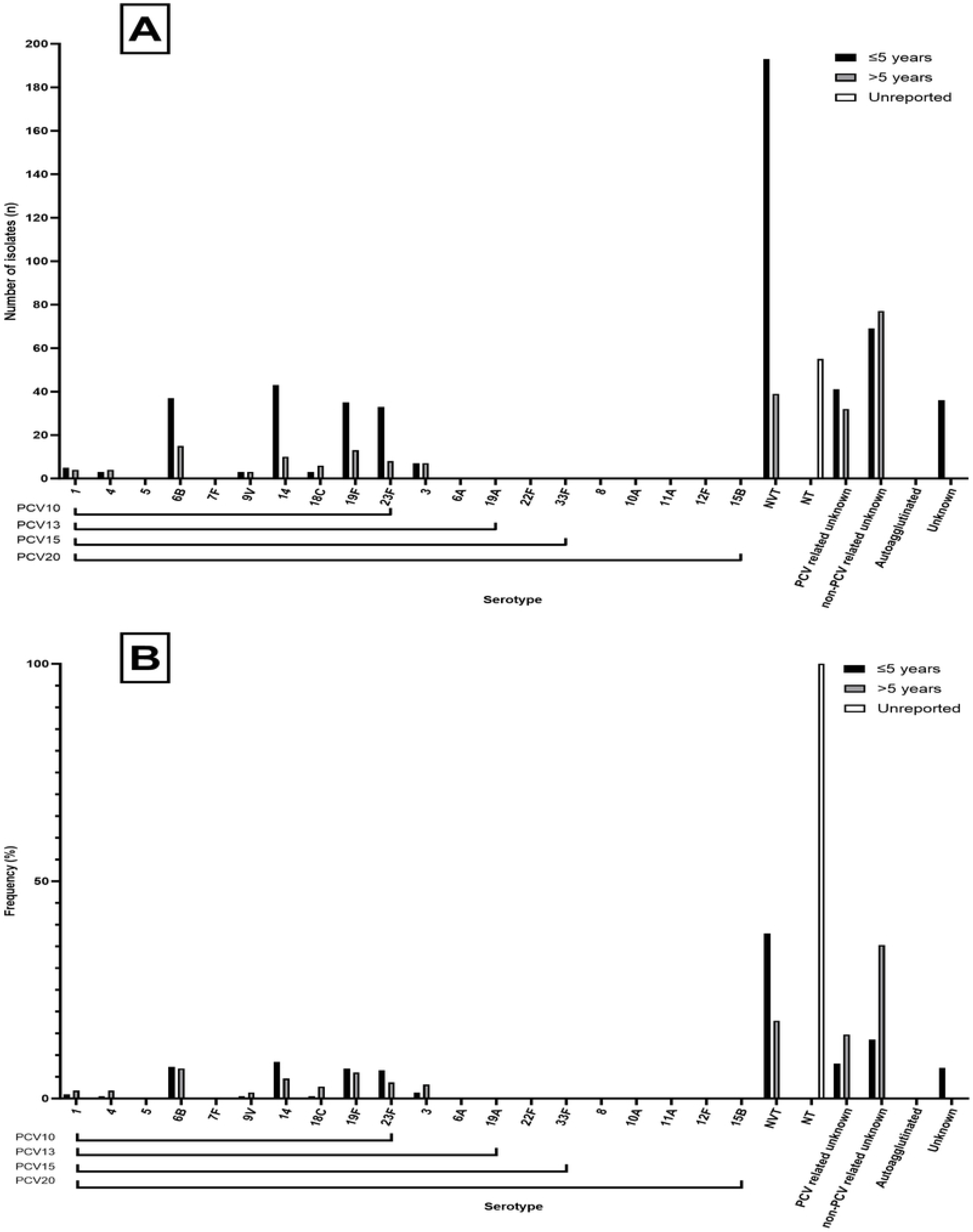
Serotypes from across Thailand carriage studies. (A) Number of isolates for each respective serotype found in PCV10, 13, 15 and 20, including number of NVTs and unknown serotypes from identified studies for which source could not be identified for Thailand carriage. (B) Frequency of isolates for each respective serotype found in PCV10, 13, 15 and 20, including number of NVTs and unknown serotypes from identified studies.

**Fig33.**
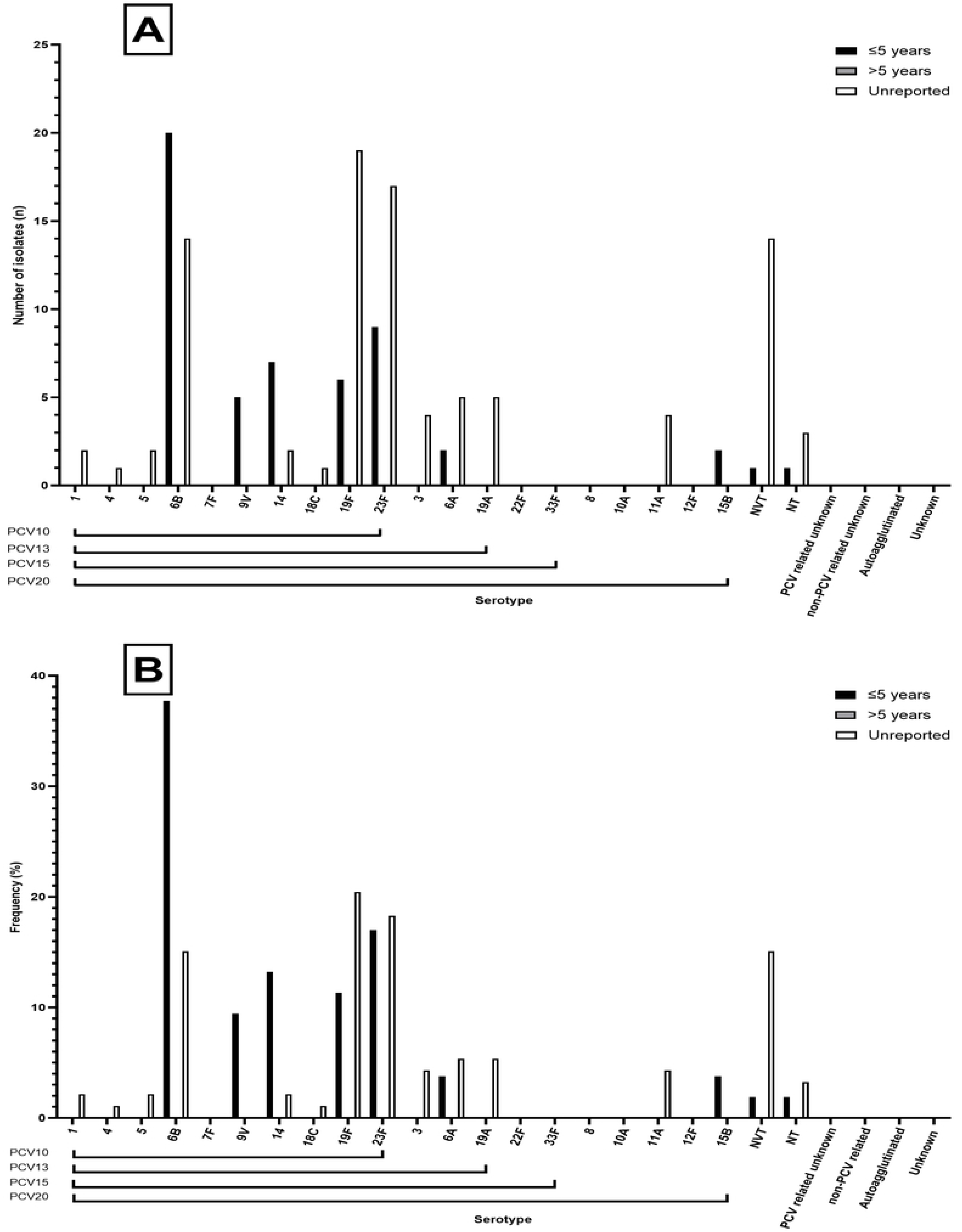
Serotypes from across Thailand studies where source could not be determined. (A) Number of isolates for each respective serotype found in PCV10, 13, 15 and 20, including number of NVTs and unknown serotypes from identified studies for which source could not be identified for Thailand unknown source. (B) Frequency of isolates for each respective serotype found in PCV10, 13, 15 and 20, including number of NVTs and unknown serotypes from identified studies.

### Vietnam

A total of n = 2,229 isolates were identified. N = 356 (15.97% [95%CI: 14.47 – 17.56]) were from IPD, n = 492 isolates (22.07%, [95%CI: 20.37 – 23.85]) from non-IPD and n = 1,381 isolates from carriage (61.96%, [95%CI: 59.90 – 63.98]). The most common vaccine types across all sources were 19F (n = 469, 21.04% [95%CI: 19.37 – 22.79]), followed by 23F (n = 264, 11.84% [95%CI: 10.53 – 13.26]) and 14 (n = 191, 8.57% [95%CI: 7.44 – 9.81]) (Table 13). The three most common NVTs were 6AB (n = 174, 7.81%, [95%CI: 6.73 – 9.00]), 19 (n = 104, 4.67% [95%CI: 3.83 – 5.63]) and 23 (n = 77, 3.45% [95%CI: 2.74 – 4.30]). PCV10 and PCV13, PCV15 and PCV20 serotypes accounted for 47.33% [95%CI: 45.24 – 49.43], 54.60% [95%CI: 52.50 – 56.68], 54.64% [95%CI: 52.55 – 56.73] and 55.81% [95%CI: 53.72 – 57.88] of all isolates respectively. Non-typeables accounted for n = 202 (9.06%, [95%CI: 7.90 – 10.33]) and unknown serotypes accounted for n = 79 (3.54% [95%CI: 2.82 – 4.40]) of total isolates. Fig34, Fig35, Fig36 show the split of serotype counts from across IPD, non-IPD and carriage sources.

**Table 13.** Serotype counts and percentages for all countries in Vietnam, split by age category and invasive, non-invasive and carriage sources.

**Fig34.**
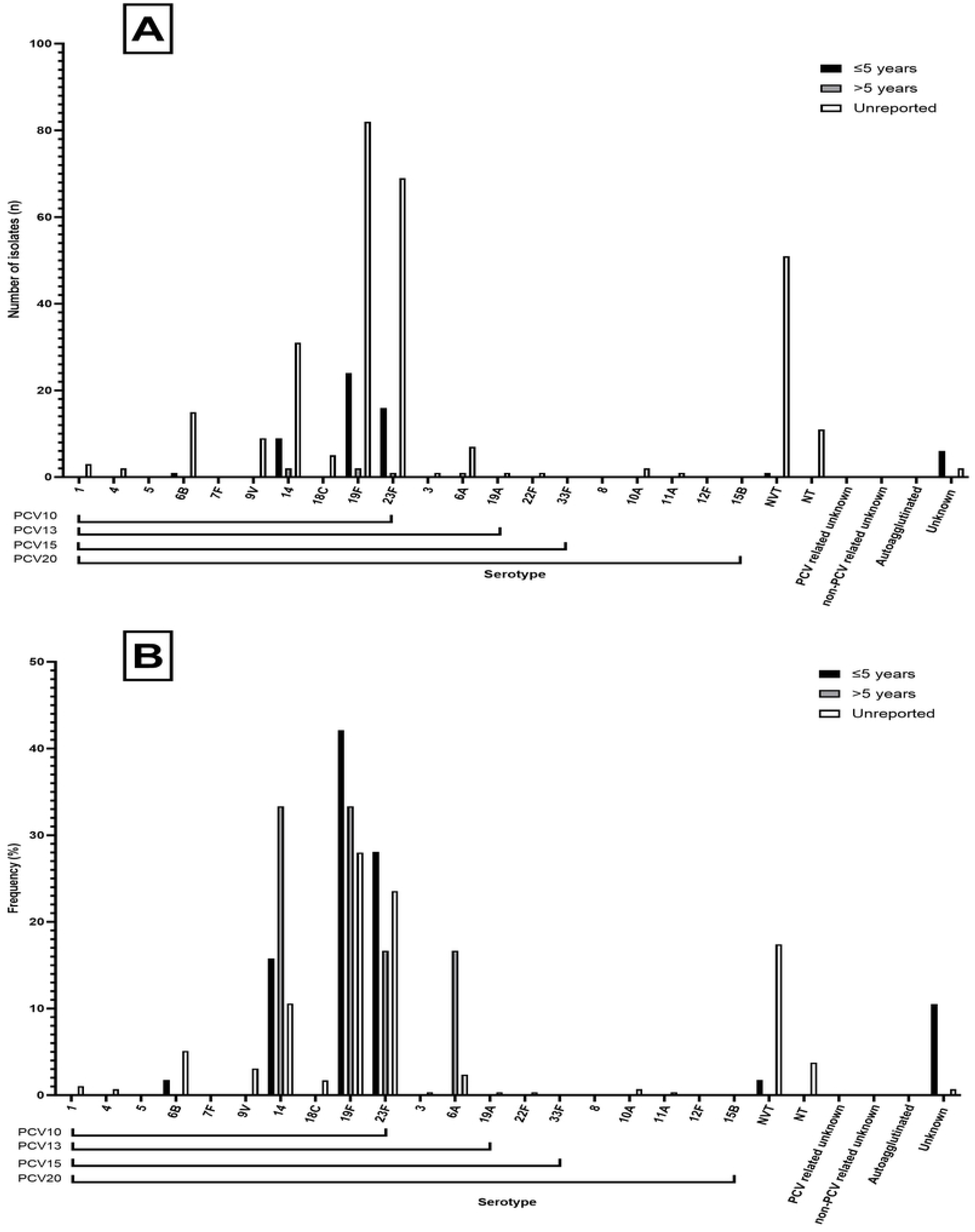
Serotypes from across Vietnam invasive disease studies. (A) Number of isolates for each respective serotype found in PCV10, 13, 15 and 20, including number of NVTs and unknown serotypes from identified studies for which source could not be identified for Vietnam invasive disease. (B) Frequency of isolates for each respective serotype found in PCV10, 13, 15 and 20, including number of NVTs and unknown serotypes from identified studies.

**Fig35.**
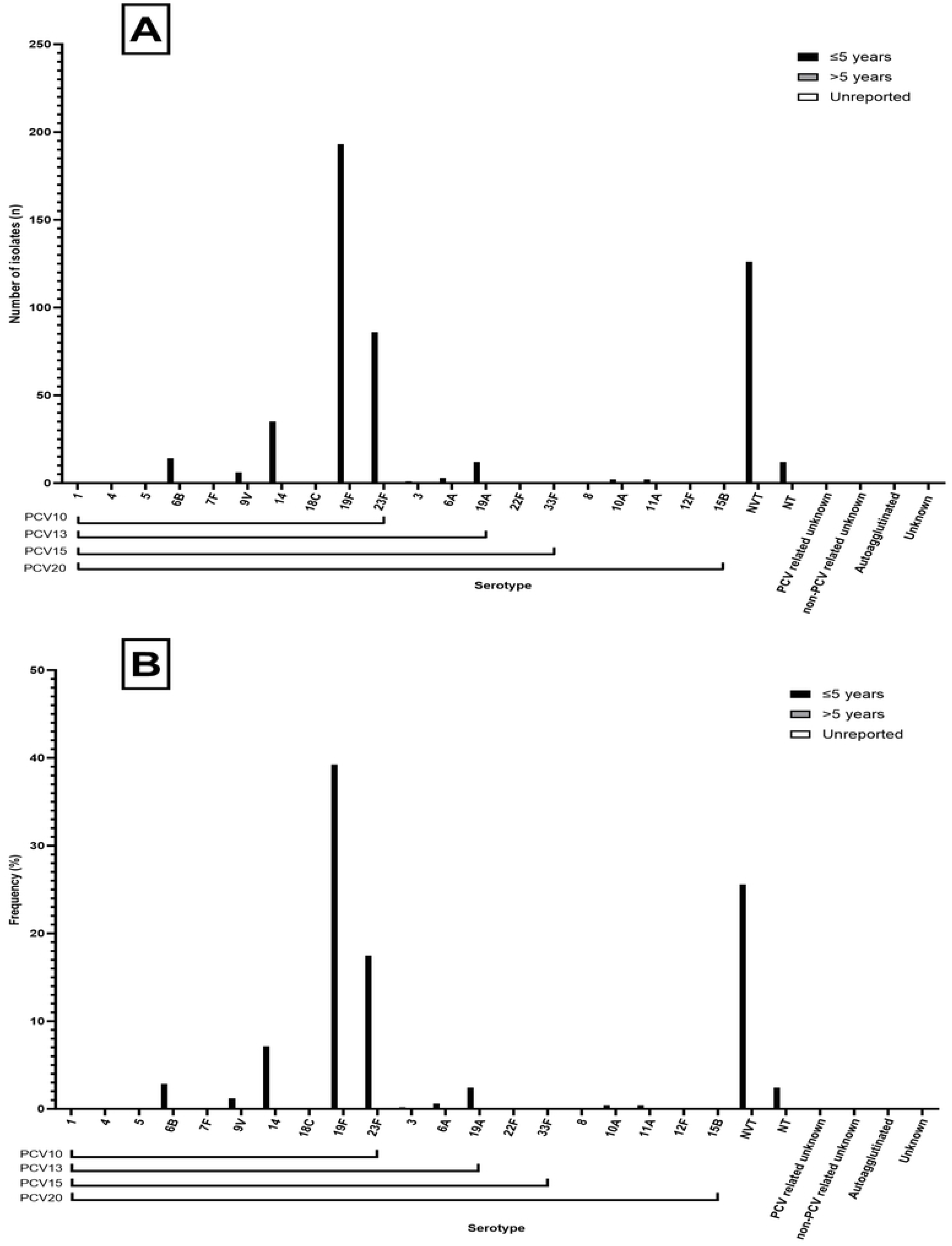
Serotypes from across Vietnam non-invasive disease studies. (A) Number of isolates for each respective serotype found in PCV10, 13, 15 and 20, including number of NVTs and unknown serotypes from identified studies for which source could not be identified for Vietnam non-invasive disease. (B) Frequency of isolates for each respective serotype found in PCV10, 13, 15 and 20, including number of NVTs and unknown serotypes from identified studies.

**Fig36.**
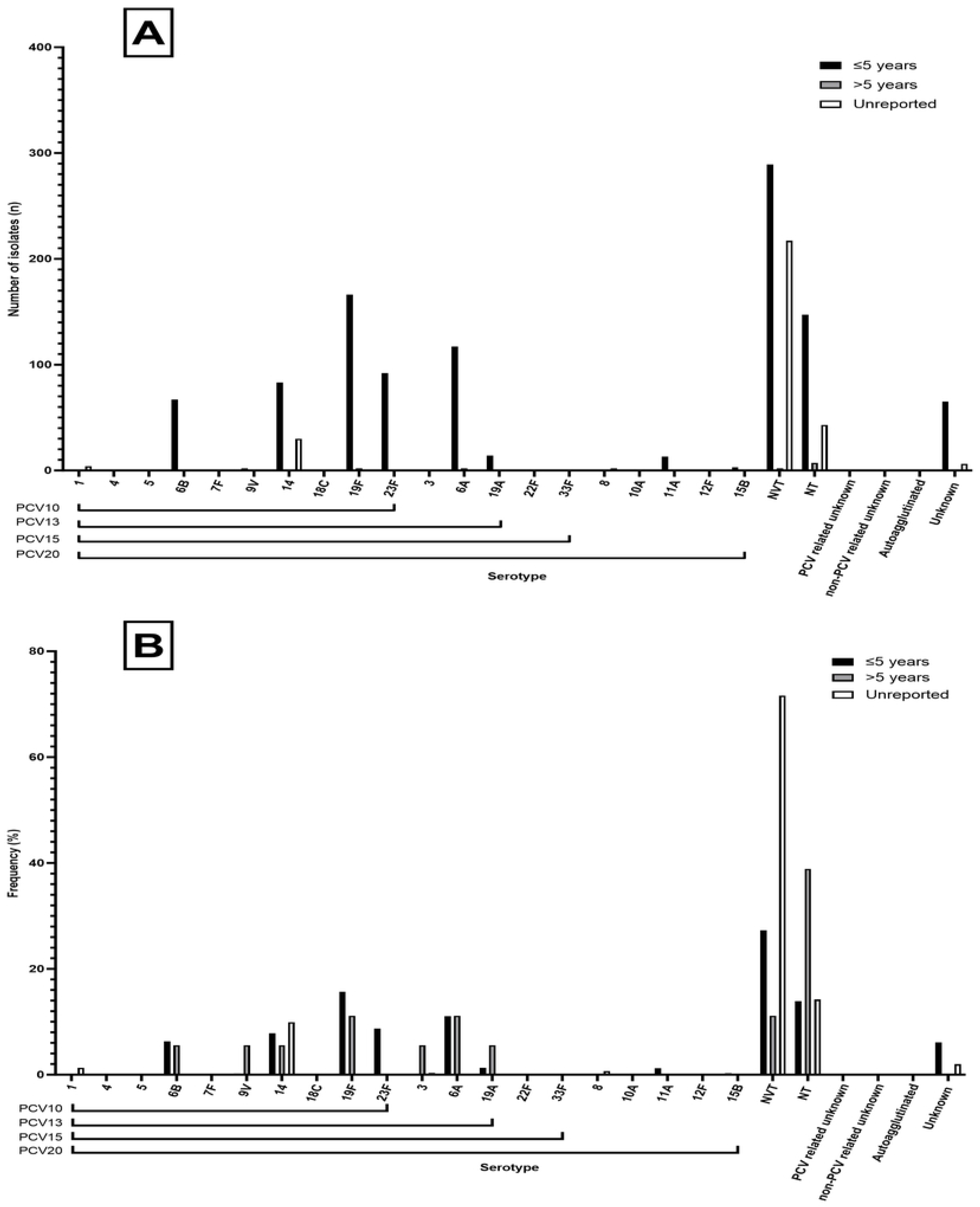
Serotypes from across Vietnam carriage studies. (A) Number of isolates for each respective serotype found in PCV10, 13, 15 and 20, including number of NVTs and unknown serotypes from identified studies for which source could not be identified for Vietnam carriage. (B) Frequency of isolates for each respective serotype found in PCV10, 13, 15 and 20, including number of NVTs and unknown serotypes from identified studies.

Percentage coverage represents the minimum coverage of each vaccine, as ‘unknown’ variables are included in the calculation, classed as serotypes not covered by vaccines. Vaccine coverage of serotypes are for all disease types, split by age category in each country. Coverage is calculated as the percentage of VT’s out of the total number of isolates, across PCV10, PCV13, PCV15 and PCV20, shown in Fig37.

**Fig37.** Forest plot of vaccine coverage for each of the currently licensed conjugates in each of the countries belonging to ASEAN. The plot shows the calculated coverage of conjugate-covered serotypes against the total number of serotypes identified in the studies.

PCV coverage over the three age categories and across pooled ASEAN data shows the lowest theoretical coverage, as unknown serotypes were also included in the calculation, under non-vaccine type numbers.

## Discussion

Despite high mortality in the ASEAN region associated with childhood pneumonia, pneumococcal serotype prevalence was incompletely understood. This study fills an important knowledge gap that has not been addressed since 2012.^24^ Over the last ten years the vaccination landscape has changed due to the introduction of the PCV13, as well as the development of other vaccines with higher valency. Additionally, some countries have since successfully implemented PCVs into their NIPs, while others have yet to do so. Serotype replacement in disease, the development of new vaccines and the heavy burden pneumonia continues to extract in the location necessitated this review of pneumococcal serotype epidemiology in Southeast Asia.

Based on our literature search and meta-analysis we found that the most common serogroups in IPD were 6 (23.19%), 19 (22.99%) and 23 (16.03%) whilst the most frequent serotypes were 1 (39.87%), 3 (28.39%), and 4 (21.09%). Notably, these results fit into the landscape of worldwide serotype prevalence in the pre-PCV era when serotypes 19F, 6B, 6A, and 23F were amongst the top ten most common serotypes observed in IPD, non-IPD and carriage in children <5 years of age (126, 127). These 10 serotypes were deemed to be responsible for >70% of IPDs globally, and 65-76% of paediatric carriage events in Europe and Asia prior to the introduction of PCVs. Our serogroup results are also in concordance with data reported in 2012, however the IPD causing serotypes have changed from 3, 34, and 1. Within serogroup 19, 19F was the most frequent which is especially worrying given its’ high rates of resistance to penicillin, a first-line antibiotic for pneumococcal infections. In addition, some strains of serotype 19F may also be resistant to other antibiotics, such as macrolides and fluoroquinolone.^128^ The use of conjugate vaccines can help prevent pneumococcal disease caused by antibiotic-resistant strains, and previous studies have proven their efficacy against serotype 19F in IPD (129–131). Within serogroup 6 and 23, the most frequently observed serotypes were 6B and 23F. Both are also included in all currently licenced and available PCVs. Moreover, all three most common IPD causing serotypes are also VTs - serotypes 1 and 4 are covered by PCV10 and above, and all three all included in PCV13 and other higher valent vaccines on the market. This demonstrates that ASEAN countries that are currently using PCV13 have likely achieved an adequate vaccine coverage and successfully mitigated pneumococcal diseases caused by these serotypes. However, it is important to note that these figures should be regarded as a conservative estimate due to differing levels of reporting accuracy and diagnosis between countries in ASEAN. Additionally, ongoing surveillance will be crucial, particularly for countries like Malaysia, which recently implemented PCV10 in their NIP. Monitoring will help assess the effectiveness of the vaccine and enable the adoption of a higher valency formulation if required.

The frequency of carried serotypes across countries and age groups slightly differed from those of IPD. Notably, the most carried serotype was 34, followed by serotypes 3 and 4. It is not uncommon to observe prevalent NVTs, such as 34, in countries where PCVs are not widely used or unavailable. For example, in 2015, the most common non-PCV13 isolates were serotype 34 in hospitalized children in Shanghai Children Hospital (132). This serotype is considered to have intermediate to high virulence i.e., capable of causing invasive diseases but less virulent than those of greater concern such as 1, 3, 7F, and 19A (133). Nevertheless, our data reveals an increase in IPDs caused by serotype 34, which ranked fourth among the most common serotypes (3.76%) in this disease group. This represents a rise compared to the 1% observed during the systematic review conducted in 2012 (24). It is important to note that newly emerging NVTs like serotype 34 may result from serotype-replacement after vaccination as was observed following the introduction of PCV7 in England (131). To date, serotype 34 is generally known to be susceptible to several antibiotics, including penicillin, amoxicillin, and some cephalosporins (134) however, resistance has been noted and therefore warrants continued vigilance (59, 135). The frequency of vaccine serotypes causing IPDs was the highest in Thailand, followed by Malaysia, Vietnam, Singapore, Philippines, Cambodia, Laos, Indonesia and Myanmar. Country-specific burdens however remain problematic to determine given the ongoing lack of data and this ranking should be viewed with caution. For example, the available data on IPD serotypes in Laos and Indonesia was limited and we were unable to locate any information on the prevalence of IPD in Myanmar. Contrastingly, in carriage Myanmar had the highest reported vaccine serotype prevalence, followed by Indonesia, Vietnam, Cambodia, Thailand, Malaysia, Laos, Philippines, and Singapore.

Most of the isolates (61.31%) in the ASEAN analysed in this study were from carriage. While asymptomatic colonization of pneumococci is common, individuals carrying highly virulent serotypes could be at risk of developing pneumococcal diseases. Therefore, the identification of serotypes carried in healthy individuals can help assess the circulating serotypes in the population and identify those that could potentially cause disease in vulnerable individuals and those who have not been vaccinated. Our study revealed that the predominant serotypes detected in carriage were consistent with those observed in IPDs. Specifically, serotypes 19F, 23F, and 6B were identified as the most common serotypes in both carriage and IPD. Therefore, pneumococcal carriage studies can aid us to predict the impact vaccines can make in preventing and assess impact.

Continued surveillance of pneumococcal serotypes is essential to track changes in their prevalence as vaccines are implemented. Countries such as Malaysia, which most recently have adopted PCV10, should pay close attention to ongoing surveillance of pneumococcal serotypes. This is particularly crucial since the vaccine formulation does not include other high-virulence serotypes such as 19A, which are known to cause IPD and was found the 2^nd^ most common serotype within serogroup 19 in our analysis and also overall the 3^rd^ most frequent serotype in Malaysia. With the increasing immunization of the population through PCV10, the number of vaccine serotypes in circulation is expected to decrease. Consequently, serotypes that are not covered by PCV10 may experience less competition and start to increase in prevalence. While PCV13 provides broader coverage of pneumococcal serotypes than PCV10, the latter includes protein D from non-typeable *Haemophilus influenzae* (NTHi) as a carrier for the vaccine’s pneumococcal serotypes. This may offer a more comprehensive protection against the non-invasive disease acute otitis media (AOM), as shown in a study (136). Although Synflorix® (PCV10) does not include serotype 19A in its selection, it may still provide some degree of cross-protection against this serotype. However, studies investigating this are inconclusive (137).

NT pneumococci accounted for 10.42% [95%CI: 9.95 – 10.89] of total serotypes. NT pneumococci can cause clinical disease, particularly in neonates and young infants, and studies have demonstrated an increase in NT carriage following the implementation of PCV (138). Further, their potential role in the spread of antimicrobial resistance has been shown with NT pneumococci exhibiting higher rates of recombination of genes *in vitro* compared with capsulated pneumococci (139). This highlights the importance of further studies so that epidemiology and pathogenesis can be better understood.

Despite the known advantages of using PCV the main barriers to widespread immunisation in MICs need to be acknowledged. Insufficient data reporting and the absence of pneumococcal surveillance programs pose challenges in evaluating the public health requirements of the population and consequently developing evidence-based public health policies (21). However, epidemiological data is vital for informing vaccine policy (140) which cannot rely on a single review or cost-benefit analysis. Instead, it necessitates continuous and rigorous research to establish a comprehensive evidence base. Cross-border collaboration has been promoted as a critical health improvement strategy (141) therefore, the ASEAN network should aid each other in the development of policy. Pneumococcal surveillance programmes are needed to better understand the pneumococcal burden, as well as track progress on vaccination programmes and offers insight into herd immunity effects (142).

The most recent country to update their pneumococcal vaccination policy is Malaysia opting to introduce PCV10, which has been accessible through the NIP since December 2020. This decision is beneficial as it will aid in reducing pneumococcal disease and lowering antimicrobial resistance in the country. Malaysia has been shown to have resistant pneumococci and particularly high penicillin resistance (123) along with multi-drug resistant pneumococci which is comparable to other countries in Asia (68). The implementation of PCV10 in Malaysia underscores the need for comprehensive pneumococcal surveillance programs across the country. As we have previously emphasized (143) such programs are critical for evaluating vaccine efficacy and informing policy decisions.

One of the key strengths of this study was its thorough search process, which involved searching multiple databases without limiting the publication start date to ensure that all relevant studies from the pre-PCV era were included. Additionally, our study has taken into consideration the potential use of the newly licensed PCV15 and PCV20 vaccines to determine their theoretical coverage of various serotypes. However, this study has certain limitations. The reviewed studies often had small sample sizes, non-standardized sampling methods, and inadequate duration. Moreover, the studies were frequently conducted in a single location within a country, thereby failing to provide a precise representation of the epidemiology across the broader population. A few studies only reported serogroups without conducting additional serotyping, while others reported ‘unknown’ serotypes. This makes it difficult to report increases in specific serotypes and fails to provide a comprehensive picture of serotype distribution. These findings also do not contribute meaningfully to the study calculations as it is unclear whether the serotypes are vaccine-types or not. Some studies included in this review did not account for antibiotic use or prior vaccination status of the participants, which would affect the serotypes observed.

This study focused on the serotype prevalence prior to the introduction of PCV vaccines, with the calculated vaccine coverages heavily reliant on the availability of data published during this time period. In order to gain a more accurate understanding of the actual vaccine coverage in the region, this study highlights the potential of conducting a comprehensive review and meta-analysis of the data published following the implementation of the vaccines.

## Conclusion

This study shows it is imperative that ongoing surveillance of pneumococci in the ASEAN region be established, utilising comparable approaches to ensure high-quality studies that yield valuable insights into vaccine policy and program efficacy. Comprehensive research encompassing IPD, non-IPD, and pneumococcal carriage is necessary to monitor changes in serotype distribution, assess the impact of vaccination on circulating serotypes and strengthen vaccine policies. Such efforts will help safeguard public health measures ultimately reducing the burden of pneumococcal disease in the most affected parts of the world.

## Data Availability

The data is available within the paper.

## List of Abbreviations

ASEAN: Association of Southeast Asian Nations
IPD: Invasive Pneumococcal Disease
LMIC: Low and Middle-Income Countries
Non-IPD: Non-Invasive Pneumococcal Disease
NT: Non-Typeable
NVT: Non-Vaccine Type
PCV: Pneumococcal Conjugate Vaccine
VRT: Vaccine-Related Types
VT: Vaccine Type

## Ethics approval and consent to participate

Not applicable.

## Consent for publication

Not applicable.

## Availability of data and Materials

All data generated or analysed during this study are included in this published article.

## Authors contributions

SCC planned the review. AJJL and ED wrote the initial draft and conducted the data analysis. DWC and SCC reviewed the draft manuscript and directed the edits. DWC and SCC reviewed the final draft and gave recommendations to additional research, structure and clarity. AJJL and ED made corrections and additions to the final draft and AJJL made the submission.

## Acknowledgements

This work arose from a dissertation submitted by AJJL in partial fulfilment of the requirements for the Degree of Master of Science (Public Health) at the University of Southampton.

## References

1. Cleary DW, Clarke SC. The nasopharyngeal microbiome. Emerging Topics in Life Sciences. 2017; 1(4): 297–312.

2. Ndlangisa K, du Plessis M, Allam M, Wolter N, de Fouveia L, Klugman K, et al. Invasive disease caused simultaneously by dul serotypes of streptococcus pneumoniae. Journal of Clinical Microbiology. 2017; 56(1).

3. Bogaert D, de Groot R, Hermans PWM. Streptococcus pneumoniae colonisation: the key to pneumococcal disease. The Lancet Infectious Diseases. 2004; 4(3): 144–154.

4. Ghimire M, Bhattacharya S, Narain J. Pneumonia in South-East Asia Region: Public Health perspective. The Indian Journal of Medical Research. 2012; 135: 459–468.

5. Rudan I, O’Brien KL, Nair H, Liu L, Theodoratou E, Qazi S, et al. Epidemiology and etiology of childhood pneumonia in 2010: estimates of incidence, severe morbidity, mortality, underlying risk factors and causative pathogens for 192 countries. Journal of Global Health. 2013; 3(1): 010401.

6. Global Pneumococcal Sequencing Project. Serotypes. [Online].; 2022. Cited 2023 03 02. Available from: https://www.pneumogen.net/gps/serotypes.html.

7. Papadatou I, Tzovara I, Licciardi PV. The Role of Serotype-Specific Immunological Memory in Pneumococcal Vaccination: Current Knowledge and Future Prospects. Vaccines (Basel). 2019; 7(1): 13.

8. Falkenhorst G, Remschmidt C, Harder T, Hummers-Pradier E, Wichmann O, Bogdan C. Effectiveness of the 23-valent pneumococcal polysaccharide vaccine (PPV23) against pneumococcal disease in the elderly: systematic review and meta-analysis. PLOS ONE. 2017; 12(1): e0169368.

9. Westerink MAJ, Schroeder HW, Nahm MH. Immune responses to pneumococcal vaccines in children and adults: rationale for age-specific vaccination. Aging and Disease. 2011; 3(1): 51–67.

10. European Medicines Agency. Synflorix. [Online].; 2023. Cited 2023 05 16. Available from: https://www.ema.europa.eu/en/medicines/human/EPAR/synflorix.

11. Berman-Rosa M, O’Donnell S, Barker M, Quach C. Efficacy and Effectiveness of the PCV-10 and PCV-13 Vaccines Against Invasive Pneumococcal Disease. Pediatrics. 2020; 145(4): e20190377.

12. CDC. Pneumococcal Vaccination: Summary of Who and When to Vaccinate. [Online].; 2023. Cited 2023 05 03. Available from: https://www.cdc.gov/vaccines/vpd/pneumo/hcp/who-when-to-vaccinate.html.

13. FDA. Vaxneuvance. [Online].; 2022. Cited 2022 07 10. Available from: https://www.fda.gov/vaccines-blood-biologics/vaccines/vaxneuvance.

14. Kobayashi M, Farrar JL, Gierke R, Britton A, Childs L, Leidner AJ, et al. Use of 15-Valent Pneumococcal Conjugate Vaccine and 20-Valent Pneumococcal Conjugate Vaccine Among U.S. Adults: Updated Recommendations of the Advisory Committee on Immunization Practices - United States, 2022. Morbidity and Mortality Weekly Report. 2022; 71(4): 109–117.

15. EMA. Vaxneuvance. [Online].; 2022. Cited 2023 03 02. Available from: https://www.ema.europa.eu/en/medicines/human/EPAR/vaxneuvance.

16. Pfizer. European Medicines Agency Approves Pfizer’s 20-Valent Pneumococcal Conjugate Vaccine Against Invasive Pneumococcal Disease and Pneumonia in Adults. [Online].; 2022. Cited 2022 07 10. Available from: https://www.pfizer.com/news/press-release/press-release-detail/european-medicines-agency-approves-pfizers-20-valent#:~:text=On%20June%208%2C%202021%2C%20Pfizer,age%2018%20years%20or%20older.

17. CDC. Pneumococcal Vaccination: What Everyone Should Know. [Online]. Cited 2023 02 27. Available from: https://www.cdc.gov/vaccines/vpd/pneumo/public/index.html.

18. EMA. Apexxnar. [Online].; 2022. Cited 2023 03 02. Available from: https://www.ema.europa.eu/en/medicines/human/EPAR/apexxnar.

19. Pharmaceutical Journal. MHRA approves first vaccine in ten years to protect children against pneumococcal diseases. The Pharmaceutical Journal. 2022; 309(7967): 7967.

20. Chapman R, Sutton K, Dillon-Murphy D, Patel S, Hilton B, Farkouh R, et al. Ten year public health impact of 13-valent pneumococcal conjugate vaccination in infants: A modelling analysis. Vaccine. 2020; 38(45): 7138–7145.

21. Tricarico S, McNeil HC, Head MG, Cleary DW, Clarke SC. Informing pneumococcal conjugate vaccine policy in middle income countries: The case of Malaysia. Vaccine. 2017; 18(35): 2288–2290.

22. UNICEF. Pneumonia. [Online].; 2022. Cited 2023 05 03. Available from: https://data.unicef.org/topic/child-health/pneumonia/.

23. Gamil A, Chokephabulkit K, Phongsamart W, Techasaensiri C, Pirilam B, Thamaree R. Pneumococcal disease in Thailand. International Journal of Infectious Diseases. 2021; 102(1): 429–436.

24. Jauneikaite E, Jefferies J, Hibberd M, Clarke SC. Prevalence of Streptococcus pneumoniae serotypes causing invasive and non-invasive disease in South East Asia: A review. Vaccine. 2012; 30(24): 3503–3514.

25. CASP. Systematic Review. [Online].; 2023. Cited 2023 02 14. Available from: https://casp-uk.net/glossary/systematic-review/.

26. JBI. Checklist for prevalence studies. [Online].; 2023. Cited 2023 02 14. Available from: https://jbi.global/sites/default/files/2019-05/JBI_Critical_Appraisal-Checklist_for_Prevalence_Studies2017_0.pdf.

27. Turner P, Turner C, Jankhot A, Helen N, Lee SJ, Day NP, et al. A longitudinal study of Streptococcus pneumoniae carriage in a cohort of infants and their mothers on the Thailand-Myanmar border. PLOS ONE. 2012; 7(5): e38271.

28. Turner P, Turner C, Suy K, Soeng S, Ly S, Miliya T. Pneumococcal Infection among Children before Introduction of 13-Valent Pneumococcal Conjugate Vaccine, Cambodia. Emerging Infectious Diseases. 2015; 21(11): 2080–2083.

29. Moore C, Giess A, Soeng S, Sar P, Kumar V, Nhoung P, et al. Characterisation of invasive streptococcus pneumoniae isolates from Cambodian children between 2007 - 2012. PLOS ONE. 2016; 11(7): e0159358.

30. Inghammar M, By Y, Farris C, Phe T, Borand L, Kerleguer A, et al. Serotype Distribution of Clinical Streptococcus pneumoniae Isolates before the Introduction of the 13-Valent Pneumococcal Conjugate Vaccine in Cambodia. The American Journal of Tropical Medicine and Hygiene. 2018; 98(3): 791–796.

31. Dananché C, Paranhos-Baccalà G, Messaoudi M, Sylla M, Awasthi S, Bavdekar A, et al. Serotypes of Streptococcus pneumoniae in Children Aged >

32. Turner P, Leab P, Ly S, Sao S, Miliya T, Heffelfinger JD, et al. Impact of 13-Valent Pneumococcal Conjugate Vaccine on Colonization and Invasive Disease in Cambodian Children. Clinical Infectious Diseases. 2020; 70(8): 1580–1588.

33. Farida H, Severin JA, Gasem MH, Keuter M, Wahyono H, van den Broek P, et al. Nasopharyngeal Carriage of Streptococcus pneumonia in Pneumonia-Prone Age Groups in Semarang, Java Island, Indonesia. PLOS ONE. 2014; 9(1): e87431.

34. Hadinegoro SR, Prayitno A, Khoeri MM, Djelantik IGG, Dewi NE, Indriyani SAK, et al. Nasopharyngeal carriage of streptococcus pneumoniae in healthy children under five years old in central Lombok regency, Indonesia. The Southeast Asian journal of tropical medicine and public health. 2016; 47(3): 485–493.

35. Harimurti K, Saldi SRF, Dewiasty E, Khoeri MM, Yunihastuti E, Putri T, et al. Nasopharyngeal carriage of Streptococcus pneumoniae in adults infected with human immunodeficiency virus in Jakarta, Indonesia. Journal of Infection and Public Health. 2016; 9(5): 633–638.

36. Dunne EM, Murad C, Sudigdoadi S, Fadlyana E, Tarigan R, Indriyani SAK, et al. Carriage of Streptococcus pneumoniae, Haemophilus influenzae, Moraxella catarrhalis, and Staphylococcus aureus in Indonesian children: A cross-sectional study. PLOS ONE. 2018; 13(4): e0195098.

37. Safari D, Kurniati N, Waslia L, Khoeri MM, Putri T, Bogaert D, et al. Serotype Distribution and Antibiotic Susceptibility of Streptococcus pneumoniae Strains Carried by Children Infected with Human Immunodeficiency Virus. PLOS ONE. 2014; 9(10): e110526.

38. Safari D, Mudaliana S, Harimurti K, Waslia L, Subekti D. Nasopharyngeal colonization of streptococcus pneumoniae in elderly people in Jakarta Indonesia. International Journal of Infectious Diseases. 2012; 16(1): E313.

39. Soewignjo S, Gessner BD, Sutanto A, Steinhoff M, Prijanto M, Nelson C, et al. Streptococcus pneumoniae nasopharyngeal carriage prevalence, serotype distribution, and resistance patterns among children on Lombok Island, Indonesia. Clinical Infectious Diseases. 2001; 32(7): 1039–43.

40. Said WF, Sukoto E, Khoeri MM, Kumalawati J, Safari D. Serotype distribution and antimicrobial susceptibility of Streptococcus pneumoniae isolates from adult patients in Jakarta, Indonesia. Journal of Infection and Public Health. 2017; 10(6): 833–835.

41. Yuliarti K, Hadinegoro SR, Supriyatno B, Karuniawati A. Invasive pneumococcal disease among hospitalized children aged 28 days to 60 months in Jarkarta. Southrast Asian Journal of Tropical Medicine and Public Health. 2012; 43(1): 136–144.

42. Amanda G, Tafroji W, Sutoyo DK, Burhan E, Haryanto B, Safari D. Serotype distribution and antimicrobial profile of Streptococcus pneumoniae isolated from adult patients with community-acquired pneumonia in Jakarta, Indonesia. Journal of Microbiology, Immunology and Infection. 2021; 54(6): 1175–1178.

43. Murad C, Dunne EM, Sudigdoadi S, Fadlyana E, Tarigan R, Pell CL, et al. Pneumococcal carriage, density, and co-colonization dynamics: A longitudinal study in Indonesian infants. International Journal of Ifectious Diseases. 2019; 86: 73–81.

44. Purwanto DS, Loho T, Tafroji W, Mangunatmadja I, Immanuel S, Timan IS, et al. Isolation and identification of Streptococcus pneumoniae serotype 6B from a patient with bacterial meningitis infection in Jakarta, Indonesia. Access Microbiology. 2020; 2(5).

45. Safari D, Mudaliana S, Harumurti K, Waslia L, Subekti D. Staphylococcus aureus and Streptococcus pneumoniae prevalence among elderly adults in Jakarta, Indonesia. Southeast Asian Journal of Tropical Medicine and Public Health. 2012; 16(1): e313.

46. Safari D, Valentiya F, Salsabila K, Paramaiswari WT, Tafroji W, Hammerschmidt S, et al. The prevalence of pilus islets in Streptococcus pneumoniae isolates from healthy children in Indonesia. Access Microbiology. 2021; 3(1): acmi000184.

47. Wahyono DJ, Khoeri MM, Darmawan AB, Wijayanti SPM, Mumpuni A, Nawangtantri G, et al. Nasopharyngeal carriage rates and serotype distribution of Streptococcus pneumoniae among school children with acute otitis media in Central Java, Indonesia. Access Microbiology. 2021; 3(7): 000249.

48. Salsabila K, Paramaiswari WT, Amalia H, Ruyani A, Tafroji W, Winarti Y, et al. Nasopharyngeal carriage rate, serotype distribution, and antimicrobial susceptibility profile of Streptococcus pneumoniae isolated from children under five years old in Kotabaru, South Kalimantan, Indonesia. Journal of Microbiology, Immunology and Infection. 2022; 55(3): 482–488.

49. Harimurti K, Saldi SRF, Dewiasty E, Alfarizi T, Dharmayuli M, Khoeri MM, et al. Streptococcus pneumoniae carriage and antibiotic susceptibility among Indonesian pilgrims during the Hajj pilgrimage in 2015. PLoS One. 2021; 16(1): e0246122.

50. Muktiari D, Khoeri MM, Tafroji W, Waslia L, Safari D. Serotypes and antibiotic susceptibility profile of Streptococcus pneumoniae isolated from nasopharynges of children infected with HIV in Jakarta, Indonesia, pre- and post-pneumococcal vaccination. Access Microbiology. 2021; 3(3): 000215.

51. Prayitno A, Supriyatno B, Munasir Z, Karuniawati A, Hadinegoro SRS, Prihartono J, et al. Pneumococcal nasopharyngeal carriage in Indonesia infants and toddlers post-PCV13 vaccination in a 2+1 schedule: A prospective cohort study. PLoS ONE. 2021; 16(1): e0245789.

52. Safari D, Widhidewi NW, Paramaiswari WT, Lila Paramasatiari AAA, Maharani Putri HF, A Asih Ratnadi IG, et al. Prevalence, Serotype Distribution, and Antimicrobial Susceptibility Profile of Streptococcus pneumoniae among Patients with Acute Respiratory Tract Infection. WHO South-East Asia Journal of Public Health. 2021; 10(2): 66–69.

53. Moore CE, Sengduangphachanh A, Thaojaikon T, Sirisouk J, Foster D, Phetsouvanh R, et al. Enhanced determination of Streptococcus pneumoniae serotypes associated with invasive disease in Laos by using a real-time polymerase chain reaction serotyping assay with cerebrospinal fluid. American Society of Tropical Medicine and Hygiene. 2010; 83(3): 451–457.

54. Satzke C, Dunne EM, Choummanivong M, Ortika BD, Neal EFG, Pell CL, et al. Pneumococcal carriage in vaccine-eligible children and unvaccinated infants in Lao PDR two years following the introduction of the 13-valent pneumococcal conjugate vaccine. Vaccine. 2019; 37(2): 296–305.

55. Cheong YM, Jegathesan M, Henrichsen J, Wong YH, Ng AJ, Louis A. Antibiotic susceptibility and serotype distribution of Streptococcus pneumoniae in Malaysian children. Journal of Tropical Pediatrics. 1988; 34(4): 182–185.

56. Shakrin N, Masri S, Taib N, Nordin S, Jamal F, Desa M. Genotypic characterization of Malaysian human isolates of Streptococcus pneumoniae from carriage and clinical sources. Comparative Immunology, Microbiology and Infectious Disease. 2014; 37(5-6): 347–354.

57. Rohani M, Raudzah A, Ng A, Ng P, Zaidatul A, Asmah I, et al. Epidemiology of Streptococcus pneumoniae infection in Malaysia. Epidemiology and Infection. 1999; 122(1): 77–82.

58. Desa M, Lin T, Yasin R, Parasakthi N. Penicillin susceptibility and molecular characteristics of clinical isolates of Streptococcus pneumoniae at the University of Malaya Medical Center, Kuala Lumpur, Malaysia. International Journal of Infectious Diseases. 2003; 7(3): 190–197.

59. Le C, Palanisamy N, Mohd Yusof M, Sekaran S. Capsular seroytpe and antibiotic resistance of streptococcus pnuemoae isolates in Malaysia. PLOS ONE. 2011; 6(5): e19547.

60. Jefferies J, Mohd Yusof M, Devi Sekaran S, Clarke S. Novel Clones of Streptococcus pneumoniae causing invasive disease in Malaysia. PLOS ONE. 2014; 9(6): e97912.

61. Rohani M, Zin N, Hussin A, Nawi S, Hanapiah S, Wahab Z, et al. Current trend of pneumococcal serotypes distribution and antibiotic susceptibility pattern in Malaysian hospitals. Vaccine. 2011; 29(34): 5688–5693.

62. Yatim M, Masri S, Desa M, Taib N, Nordin S, Jamal F. Determination of phenotypes and pneumococcal surface protein A family types of Streptococcus pneumoniae from Malaysian healthy children. Journal of Microbiology, Immunology and Infection. 2013; 46(3): 180–186.

63. Subramaniam P, Jabar K, Kee B, Chong C, Nathan A, de Bruyne J, et al. Serotypes & penicillin susceptibility of Streptococcus pneumoniae isolated from children admitted to a tertiary teaching hospital in Malaysia. Indian Journal of Medical Research. 2018; 148(2): 225.

64. Goh SL, Kee BP, Jabar KA, Chua KH, Nathan AM, Bruyne J, et al. Molecular detection and genotypic characterisation of Streptococcus pneumoniae isolated from children in Malaysia. Pathogens and Global Health. 2020; 114(1): 46–54.

65. Arushothy R, Ahmad N, Amran F, Hashim R, Samsudin N, Rosalina Che Azih C. Pneumococcal serotype distribution and antibiotic susceptibility in Malaysia: A four-year study (2014-2017) on invasive paediatric isolates. International Journal of Infectious Diseases. 2019; 80: 129–133.

66. Bahtar A, Abdul Manap R, Ban A. A study of the prevalence of streptococcus pneumoniae serotypes in patients hospitalised for community-acquired pneumonia. European Respiratory Journal. 2016; 48.

67. Cleary DW, Morris DE, Anderson RA, Jones RA, Alattraqchi AG, Rahman NI, et al. The upper respiratory tract microbiome of indigenous Orang Asli in north-eastern Peninsular Malaysia. NPJ Biofilms and Microbiomes. 2021; 7(1).

68. Dzaraly ND, Desa MNM, Muthanna A, Masri SN, Taib NM, Suhaili Z, et al. Antimicrobial susceptibility, serotype distribution, virulence profile and molecular typing of piliated clinical isolates of pneumococci from east coast, Peninsular Malaysia. Scientific Reports. 2021; 11(8220).

69. Morris DE, McNeil H, Hocknell RE, Anderson, Tuck AC, Tricarico S, et al. Carriage of upper respiratory tract pathogens in rural communities of Sarawak, Malaysian Borneo. Pneumonia. 2021; 13(6).

70. Ngoi ST, Muhamad AN, Teh CSJ, Chong CW, Jabar KA, Chai LC, et al. β-Lactam Resistance in Upper Respiratory Tract Pathogens Isolated from a Tertiary Hospital in Malaysia. Pathogens. 2021; 10(12): 1602.

71. Yusof HA, Desa MNM, Masri SN, Malina O, Jamal F. Hyaluronatelyase production by Streptococcus pneumoniae isolated from patients and carriers. Tropical Biomedicine. 2015; 32(3): 413–418.

72. Htoon TT, Oo NAT, Aung WW, Thu HM, Mon M, Yamaguchi M, et al. Serotype Distribution and Antimicrobial Susceptibility Profile of Streptococcus pneumoniae Among Myanmar Children with Acute Respiratory Infection. International Journal of Bacteriology & Parasitology. 2018; 2018(1).

73. Turner P, Melchiorre S, Moschioni M, Barocchi MA, Turner C, Watthanaworawit W, et al. Assessment of Streptococcus pneumoniae pilus islet-1 prevalence in carried and transmitted isolates from mother– infant pairs on the Thailand–Burma border. Clinical Microbiology and Infection. 2012; 18(10): 970–975.

74. Turner P, Hinds J, Turner C, Jankhot A, Gould K, Bentley SD, et al. Improved detection of nasopharyngeal cocolonization by multiple pneumococcal serotypes by use of latex agglutination or molecular serotyping by microarray. Journal of Clinical Microbiology. 2011; 49(5): 1784–1789.

75. Turner P, Turner C, Jankhot A, Phakaudom K, Nosten F, Goldblatt D. Field Evaluation of Culture plus Latex Sweep Serotyping for Detection of Multiple Pneumococcal Serotype Colonisation in Infants and Young Children. PLoS One. 2013; 8(7).

76. Yamaguchi M, Win HPM, Higashi K, Ono M, Hirose Y, Motooka D, et al. Epidemiological analysis of pneumococcal strains isolated at Yangon Children’s Hospital in Myanmar via whole-genome sequencing-based methods. Microbial Genomics. 2021; 7(2): 000523.

77. Capeding MR, Sombrero LT, Lucero MG, Saniel MC. Serotype distribution and antimicrobial resistance of invasive Streptococcus pneumoniae isolates in Filipino children. Journal of Infectious Diseases. 1994; 169(2): 479–480.

78. Capeding MR, Bravo L, Santos J, Kilgore PE, Kim SA, Balter I, et al. Prospective surveillance study of invasive pneumococcal disease among urban children in the Philippines. The Pediatric Infectious Disease Journal. 2013; 32(10): e383–389.

79. Lupisan SP, Herva E, Sombrero LT, Quiambao BP, Capeding MRZ, Abucejo PE, et al. Invasive bacterial infections of children in a rural province in the central Philippines. The American Journal of Tropical Medicine and Hygiene. 2000; 62(3): 341–346.

80. Sombrero L, Nissinen A, Esparar G, Lindgren M, Siira L, Virolainen A&ObotAc. Low incidence of antibiotic resistance among invasive and nasopharyngeal isolates of Streptococcus pneumoniae from children in rural Philippines between 1994 and 2000. European Journal of Clinical Microbiology & Infectious Diseases. 2008; 27(10).

81. Sia SB, Lagrada ML, Gayeta JM, Masim MAL, Abad JP, Magbanua MA, et al. Serotype distribution and antimicrobial resistance of Streptococcus pneumoniae in the Philippines, 2012–2018. Western Pacific Surveillance and Response Journal. 2021; 12(4): 1–8.

82. Wright MS, McCorrison J, Gomez AM, Beck E, Harkins D, Shankar J, et al. Strain Level Streptococcus Colonization Patterns during the First Year of Life. Frontiers in Microbiology. 2017; 8: 1661.

83. Chong CY, Koh-Cheng T, Yee-Hui M, Nancy TWS. Invasive pneumococcal disease in Singapore children. Vaccine. 2008; 26(27-28): 3427–3431.

84. Hsu LY, Lui SW, Lee JL, Hedzlyn HMM, Kong DHL, Shameen S, et al. Adult invasive pneumococcal disease pre- and peri-pneumococcal conjugate vaccine introduction in a tertiary hospital in Singapore. Journal of Medical Microbiology. 2009; 58(1): 101–104.

85. Jauneikaite E, Jefferies JMC, Churton NWV, Lin RTP, Hibberd ML, Clarke SC. Genetic diversity of Streptococcus pneumoniae causing meningitis and sepsis in Singapore during the first year of PCV7 implementation. Emerging Microbes and Infections. 2014; 3(6): e39.

86. Jefferies JMC, Tee WSN, Clarke SC. Molecular analysis of Streptococcus pneumoniae clones causing invasive disease in children in Singapore. Journal of Medical Microbiology. 2011; 60(6).

87. Soh SWL, Poh CL, Lin RVTP. Serotype Distribution and Antimicrobial Resistance of Streptococcus pneumoniae Isolates from Pediatric Patients in Singapore. Antimicrobial Agents and Chemotherapy. 2000; 44(8): 2193–2196.

88. Thoon KC, Chong CY, Tee NWS. Early impact of pneumococcal conjugate vaccine on invasive pneumococcal disease in Singapore children, 2005 through 2010. International Journal of Infectious Disease. 2012; 16(3): e209–e215.

89. Vasoo S, Singh K, Hsu LY, Chiew YF, Chow C, Lin RTP, et al. Increasing antibiotic resistance in Streptococcus pneumoniae colonizing children attending day-care centres in Singapore. Respirology. 2011; 16(8): 1241–1248.

90. Martinez-Vega R, Jauneikaite E, Thoon KC, Chua HY, Chua AH, Khong WX, et al. Risk factor profiles and clinical outcomes for children and adults with pneumococcal infections in Singapore: A need to expand vaccination policy? PLoS ONE. 2019; 14(10): e0220951.

91. Dejsirilert S, Overweg K, Sluijter M, Saengsuk L, Gratten M, Ezaki T, et al. Nasopharyngeal carriage of penicillin-resistant Streptococcus pneumoniae among children with acute respiratory tract infections in Thailand: a molecular epidemiological survey. Journal of Clinical Microbiology. 1999; 37(6): 1832–1838.

92. Levine S, Dejsirilert S, Sangsuk L, Chantra S, Feikin DR, Dowell SF, et al. Serotypes and antimicrobial resistance of streptococcus pneumoniae in Thailand 2002-2004. The Pediatric Infectious Disease Journal. 2006; 25(2): 176–178.

93. Salter SJ, Turner C, Watthanaworawit W, de Goffau MC, Wagner J, Parkhill J, et al. A longitudinal study of the infant nasopharyngeal microbiota: The effects of age, illness and antibiotic use in a cohort of South East Asian children. PLOS NEGLECTED TROPICAL DISEASES. 2017; 11(10): e0005975.

94. Lochindarat S, Teeratakulpisarn J, Warachit B, Chanta C, Thapa K, Gilbert GL, et al. Bacterial etiology of empyema thoracis and parapneumonic pleural effusion in Thai children aged less than 16 years. The Southeast Asian Journal of Tropical Medicine and Public Health. 2014; 45(2): 442–454.

95. Phongsamart W, Srifeungfung S, Dejsirilert S, Chatsuwan T, Nunthapisud P, Treerauthaweeraphong V, et al. Serotype distribution and antimicrobial susceptibility of S. pneumoniae causing invasive disease in Thai children younger than 5 years old, 2000–2005. Vaccine. 2007; 25(7): 1275–1280.

96. Phongsamart W, Srifeungfung S, Chatsuwan T, Nunthapisud P, Treerauthaweeraphong V, Rungnobhakhun P, et al. Changing trends in serotype distribution and antimicrobial susceptibility of Streptococcus pneumoniae causing invasive diseases in Central Thailand, 2009–2012. Human Vaccines & Immunotherapeutics. 2014; 10(7): 1866–1873.

97. Rhodes J, Dejsirilert S, Maloney SA, Jorakate P, Kaewpan A, Salika P, et al. Pneumococcal Bacteremia Requiring Hospitalization in Rural Thailand: An Update on Incidence, Clinical Characteristics, Serotype Distribution, and Antimicrobial Susceptibility, 2005-2010. PLOS ONE. 2013; 8(6): e66038.

98. Srifeungfung S, Chokephaibulkit K, Tribuddharat C. Serotypes and antimicrobial susceptibilities of Streptococcus pneumoniae isolated from hospitalized patients in Thailand. The Southeast Asian Journal of Tropical Medicine and Public Health. 2007; 38(3): 469–477.

99. Srifeungfung S, Tribuddharat C, Comerungsee S, Chatsuwan T, Treerauthanaweeraphong V, Rungnobhakhun P, et al. Serotype coverage of pneumococcal conjugate vaccine and drug susceptibility of Streptococcus pneumoniae isolated from invasive or non-invasive diseases in central Thailand, 2006– 2009. Vaccine. 2010; 28(19): 3440–3444.

100. Srifeungfung S, Phongsamart W, Tribuddharat C, Chatsuwan T, Rungnobhakhun P, Sapcharoen S, et al. Serotype distribution and antibiotic susceptibility of invasive Streptococcus pneumoniae isolates in patients aged 50 years or older in Thailand. Human Vaccines & Immunotherapeutics. 2014; 10(1): 40–44.

101. Wangirapan A, Ayuthaya SIN, Katip W, Kasatpibal N, Mektrirat R, Anukool U, et al. Serotypes and Vaccine Coverage of Streptococcus Pneumoniae Colonization in the Nasopharynx of Thai Children in Congested Areas in Chiang Mai. Pathogens. 2020; 9(12): 988.

102. Baggett HC, Peruski LF, Olsen SJ, Thamthitiwat S, Rhodes J, Dejsirilert S, et al. Incidence of Pneumococcal Bacteremia Requiring Hospitalization in Rural Thailand. Clinical Infectious Diseases. 2009; 48(2): S65–S74.

103. Suphanklang J, Santimaleeworagun W, Thunyaharn S, Traipattanakul J. Pneumococcal meningitis at a Thai hospital over a 10-year period: clinical outcomes, serotypes, and antimicrobial susceptibility patterns. The Southeast Asian Journal of Tropical Medicine and Public Health. 2017; 48(6): 1281–1289.

104. Suwanpakdee D, Samakoses R, Sirinavin S, Kerdpanich A, Simasathien S, Thunyaharn S, et al. Invasive pneumococcal disease in Phramongkutklao Hospital 2004-2008: clinical data, serotype distribution and antimicrobial resistance patterns. Journal of the Medical Association of Thailand. 2010; 93(5): S40–50.

105. Watanabe H, Asoh N, Hoshino K, Watanabe K, Oishi K, Kositsakulchai W, et al. Antimicrobial susceptibility and serotype distribution of Streptococcus pneumoniae and molecular characterization of multidrug-resistant serotype 19F, 6B, and 23F Pneumococci in northern Thailand. Journal of Clinical Microbiology. 2003; 41(9): 4178–4183.

106. Piralam B, Prosperi C, Thamthitwat S, Bunthi C, Sawatwong P, Sangwichian O, et al. Pneumococcal colonization prevalence and density among Thai children with severe pneumonia and community controls. PLOS ONE. 2020; 15(4): e0232151.

107. Chongtrakool P, Aphiyakul Y, Choochur P, Pummangura C, Srifuengfung M, So-Ngern A, et al. Prevalence and Antimicrobial Susceptibility of Streptococcus pneumoniae Isolated from Hospital in Thailand between 2016 and 2020. JOURNAL OF THE MEDICAL ASSOCIATION OF THAILAND. 2022; 105(2): 113–120.

108. Phongsamart W, Srifeungfung S, Chatsuwan T, Rungnobhakhun P, Maleesatharn A, Chokephaibulkit K. Streptococcus pneumoniae Causing Invasive Diseases in Children and Adults in Central Thailand, 2012– 2016. Vaccines (Basel). 2022; 10(8): 1368.

109. Bogaert D, Ha NT, Sluijter M, Lemmens N, De Groot R, Hermans PWM. Molecular epidemiology of pneumococcal carriage among children with upper respiratory tract infections in Hanoi, Vietnam. Journal of Clinical Microbiology. 2002; 40(11): 3903–3908.

110. Dhoubhadel BG, Yasunami M, Nguyen HAT, Suzuki M, Vu TH, Nguyen ATT, et al. Bacterial Load of Pneumococcal Serotypes Correlates with Their Prevalence and Multiple Serotypes Is Associated with Acute Respiratory Infections among Children Less Than 5 Years of Age. PLOS ONE. 2014; 9(10): e110777.

111. Parry CM, Diep TS, Wain J, Hoa NTT, Gainsborough M, Nga D, et al. Nasal Carriage in Vietnamese Children of Streptococcus pneumoniae Resistant to Multiple Antimicrobial Agents. Antimicrobial Agents and Chemotherapy. 2000; 44(3): 484–488.

112. Nguyen HAT, Fujii H, Vu HTT, Parry CM, Dang AD, Ariyoshi K, et al. An alarmingly high nasal carriage rate of Streptococcus pneumoniae serotype 19F non-susceptible to multiple beta-lactam antimicrobials among Vietnamese children. BMC Infectious Diseases. 2019; 19(1).

113. Schultsz C, Vien LM, Campbell JI, Chau NVV, Diep TS, Hoang NVM, et al. Changes in the nasal carriage of drug-resistant Streptococcus pneumoniae in urban and rural Vietnamese schoolchildren. Transactions of the Royal Society of Tropical Medicine and Hygiene. 2007; 101: 484–492.

114. Vu HTT, Yoshida LM, Suzuki M, Nguyen HAT, Nguyen CDL, Nguyen ATT, et al. Association Between Nasopharyngeal Load of Streptococcus pneumoniae, Viral Coinfection, and Radiologically Confirmed Pneumonia in Vietnamese Children. The Pediatric Infectious Disease Journal. 2011; 30(1): 11–18.

115. Watanabe K, Anh DD, Huong PLT, Nguyet NT, Anh NTH, Thi NT, et al. Drug-resistant pneumococci in children with acute lower respiratory infections in Vietnam. Pediatrics International. 2008; 50: 514–518.

116. Parry CM, Duong NM, Zhou J, Main NTH, Diep TS, Thinh LQ, et al. Emergence in Vietnam of Streptococcus pneumoniae Resistant to Multiple Antimicrobial Agents as a Result of Dissemination of the Multiresistant Spain23F-1 Clone. Antimicrobial Agents and Chemotherapy. 2002; 46(11): 3512–3517.

117. Higgins RA, Temple B, Dai VTT, Phan TV, Toan NT, Spry L, et al. Immunogenicity and impact on nasopharyngeal carriage of a single dose of PCV10 given to vietnamese children at 18 months of age. The Lancet Regional Health - Western Pacific. 2021; 16: 100273.

118. Mohamed YH, Toizumi M, Uematsu M, Nguyen HAT, Le LT, Takegata M, et al. Prevalence of Streptococcus pneumoniae in conjunctival flora and association with nasopharyngeal carriage among children in a Vietnamese community. Scientific Reports. 2021; 11(1): 337.

119. Son BA, Hai TX, Cuong TV, Chinh DD, Le THH, Dung NM, et al. Serotype distribution and antibiotic resistance of Streptococcus pneumoniae isolates collected from unvaccinated children with pneumonia at a province in central Vietnam. Iranian Journal of Microbiology. 2022; 14(5): 653–661.

120. Toizumi M, Satoh C, Quilty BJ, Nguyen HAT, Madaniyazi L, Le LT, et al. Effect of pneumococcal conjugate vaccine on prevalence of otitis media with effusion among children in Vietnam. Vaccine. 2022; 40(36): 5366–5375.

121. Song JH, Lee NY, Ichiyama S, Yoshida R, Hirakata Y, Fu Y, et al. Spread of drug-resistant Streptococcus pneumoniae in Asian countries: Asian Network for Surveillance of Resistant Pathogens (ANSORP) Study. Clinicl Infectious Diseases. 1999; 28(6): 1206–1211.

122. Kim SH, Song JH, Chung DR, Thamlikitkul V, Yang Y, Wang H, et al. Changing trends in antimicrobial resistance and serotypes of Streptococcus pneumoniae isolates in Asian countries: an Asian Network for Surveillance of Resistant Pathogens (ANSORP) study. Antimicrobial Agents and Chemotherapy. 2012; 56(3): 1418–26.

123. Song J, Jung S, Ko KS, Kim NY, Son JS, Chang H, et al. High Prevalence of Antimicrobial Resistance among Clinical Streptococcus pneumoniae Isolates in Asia (an ANSORP Study). Antimicrobial Agents and Chemotherapy. 2004; 48(6): 2101–2107.

124. Lee NY, Song JH, Kim S, Peck KR, Ahn KM, Lee SI, et al. Carriage of Antibiotic-Resistant Pneumococci among Asian Children: A Multinational Surveillance by the Asian Network for Surveillance of Resistant Pathogens (ANSORP). Clinical Infectious Diseases. 2001; 32(10): 1463–1469.

125. Kim S, Chung D, Song J, Baek J, Thamlikitkul V, Wang H, et al. Changes in serotype distribution and antimicrobial resistance of Streptococcus pneumoniae isolates from adult patients in Asia: Emergence of drug resistance non-vaccine serotypes. Vaccine. 2020; 38(38): 6065–6073.

126. Johnson HL, Deloria-Knoll M, Levine OS, Stoszek SK, Hance LF, Reithinger R, et al. Systematic Evaluation of Serotypes Causing Invasive Pneumococcal Disease among Children Under Five: The Pneumococcal Global Serotype Project. PLoS Medicine. 2010; 7(10): e1000348.

127. Clifford S, Knoll MD, O’Brien KL, Pollington TM, Moodley R, Prieto-Merino D, et al. Global landscape of Streptococcus pneumoniae serotypes colonising healthy individuals worldwide before vaccine introduction; a systematic review and meta-analysis. MedRxiv. 2023.

128. Song JH, Dagan R, Klugman KP, Fritzell B. The relationship between pneumococcal serotypes and antibiotic resistance. Vaccine. 2012; 30(17): 2728–2737.

129. Lo S, Gladstone RA, van Tonder AJ, Lees JA, du Plessis M, Benisty R, et al. Pneumococcal lineages associated with serotype replacement and antibiotic resistance in childhood invasive pneumococcal disease in the post-PCV13 era: an international whole-genome sequencing study. The Lancet Infectious Diseases. 2019; 19(7): 759–769.

130. Whitney CG, Farley MM, Hadler J, Harrison LH, Bennett NM, Lynfield R, et al. Decline in Invasive Pneumococcal Disease after the Introduction of Protein–Polysaccharide Conjugate Vaccine. The New England Journal of Medicine. 2003; 348: 1737–1746.

131. Flasche S, Jan Van Hoek A, Sheasby E, Waight P, Andrews N, Sheppard C, et al. Effect of Pneumococcal Conjugate Vaccination on Serotype-Specific Carriage and Invasive Disease in England: A Cross-Sectional Study. PLoS Medicine. 2011; 8(4): e1001017.

132. Zhao W, Pan F, Wang B, Wang C, Sun Y, Zhang T, et al. Epidemiology Characteristics of Streptococcus pneumoniae From Children With Pneumonia in Shanghai: A Retrospective Study. Frontiers n Cellular and Infection Microbiology. 2019; 9.

133. Brueggemann AB, Peto TEA, Crook DW, Butler JC, Kristinsson KG, Spratt BG. Temporal and Geographic Stability of the Serogroup-Specific Invasive Disease Potential of Streptococcus pneumoniae in Children. The Journal of Infectious Diseases. 2004; 190(7): 1203–1211.

134. Yoon JG, Jang AY, Kim MJ, Seo YB, Lee J, Choi YH, et al. Persistent serotype 3 and 19A invasive pneumococcal diseases in adults in vaccine era: Serotype-dependent difference in ceftriaxone susceptibility. Vaccine. 2022; 40(15): 2258–2265.

135. Kandasamy r, Lo S, Gurung M, Carter MJ, Gladstone R, Lees J, et al. Effect of childhood vaccination and antibiotic use on pneumococcal populations and genome-wide associations with disease among children in Nepal: an observational study. The Lancet Microbe. 2022; 3(7): e503–e511.

136. Rahman T, de Gier C, Orami T, Seppanen EJ, Granland CM, Francis JP, et al. PCV10 elicits Protein D IgG responses in Papua New Guinean children but has no impact on NTHi carriage in the first two years of life. Vaccine. 2021; 39(26): 3486–3492.

137. Hausdorff WP, Hoet B, Schuerman L. Do pneumococcal conjugate vaccines provide any cross-protection against serotype 19A? BMC Pediatrics. 2010; 10.

138. Chen HH, Hsu MH, Wu TL, Li HC, Chen CL, Janapatla RP, et al. Non-typeable Streptococcus pneumoniae infection in a medical center in Taiwan after wide use of pneumococcal conjugate vaccine. Journal of Microbiology, Immunology abd Infection. 2020; 53(1): 94–98.

139. Marks LR, Reddinger RM, Hakansson AP. High Levels of Genetic Recombination during Nasopharyngeal Carriage and Biofilm Formation in Streptococcus pneumoniae. mBio. 2012; 3(5): e00200–e00212.

140. Richardson A, Morris DE, Clarke SC. Vaccination in Southeast Asia—Reducing meningitis, sepsis and pneumonia with new and existing vaccines. Vaccine. 2014; 32(33): 4119–4123.

141. Lee KS, Ming LC, Lean QY, Yee SM, Patel R, Taha NA, et al. Cross-border Collaboration to Improve Access to Medicine: Association of Southeast Asian Nations Perspective. Journal of Epidemiology and Global Health. 2019; 9(2): 93–97.

142. Coughtrie AL, Jefferies JM, Cleary D, Doncaster CP, Faust S, Kraaijeveld A, et al. Microbial epidemiology and carriage studies for the evaluation of vaccines. Journal of Medical Microbiology. 2019; 68(10).

143. Lister AJJ, Le CF, Cheah ESG, Desa MNM, Cleary DW, Clarke SC. Serotype distribution of invasive, non-invasive and carried Streptococcus pneumoniae in Malaysia: a meta-analysis. Pneumonia. 2021; 13(9).

